# Interaction studies of risk proteins in human induced neurons reveal convergent biology and novel mechanisms underlying autism spectrum disorders

**DOI:** 10.1101/2021.10.07.21264575

**Authors:** Greta Pintacuda, Yu-Han H. Hsu, Kalliopi Tsafou, Ka Wan Li, Jacqueline M. Martín, Jackson Riseman, Julia C. Biagini, Joshua K.T. Ching, Miguel A. Gonzalez-Lozano, Shawn B. Egri, Jake Jaffe, August B. Smit, Nadine Fornelos, Kevin C. Eggan, Kasper Lage

## Abstract

Sequencing studies of autism spectrum disorders (ASDs) have identified numerous risk genes with enriched expression in the human brain, but it is still unclear how these genes converge into cell type-specific networks and how their encoded proteins mechanistically contribute to ASDs. To address this question, we performed brain cell type-specific interaction proteomics to build a protein-protein interaction network for 13 ASD risk genes in human excitatory neurons derived from iPS cells. The network contains many (>90%) interactions not reported in the literature and is enriched for transcriptionally perturbed genes observed in layer 2/3 cortical neurons of ASD patients, indicating that it can be explored for ASD-relevant biological discovery. We leveraged the network dataset to show that the brain-specific isoform of ANK2 is important for its interactions with synaptic proteins and characterized a PTEN-AKAP8L interaction that influences neuronal growth through the mTOR pathway. The IGF2BP1-3 complex emerges as a point of convergence in the network, and we showed that this complex is involved in a transcriptional circuit concentrating both common and rare variant risk of ASDs. Finally, we found the network itself enriched for ASD rare variant risk, indicating that it can complement genetic datasets for prioritizing additional risk genes. Our findings establish brain cell type-specific interactomes as an organizing framework to facilitate interpretation of genetic and transcriptomic data in ASDs and illustrate how both individual and convergent interactions lead to biological insights into the disease.

## Introduction

Autism spectrum disorders (ASDs) comprise a group of heritable neurodevelopmental conditions associated with challenges in social interaction, cognition, and behavior. In recent years, substantial progress has been made in understanding the genetic architecture of ASDs, establishing that their clinical manifestations reflect polygenic contributions from both common and rare genetic risk variants across a wide spectrum of allele frequencies (*1–9*). While common variants account for most of ASD heritability (*3*), rare variants have larger effect sizes, affect protein-coding genes (*10*), and are therefore more easily amenable to reductionistic experimental analyses and functional interpretation (*11*). However, even though some ASD risk genes implicated by rare variants immediately incriminate specific physiological processes and molecular mechanisms, such as chromatin remodeling and synaptic morphogenesis (*12, 13*), the majority do not individually offer a comprehensive picture of the biology of autism. More broadly, it remains to be determined to which extent ASD risk genes integrate into high-order pathways and networks (hereafter, we will collectively refer to these as networks) during neurodevelopment (*14, 15*) and which insights grouping genes in such cell type specific networks will offer.

Integration of genetic and protein-protein interaction (PPI) data has shown that genes implicated in immunological, metabolic, and cardiovascular diseases often converge into PPI networks (*16–19*). In the context of ASDs, early studies of *de novo* mutations and structural variation in risk genes also suggested that their encoded proteins form PPI networks (*20–22*). These studies establish that PPIs can provide a systematic framework for biological interpretation of ASD risk genes, however, they overwhelmingly relied on PPI data derived from public databases and thus lack cell type-specificity, as experiments from highly proliferative and immortalized cell types (e.g., HEK or cancer cell lines) are vastly over-represented in these databases (*18, 23, 24*).

Recent developments in genetics, transcriptomics, and induced pluripotent stem cell (iPSC) technology have now opened up the possibility to address this limitation. Indeed, integration of genetic and transcriptomic datasets has shown that genes expressed in excitatory neurons are particularly enriched for genetic risk in ASDs (*10, 25*). Furthermore, analyses of brain gene co-expression and regulatory networks (*26*), as well as single-cell RNA sequencing in *postmortem* brain tissues from patients and controls (*27–30*), have also identified cortical excitatory neurons as a key cell type in ASDs. In tandem, new iPSC protocols for producing large numbers of induced neurons (iNs) that resemble cortical excitatory neurons, provide the opportunity to systematically execute interaction proteomics in an *in vitro* neuronal cell model of key relevance to ASDs (*31, 32*).

In this study, we brought together emerging catalogs of risk genes, a cell model of human cortical excitatory neurons, proteomic approaches, and computational modeling to build a brain cell type-specific PPI network for ASD risk proteins. Specifically, through data integration of 26 immunoprecipitation (IP) experiments of 13 ASD risk proteins performed in iNs, we generated a combined brain cell type-specific PPI network in which >90% of the protein interactions are unreported in the literature, likely due to their selective expression in the brain. These data are nevertheless highly reproducible (>91% validation rate) and strongly overlap with interactions of the same proteins identified from IPs performed in human brain homogenates, indicating that iNs can capture PPIs present in analogous complex human primary tissues. As further support for the iN cell model, we found the network significantly enriched for transcriptionally perturbed genes in layer 2/3 cortical neurons of non-genetically profiled ASD patients, which agrees with previous evidence that specifically implicates this key cell type in ASD pathophysiology. To exemplify how perturbing specific PPIs in the network can lead to mechanistic insights and ASD-relevant cellular phenotypes, we showed that ablating a giant brain-specific ANK2 alternative exon, which concentrates mutations observed in ASD patients, resulted in loss of isoform-specific interactions with synaptic proteins, while disrupting a newly found PTEN-AKAP8L interaction affected neuronal growth. Furthermore, convergent signals in the network can also provide important insights, as illustrated by the mRNA-binding complex IGF2BP1-3, which is connected to multiple ASD risk proteins in the network and is at the center of a transcriptional circuit enriched for both common and rare variant risks of ASDs. Finally, the network is also enriched for genes containing rare ASD-associated variants, indicating that it can complement large-scale genetic studies to prioritize additional risk genes.

In summary, our study demonstrates how brain cell type-specific interaction proteomics represents a rich and currently underutilized opportunity for biological discovery in ASDs, providing a rich community resource to catalyze interpretation of genetic and scRNA datasets. Our results lay a foundation for future discovery of biomarkers or targets for therapeutic intervention.

## Results

### ASD risk genes and proteins are expressed in induced neurons

A recent exome sequencing study of individuals on the autism spectrum identified over 100 ASD-associated genes using combined *de novo* and case-control analyses (*10*). 24 of these genes (i.e., *ADNP*, *ANK2*, *ANKRD11*, *ARID1B*, *ASH1L*, *CHD2*, *CHD8*, *CTNNB1*, *DEAF1*, *DSCAM*, *DYRK1A*, *FOXP1*, *GIGYF1*, *GRIN2B*, *KDM6B*, *KMT5B*, *MED13L*, *POGZ*, *PTEN*, *SCN2A*, *SHANK3*, *SLC6A1*, *SYNGAP1*, and *TLK2*) passed the statistical threshold of exome-wide significant enrichment for rare deleterious mutations. We used this set of 24 genes as the starting point of our workflow to generate an ASD PPI network in iNs (**Figure 1A**), and hereafter refer to them and their corresponding proteins as ‘index genes’ and ‘index proteins’, respectively.

**Figure 1.**
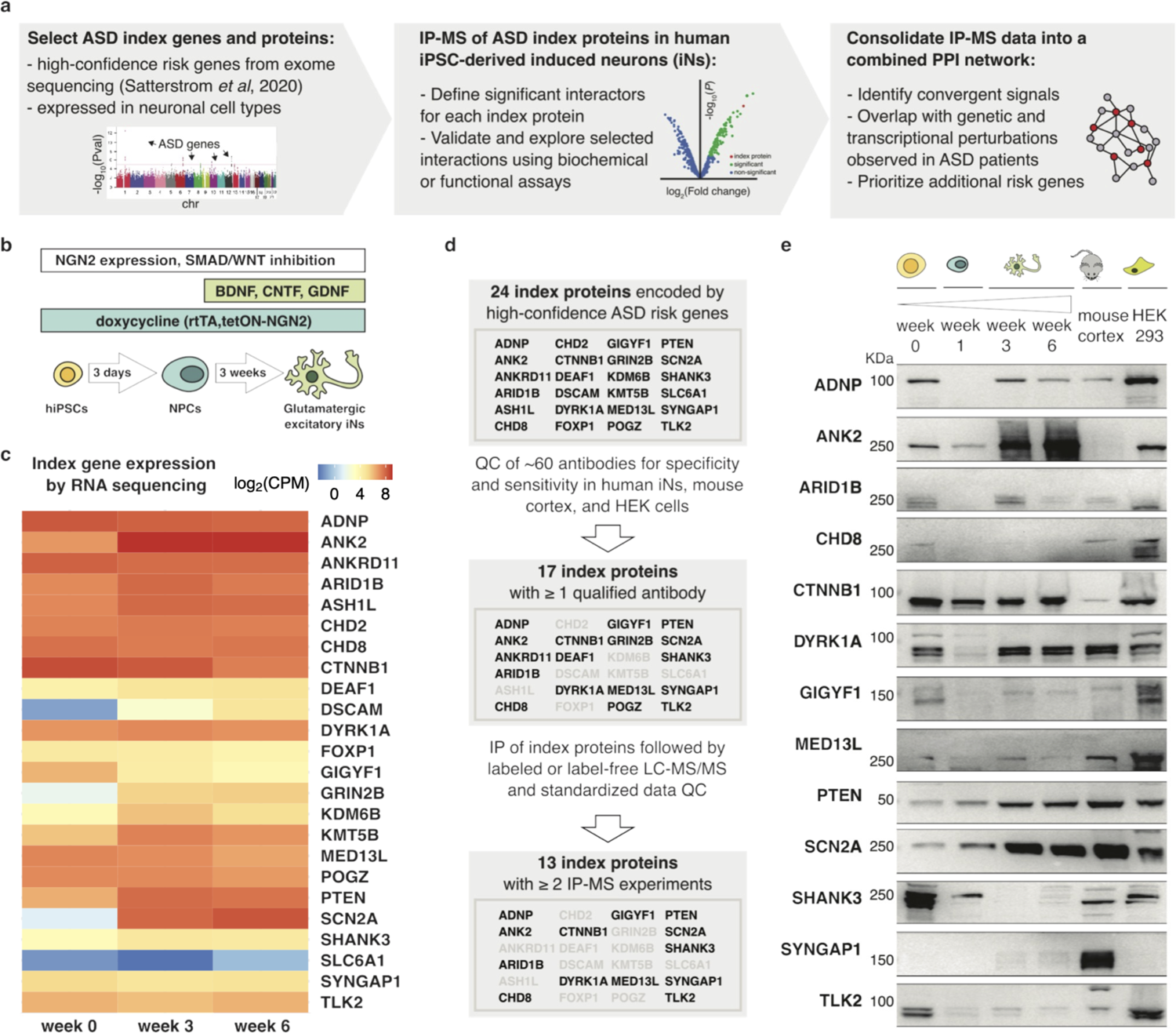
Study workflow and expression of ASD index genes and proteins in human iNs. **(a)** Schematic of the experimental and computational workflow to generate an ASD PPI network in human iNs. **(b)** Schematic of the key steps in the glutamatergic-patterning protocol utilized to generate neural progenitor cells (NPCs) and excitatory iNs from human iPSCs. **(c)** Heatmap of ASD index gene expression over the course of neuronal maturation. Bulk RNA-seq analysis was performed on iPSCs (week 0) and after 3 and 6 weeks of neuronal maturation according to the protocol described in (b). CPM=normalized counts per million; maximum log_2_(CPM) was capped at 9 for visualization. All index genes except *SLC6A1* had robustly detectable expression after week 3. **(d)** Flowchart representing the selection of index proteins studied in this work. Only proteins expressed in iNs for which IP-competent commercial antibodies are available were selected for downstream experiments. **(e)** Expression of 13 index proteins in differentiating iPSCs (as described in [b]), mouse cortex, and HEK293 cells detected by immunoblotting. The antibody utilized against ANK2 recognizes a human-specific epitope and SYNGAP1 is not expressed in HEK293 cells. Molecular weights (KDa) are marked on the left of each blot.

As a substrate for cell type-specific analyses of the index genes and proteins, we adopted and further optimized NGN2 programming combined with developmental patterning as previously described by Nehme *et al.* (*31*) to differentiate iPSCs into iNs. In particular, we used an iPS cell line (iPS3, **Methods**) where the neurogenic factor *Neurogenin 2* (*NGN2*) is integrated in the genome under a tetON promoter for rapid and controlled doxycycline-induced expression, resulting in highly homogeneous iN populations (**Figure 1B**).

We measured the expression of ASD index genes through RNA sequencing of cells during weeks 0, 3, and 6 of neuronal differentiation (**Figure 1C**). The expression patterns of these genes varied across time, with some genes showing decreased (e.g., *GIGYF1*) or increased (e.g., *ANK2*) expression levels at later time points. Generally, most index genes were expressed after three weeks of differentiation, including genes (i.e., *GRIN2B*, *SCN2A*, and *SHANK3*) with previously reported neuronal identity (*33*). Importantly, gene expression observed in iNs as early as week 3 (**Figure 1C**) resembled expression patterns seen in excitatory neuron populations of the human cortex (**Supplementary Figure 1A,B**) (*30*), supporting this cell model as a proxy for cell types in the human brain.

Next, we sought to identify immunoreagents to detect and immunoprecipitate index proteins in iNs. After testing ∼60 antibodies (**Supplementary Table 1**), we concluded that there were no adequate commercial reagents for detection of seven proteins (ASH1L, CHD2, DSCAM, FOXP1, KDM6B, KMT5B, SLC6A1), based on western blots executed on a range of cells and tissues (i.e., mouse cortex and HEK293 cells; **Figure 1D**) included as positive controls. Furthermore, four proteins (ANKRD11, DEAF1, GRIN2B, POGZ) were successfully detected by immunoblotting (**Supplementary Figure 1C**), but no immunoreagent was IP-competent. The remaining 13 proteins could be detected by immunoblotting in the form of at least one protein isoform at week 3 of iN differentiation (**Figure 1E**) and were amenable to IP.

The protein expression analysis revealed that many index proteins show distinct isoform patterns across species (e.g., TLK2 in mouse cortex), neuronal differentiation time points (e.g., SHANK3 in iPSCs), or cell types (e.g., CHD8 in HEK293 cells; **Figure 1E**). This observation highlights the importance of using human iNs in order to capture cell type-specific biochemical interactions of relevance to ASDs.

### Interaction proteomics of index proteins in induced neurons

We executed IPs followed by mass spectrometry (IP-MS) of ADNP, ANK2, ARID1B, CHD8, DYRK1A, FOXP1, MED13L, POGZ, PTEN, SHANK3, SCN2A, SYNGAP1, and TLK2 in iNs at week 4 of differentiation (**Figure 2A,B, Supplementary Figure 2**, and **Supplementary Table 2**). Each experiment was conducted in duplicates using ∼15 million cells per replicate, and protein abundances in the IP samples were quantified using labeled or label-free liquid chromatography followed by tandem mass spectrometry (LC-MS/MS). In total, we conducted 37 IP-MS experiments for the 13 index proteins, generating 2-5 experimental datasets for each protein.

**Figure 2.**
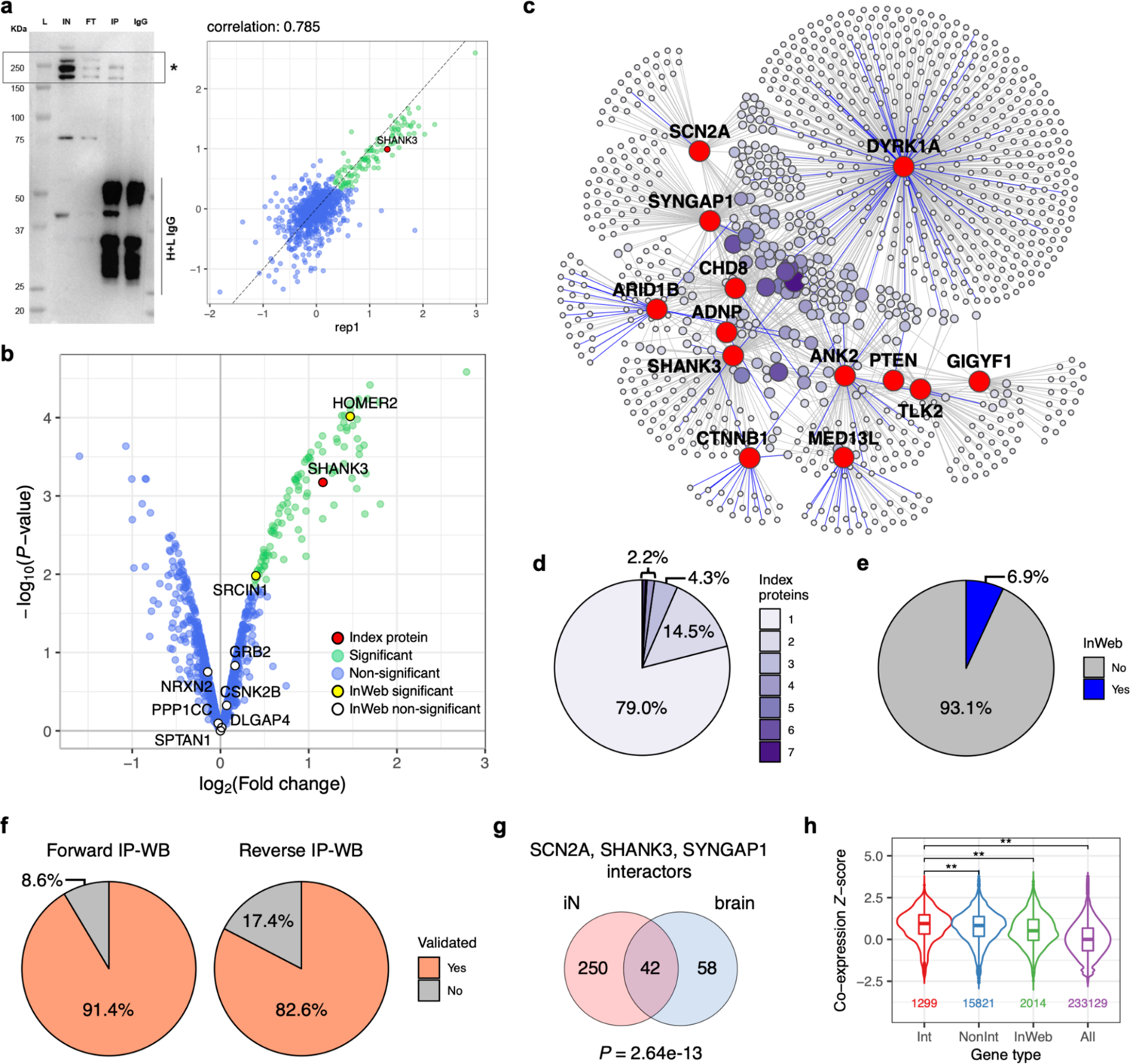
Generation of a combined PPI network for 13 ASD index proteins in iNs. **(a)** Example of a typical IP-MS experiment with SHANK3 as bait protein. Left: immunoblot of a SHANK3 IP with its main isoforms marked by a box and asterisk; L=Ladder, IN=Input, FT=Flow-through, IP=Immunoprecipitation, IgG=IgG control, H+L IgG=Heavy and light IgG chains. Molecular weights are in KDa. Right: log_2_ fold change (FC) correlation between IP-MS replicates. **(b)** Volcano plot of IP-MS experiment from (a), showing SHANK3 in red, its significant interactors (log_2_ FC > 0 and FDR ≤ 0.1) in green, and non-interactors in blue; known InWeb interactors identified as interactors or non-interactors in the experiment are highlighted in yellow or white, respectively. **(c)** Combined PPI network derived from 26 IP-MS experiments performed in iNs. Nodes represent index proteins (red) and their interactors (purple); color intensity and size of the interactor nodes scale with interactor frequency (i.e., how many index proteins are linked with each interactor). Edges indicate observed interactions, with known InWeb interactions highlighted in blue. **(d)** Distribution of interactor frequency in the combined network. **(e)** Distribution of InWeb vs. novel (i.e., non-InWeb) interactions in the combined network. **(f)** Validation rates of a subset of interactions tested in forward or reverse IPs followed by western blotting (IP-WB). **(g)** Overlap between interactors for SCN2A, SHANK3, and SYNGAP1 derived from IP-MS experiments performed in iNs vs. human brain homogenates. Overlap enrichment P-value was calculated using a one-tailed hypergeometric test that only considered the population of proteins detected in both iN and brain IPs. **(h)** Pairwise co-expression Z-scores between index genes and their interactors (Int), non-interactors (NonInt), known InWeb interactors (InWeb), and all protein-coding genes (All) derived from a spatial transcriptomic dataset in human dorsolateral prefrontal cortex. Double asterisks indicate significant difference in score distribution as calculated by two-tailed Wilcoxon rank-sum tests (*P* < 0.05/6, adjusting for 6 possible pairwise comparisons). Number of gene pairs plotted for each gene type is indicated towards the bottom of the plot.

For each IP-MS experiment, we performed quality control (QC) and data analysis using Genoppi (*24*). Specifically, we calculated the log_2_ fold change (FC) and corresponding statistical significance of each protein identified in the index protein IPs compared to control IPs (**Supplementary Table 2**). After filtering out 11 datasets that did not pass QC thresholds (i.e., log_2_ FC correlation between replicates was ≤ 0.6 or the index protein was not positively enriched at FDR ≤ 0.1), 26 high-quality datasets were used for all downstream analyses. In each dataset, we defined proteins with log_2_ FC > 0 and FDR ≤ 0.1 as ‘significant interactors’ of the index protein; the rest of the detected proteins were defined as ‘non-interactors’ (**Supplementary Table 3**). We also systematically intersected each dataset with known protein interactors of the index protein from the InWeb_InBioMap (InWeb) database (*23, 24*), which aggregates human PPI data from > 40k published articles (**Supplementary Table 3)**. This allowed us to assess if the identified interactions have been reported in the literature or are potentially novel.

We exemplify our analysis workflow using an IP of SHANK3: enrichment of SHANK3 in the IP was confirmed by immunoblotting (**Figure 2A**, left), and quantified in the IP-MS data (log_2_ FC = 1.2 and FDR = 0.012; **Figure 2B**). The IP-MS replicate log_2_ FC correlation is 0.79 (**Figure 2A**, right). Only two out of 104 significant interactors were previously reported in InWeb, suggesting that 98% of the interactions reported here are new (**Figure 2B**). Overall, the clear enrichment of SHANK3 both in the immunoblot of the IP and in the MS results, in combination with the high correlation between replicates, support the reproducibility and robustness of our data. The low number of known SHANK3 interactors likely reflects the differences between our experimental design vs. commonly used approaches in previous literature (**Supplementary Text 1**) and supports the use of neuron-specific proteomics to discover new ASD risk gene relationships.

Since the 26 IP-MS datasets for the 13 index proteins that passed QC were generated across three different MS facilities, with 9 index proteins having multiple IPs, we compared the data between facilities and between IPs of the same index protein to evaluate their agreement (**Supplementary Text 2**, **Supplementary Figure 3**, and **Supplementary Table 4**). While we observed differences in terms of data structures and detected proteins in the datasets generated by the three facilities, the significant protein interactions we identified have similar quality and degree of biological relevance regardless of facility origin, suggesting these data are complementary to each other. Moreover, our experimental and computational analyses did not reveal consistent quality differences between index protein interactors identified in only one vs. in multiple IPs (**Supplementary Text 2**) or between interactors detected across different FC ranges (**Supplementary Text 3**). Therefore, we decided to take the ‘union’ of all identified interactors when merging the IP-MS datasets into combined PPI networks in downstream analyses in order to study the most inclusive network for each index protein.

### The ASD PPI network consists of many convergent, novel, and reproducible interactions

After analyzing each IP-MS dataset individually, we merged all the interactors identified across the 26 high-quality datasets to generate a combined PPI network for all 13 index proteins. This network comprises 1,021 proteins interacting with at least one index protein (**Figure 2C** and **Supplementary Table 3**). The PPI network shows high interconnectivity, with > 20% of interactors being shared by more than one index protein (**Figure 2D** and **Supplementary Figure 4A**). Furthermore, whereas the interactors of several index proteins (ANK2, ARID1B, CTNNB1, DYRK1A, MED13L) are enriched for known InWeb interactors (**Supplementary Table 3**), > 90% of all interactions in the network have not been reported in InWeb (**Figure 2E**). Again, this observation was not unexpected given the limited number of neuronal PPI datasets in the literature (**Supplementary Text 1**) and suggests the opportunity for biological discovery linked to further characterization of newly found interactions.

To confirm the robustness and reproducibility of MS-detected interactions, we extensively validated both unique (i.e., interacting with only one index protein) and shared (i.e., interacting with multiple index proteins) interactors in independent IPs by immunoblotting (**Supplementary Figures 4B-D, 9A,C** and **Supplementary Table 5)**. Overall, we validated 65 out of the 71 tested interactions (91.5% validation rate; **Figure 2F**, left). In parallel we also performed reverse IPs using a panel of interactors as baits, and successfully detected the original index proteins in 19 out of 23 reverse IPs that showed bait enrichment (82.6% validation rate; **Figure 2F**, right, **Supplementary Figure 5**, and **Supplementary Table 5**). Together these results are in line with the expected validation rate of ∼90% based on the FDR ≤ 0.1 threshold used to define interactors, and with previously reported validation rates obtained using a similar experimental design in the same cell model (*24*).

To compare our data to protein interactions in the human brain, we performed IP-MS experiments for a subset of index proteins (ANK2, DYRK1A, SCN2A, SHANK3, SYNGAP1) in *postmortem* brain homogenates. On average, brain IP-MS datasets have lower log_2_ FC correlations between replicates compared to the iN IP-MS data (average correlation = 0.63 and 0.72 for brain and iN IPs, respectively; **Supplementary Table 2**). The lower reproducibility is likely reflecting noise and experimental artifacts of interaction proteomics executed in complex samples that comprise a mix of cell types, with variable contributions of ASD-relevant neuronal populations.

After applying the same QC metrics as previously described, we considered three of the brain IPs (for SCN2A, SHANK3, and SYNGAP1) to be high-quality and compared them against the analogous iN-derived IPs. Considering that the brain homogenates contain a mix of cells whereas the iNs are highly homogeneous, we observed good agreement in terms of log_2_ FC correlation for proteins detected in both sample types (median correlation = 0.517 between brain vs. iN IPs, compared to median = 0.628 among iN IPs; **Supplementary Figure 6A**). Furthermore, there is a statistically significant overlap between the interactors identified in the two sample types, with 42% of the brain interactors also identified as interactors in iNs (*P* = 2.6e-13; **Figure 2G** and **Supplementary Figure 6B,C**). Throughout brain development (*34*) (www.brainspan.org), both brain- and iN-derived interactors have elevated gene expression in the cortex compared to random genes; in particular, interactors that were identified in both sample types have an expression profile that is comparable to ASD risk genes identified through exome sequencing (**Supplementary Figure 7A,B**).

To further support the biological relevance of our iN-derived PPI network in complex brain tissues, we also characterized the co-expression patterns of interacting proteins in the network across four independent datasets derived from human or mouse brains using spatial transcriptomic (*35, 36*), single-cell RNA-seq (*30*), or bulk RNA-seq (www.brainspan.org) approaches (**Figure 2H** and **Supplementary Figure 7C**). In general, we found that the index proteins tend to have higher co-expression with their interactors compared to the non-interactors detected in iNs during IP-MS, known InWeb interactors found in mostly non-neuronal context, and all protein-coding genes. Therefore, both the IP-MS data from brain homogenates and the brain co-expression analysis indicate that our iN-derived PPI network can capture gene relationships found in complex tissues in the human brain, and the co-expression results further demonstrate the value of studying the ASD index proteins in a neuronal context.

### The PPI network is transcriptionally perturbed in layer 2/3 cortical excitatory neurons of patients with ASDs

We assessed the tissue specificity of the network by calculating its overlap enrichment with GTEx tissue-specific genes (*37*) compared to the rest of the genome. As expected, the network showed statistically significant enrichment (*P* < 0.05/53, adjusting for 53 tissues) in the central nervous system, as well as in several other tissue categories (**Figure 3A** and **Supplementary Table 6**). When the analysis was repeated using brain region-specific genes, the network showed significant enrichment (*P* < 0.05/13, adjusting for 13 brain regions) in the frontal cortex, cerebellar hemisphere, anterior cingulate cortex, and amygdala (**Figure 3B** and **Supplementary Table 6**). In addition, we performed SynGO (*38*) gene set analysis to confirm that the PPI network is enriched for genes involved in biological processes in the synapse, such as “protein translation at postsynapse” and “synapse organization” (**Supplementary Figure 7D** and **Supplementary Table 6**).

**Figure 3.**
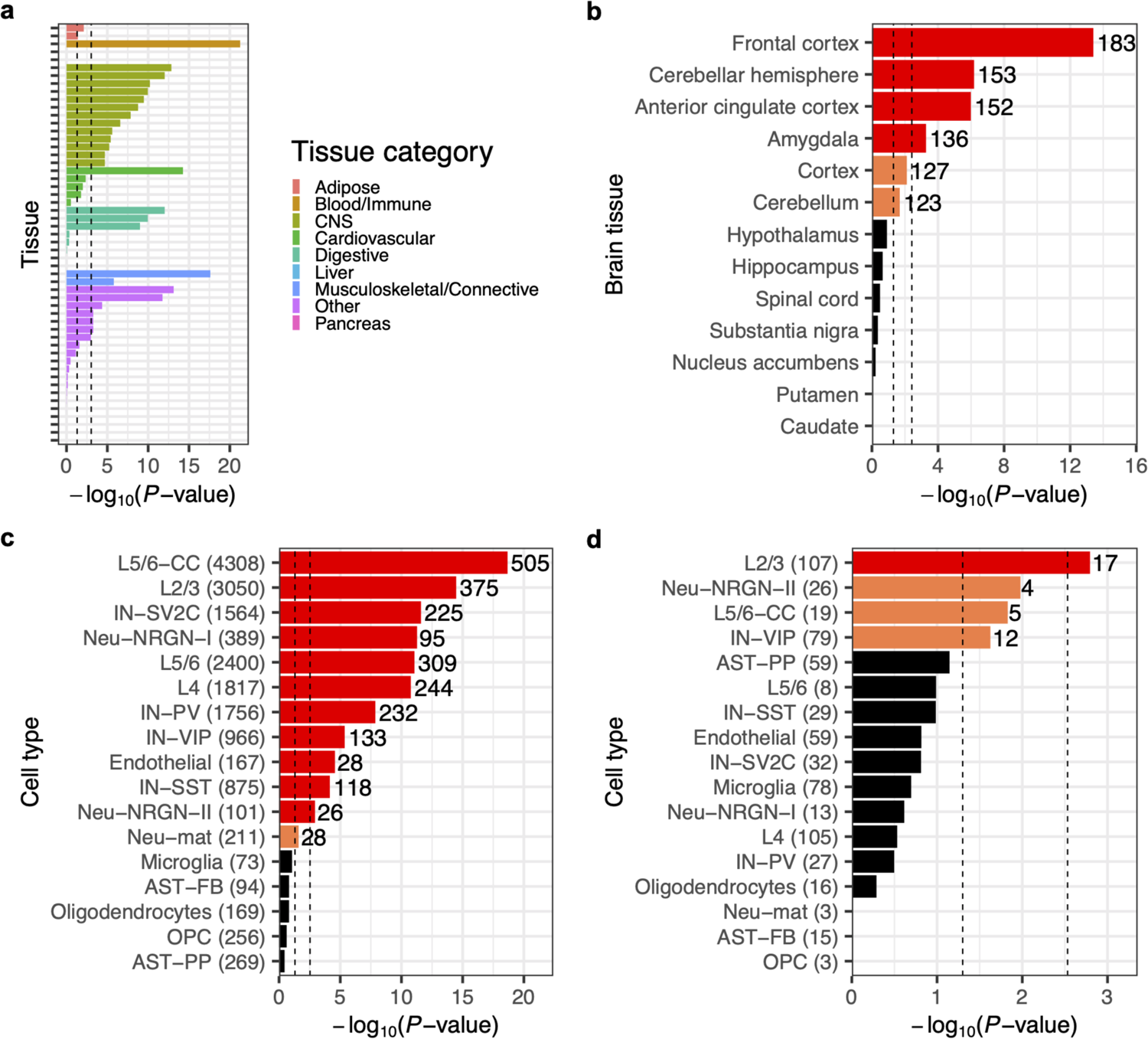
Tissue and cell type enrichment of the ASD PPI network. Overlap enrichment between the network and GTEx tissue-specific genes **(a**; 2484 genes per tissue), GTEx brain region-specific genes **(b**; 2484 genes per tissue), genes expressed in > 50% of cells in each cell type in the *postmortem* cortex **(c**; gene counts in parentheses), and differentially expressed genes in each cell type in the *postmortem* cortex of ASD patients compared to controls **(d**; gene counts in parentheses). All P-values were derived from one-tailed hypergeometric tests; for (a) and (b), a genome-wide background (i.e., all genes in the GTEx data) was used as the ‘population’ in the test; for (c) and (d), a neuronal proteome background (i.e., genes encoding all proteins detected in iNs by IP-MS) was used. Left or right vertical dashed lines indicate nominal (*P* < 0.05) or Bonferroni-corrected (*P* < 0.05/number of tissues or cell types) significance thresholds, respectively; for (b)-(d), nominally or Bonferroni-significant results are highlighted in orange or red, respectively, and labeled with the number of genes in the overlap. Tissue or cell type abbreviations: CNS, central nervous system; AST-FB and AST-PP, fibrous and protoplasmic astrocytes; OPC, oligodendrocyte precursor cells; IN-PV, IN-SST, IN-SV2C, and IN-VIP, parvalbumin, somatostatin, SV2C, and VIP interneurons; L2/3 and L4, layer 2/3 and layer 4 excitatory neurons; L5/6 and L5/6-CC, layer 5/6 corticofugal projection and cortico-cortical projection neurons; Neu-mat, maturing neurons; Neu-NRGN-I and Neu-NRGN-II, NRGN-expression neurons.

To link the network to specific cell types that are implicated in ASDs, we integrated our data with a recent single-cell RNA sequencing study (*30*), which measured gene expression in the *postmortem* cortex of ASD patients versus healthy controls and identified differentially expressed genes (DEGs) between the two groups in 17 annotated cell types. First, we performed a ‘global’ enrichment analysis to test whether the network genes show enriched overlap with commonly expressed genes in a cell type (i.e., expressed in >50% of cells in the cell type) compared to other genes in the genome. We also performed a more restricted ‘conditional’ enrichment analysis that compared the overlap of network genes to that of other genes whose proteins were also expressed in iNs (i.e., non-interactors detected in our IP-MS experiments, which did not show significant interaction with any of the index proteins; **Supplementary Table 3**). In both the global and conditional analyses, our network showed statistically significant enrichment (*P* < 0.05/17, adjusting for 17 cell types) in several neuronal cell types, including both excitatory neurons and inhibitory interneurons (**Figure 3C**, **Supplementary Figure 8A**, and **Supplementary Table 6**). Next, we tested the global and conditional enrichment of the network in terms of its overlap with cell type-specific DEGs in ASD patients compared to controls. In the global analysis, our network showed significant enrichment in several cell types including excitatory neurons, inhibitory interneurons, astrocytes, and microglia (**Supplementary Figure 8B** and **Supplementary Table 6**). Strikingly, in the more conservative conditional analysis, the layer 2/3 excitatory neurons stand out as the only significant cell type (*P* < 0.05/17; **Figure 3D** and **Supplementary Table 6**). When we further analyzed the up-vs. down-regulated DEGs separately, we found that this enrichment signal is mostly driven by DEGs that are up-regulated in ASD patients (**Supplementary Figure 8C-D** and **Supplementary Table 6**). Taken together, these results firmly establish that the PPI network captures biochemical perturbations and pathways in ASD-relevant tissues and cell types, and specifically implicate layer 2/3 excitatory neurons as a key cell type for ASD pathophysiology.

### Dissecting isoform-specific PPIs in the network: giant ANK2 knockout leads to loss of neuron-specific interactions in maturing neurons

A number of index genes in this study are disrupted in ASDs in an isoform-specific manner. This is the case for giant ankyrin-2 (ANK2) (*39*), a non-canonical and neurospecific alternatively spliced variant of the *ANK2* gene (*40*). This isoform is differentiated from canonical ANK2 due to inclusion of exon 37 (giant exon) encoding 9,240 amino acids, which contains the majority of the mutations identified in ASD patients (**Figure 4A**). When modeled in mice, human ASD-associated mutations of *ANK2* led to gain of axon branching in the central nervous system, a phenotype that is hypothesized to originate from loss of giant ANK2-specific interactions (*39*). While we validated numerous ANK2 interactors identified from the IP-MS experiments performed in iNs (**Figure 4B**, **Supplementary Figure 9A**, and **Supplementary Table 5**), they are likely a mix of interactors of all ANK2 isoforms. Thus, to focus specifically on the ASD-relevant ANK2 isoform we increased the resolution of our interaction data by building an *in vitro* model of selective loss of giant ANK2 using CRISPR/Cas9 gene editing (**Figure 4C**).

**Figure 4.**
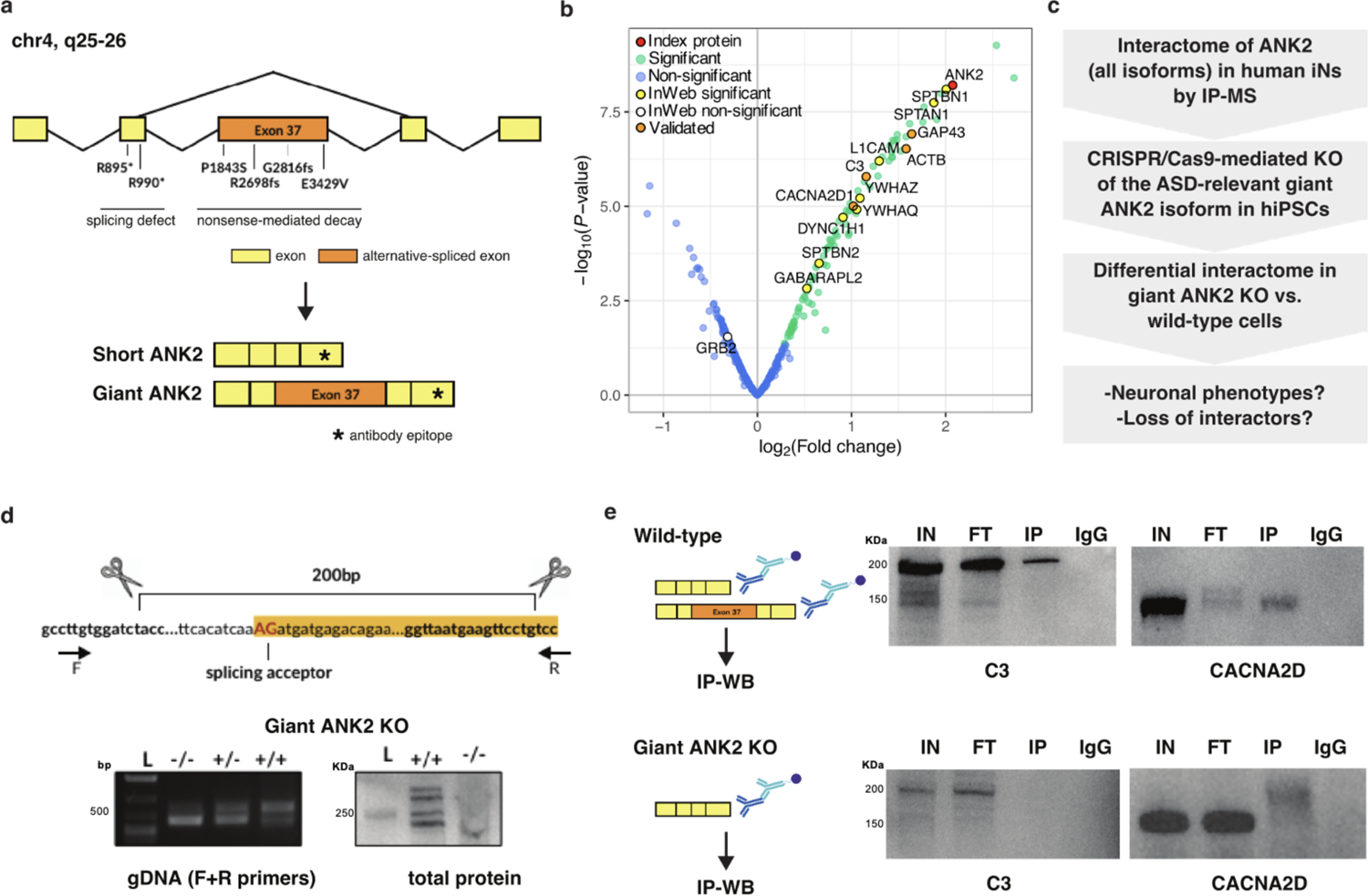
Giant ANK2 is responsible for brain-specific PPIs. **(a)** Depiction of the ANK2 gene with mutations observed in ASD patients leading to splicing defects or early decay. Giant ANK2 includes exon 37, which is alternatively spliced to produce short ANK2. The epitope recognized by the ANK2 antibody used in this study is marked with an asterisk. **(b)** Volcano plot of an IP-MS experiment for ANK2 in iNs, showing ANK2 in red, significant interactors (log_2_ FC > 0 and FDR ≤ 0.1) in green, and non-interactors in blue; known InWeb interactors identified as interactors or non-interactors in the experiment are highlighted in yellow or white, respectively; interactors validated by immunoblotting are highlighted in orange. **(c)** Experimental workflow to identify giant ANK2-specific PPIs. **(d)** Top: CRISPR/Cas9 editing strategy to generate a cell line exclusively expressing the short ANK2 isoform, obtained by deletion of the giant ANK2 exon splicing acceptor. Bottom: validation of giant ANK2 KO cell line. The deletion was confirmed by PCR on genomic DNA using primers flanking the deletion site (left), and by immunoblotting (right) showing the absence of the giant ANK2 isoform (-/-). **(e)** C3 and CACNA2D western blots on ANK2 IPs in wild-type and giant ANK2 KO iNs. L=Ladder, IN=Input, FT=Flow-through, IP=Immunoprecipitation, IgG=IgG control. Molecular weights are in KDa.

We edited an iPS cell line (iPS3, **Materials and Methods**) to exclude retention of exon 37, which differentiates the giant ANK2 from canonical (short) ANK2 isoforms (**Figure 4D**, top). Fully characterized giant ANK2 knockout (KO) iPSCs (**Figure 4D**, bottom) were viable, but showed visible morphological defects upon neuronal differentiation, defective ANK2 distribution, and resulted in high cell death (data not shown). We could not collect sufficient giant ANK2 KO iNs for IP-MS due to high cell death during neuronal maturation. Nonetheless we performed immunoblot analysis in KO iNs to test for presence of selected ANK2 interactors identified by IP-MS in corresponding wild-type (WT) neurons. KO iNs were still expressing proteins essential for various aspects of neuronal physiology, including the calcium channel subunit CACNA2D and the complement protein C3, but these proteins were no longer associated with ANK2 (**Figure 4E**), suggesting that their localization might rely on ANK2-dependent cytoskeletal architecture.

We also performed IP-MS experiments of ANK2 in WT and giant ANK2 KO isogenic neural progenitor cells (NPCs) after one week of neuronal induction, and identified a number of ANK2 interactors that are specific to either WT (which express both canonical and giant ANK2 isoforms) or KO cells (**Supplementary Table 2** and **Supplementary Figure 9B**). GO (*41, 42*) analysis of differential interactors shows that the most enriched cellular component for the WT-specific interactors is “glutamatergic synapses”, while the KO-specific interactors is most enriched for “microtubules” (**Supplementary Table 7**). These results suggest the giant ANK2 isoform plays a central role in localizing the protein to form cell projections of maturating NPCs and in guiding neuronal morphology. This hypothesis is further supported by the observation that ARPC5L is amongst the ANK2-WT-specific interactors. ARPC5L is a component of the ARP2/3 complex that regulates the synthesis of branched actin networks and is essential for both structural and functional maturation of the growth cone in maturating neurons (*43*).

These observations collectively suggest that the wide range of functional defects previously observed in giant ANK2 loss-of-function (LoF) models first result from mis-localization of giant ANK2-specific interactors. Severe defects in cell morphogenesis could be a secondary effect. This vignette demonstrates the potential of using cell type-specific and isoform-specific PPI networks to gain detailed mechanistic insights into ASDs.

### Perturbing a newly found PPI in the network: AKAP8L knockout phenocopies PTEN’s cellular phenotype in iNs

We next performed functional follow-up experiments to investigate a newly identified interaction found in the PTEN IPs. PTEN encodes a phosphatase that acts as a tumor suppressor by antagonizing PI3K/AKT/mTOR signaling (*44*). PTEN is mutated at high frequency in a large number of cancers, but germline mutations of PTEN have also been described in a subset of patients with autism and macrocephaly (*45*). However, it is unclear whether cell proliferation caused by disruption of mTOR signaling upon PTEN mutation can also be linked to these developmental disorders (*46*). To address this question, we further explored the PTEN interactors found by IP-MS in iNs.

Surprisingly, the most robust interactor we identified for PTEN in iNs is the A-kinase anchor protein AKAP8L (**Figure 5A**), which has not been associated with PTEN in previous biochemical studies (that were almost exclusively conducted in cancer cell models or tissues), suggesting a neuron-specific mTOR circuitry of PTEN that acts through AKAP8L. The PTEN-AKAP8L interaction was replicated using two independent PTEN antibodies for IP-MS (data not shown) and consistently validated by immunoblotting and reverse IP (**Figure 5B** and **Supplementary Figure 9C**).

**Figure 5.**
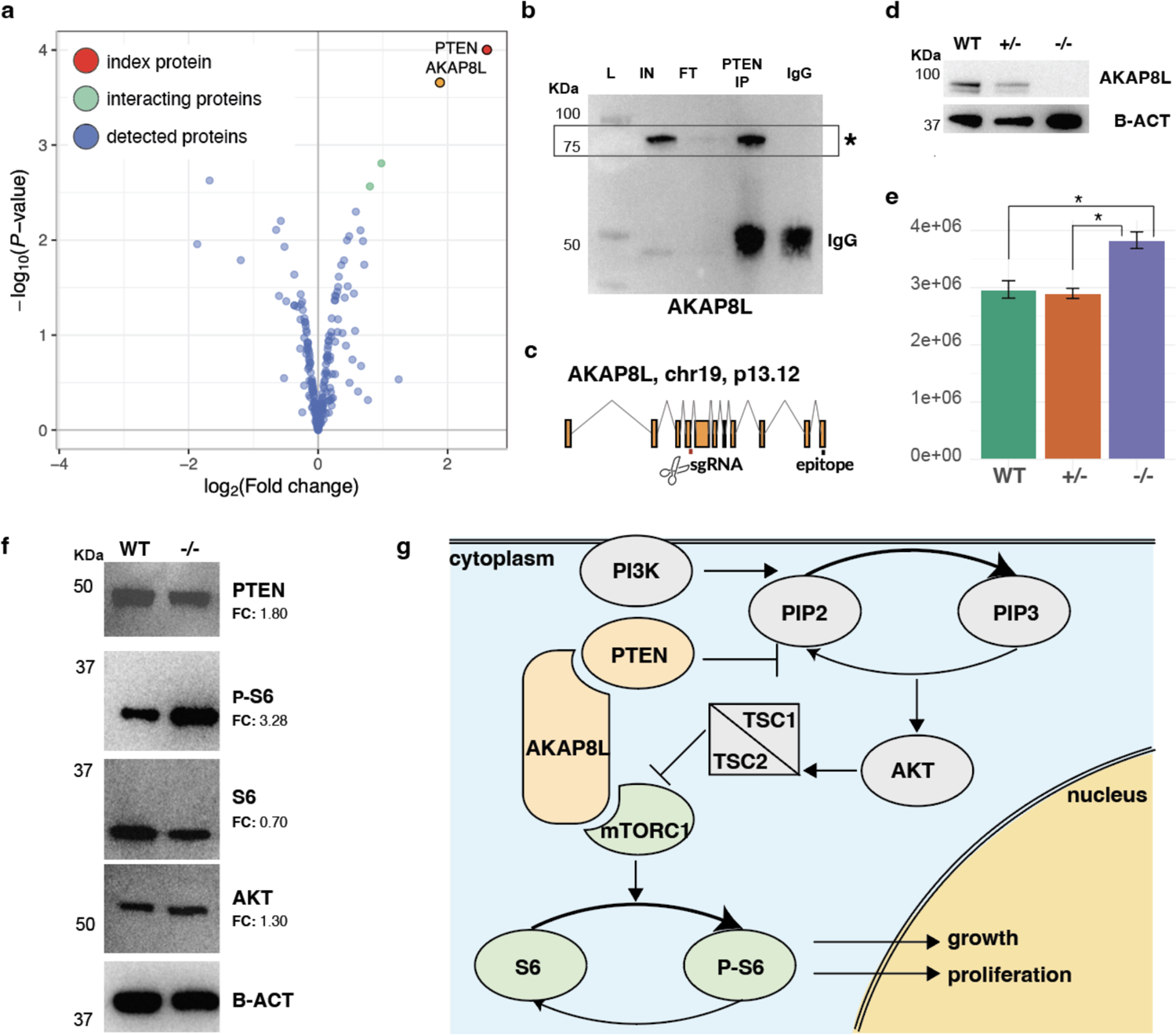
The neuronal PTEN-AKAP8L interaction regulates cell proliferation. (**a)** Volcano plot of an IP-MS experiment for PTEN in iNs, showing PTEN in red, significant interactors (log_2_ FC > 0 and FDR ≤ 0.1) in green, and non-interactors in blue; AKAP8L is marked in orange. **(b)** AKAP8L immunoblot on the PTEN IP. AKAP8L is marked with an asterisk, IgG heavy chains are marked with a vertical line; L=Ladder, IN=Input, FT=Flow-through, IP=Immunoprecipitation, IgG=IgG control. Molecular weights are in KDa. **(c)** Schematic of the CRISPR/Cas9 editing strategy to generate AKAP8L KO iPS cell lines. **(d)** Immunoblot validation of AKAP8L heterozygous (+/-) and homozygous (-/-) KO lines generated using the strategy described in (c). B-actin is used as a loading control. Molecular weights (KDa) are marked on the side of each blot. **(e)** Comparison of wild-type (WT), AKAP8L heterozygous KO (+/-), and AKAP8L homozygous KO (-/-) neural progenitor cell growth over four days upon seeding cells at identical confluences. Error bars are standard deviations of the mean of three biological replicates. P-values were calculated using a two-tailed t-test. **(f)** Immunoblot analysis of PTEN, phosphorylated S6 (P-S6), S6, and AKT in WT and AKAP8L homozygous KO iNs. Molecular weights (KDa) are marked on the side of each blot and B-actin is used as loading control. FC (fold change) was calculated as a ratio of the intensity of the bands detected in the WT vs. AKAP8L KO lanes. **(g)** Pathway depiction of the AKAP8L interaction with both PTEN and mTORC1. Proteins shown in green have been associated with increased cell growth and proliferation while proteins in orange have been linked to decreased cell growth and proliferation.

We generated AKAP8L KO (-/-) iPS cell lines through CRISPR/Cas9 gene editing (**Figure 5C**) to test if they phenocopy neurodevelopmental phenotypes observed in individuals with PTEN-associated autism and macrocephaly. We obtained isogenic lines with both heterozygous (+/-) and homozygous (-/-) AKAP8L mutations, with the homozygous KO line resulting in complete loss of AKAP8L (**Figure 5D**). Interestingly, the AKAP8L homozygous KO line showed a statistically significant increase in growth rate at the NPC stage, compared to isogenic wild-type and heterozygous lines (*P* = 0.0287 and 0.0162, respectively; **Figure 5E**).

To gain deeper mechanistic insights we tested whether this cellular phenotype is linked to a neuron-specific circuitry that includes the mTOR pathway. Although AKAP8L KO did not result in changes of PTEN levels in iNs as measured by immunoblotting, we did observe a small, but detectable increase in the levels of phosphorylated ribosomal protein S6 (**Figure 5F**). This is a marker of hyperactivation of the mTOR pathway and suggests an AKAP8L-mediated defect in the PTEN signaling cascade in ASDs. Interestingly, when we measured the levels of the serine/threonine-protein kinase AKT, we did not find any difference between wild-type and mutant cells, suggesting that AKAP8L acts in the mTORC1 and not in the mTORC2 pathway (*47*). Taken together, our results illustrate a neuronal variation on the PTEN-mTOR circuitry that includes AKAP8L and specifically suggest that the PTEN-AKAP8L interaction impacts downstream mTORC1 signaling and cell proliferation without impacting PTEN protein levels directly (**Figure 5G**).

### Network convergence implicates the IGF2BP1-3 complex in an ASD-relevant transcriptional and protein interaction circuitry

Since the combined PPI network derived from all IP-MS datasets identified a number of interactors linked to multiple index proteins (**Figure 2C,D** and **Supplementary Table 3**), we hypothesized that the degree of convergence in the network could be used to discover biological themes of relevance to ASDs. By ranking proteins in descending order based on the number of index proteins they interact with (**Supplementary Table 3**), we noticed that three insulin-like growth factor 2 mRNA-binding proteins, IGF2BP1, IGF2BP3, and IGF2BP2, are all among the top ten recurring interactors and are associated with seven, six, and five of the index proteins, respectively. All three proteins interact with CHD8, DYRK1A, SCN2A, SHANK3, and SYNGAP1; IGF2BP1 and IGF2BP3 additionally interact with TLK2, and IGF2BP1 further interacts with ANK2.

IGF2BP1-3 belong to a conserved family of single-stranded RNA-binding proteins. Together, they form a N6-methyladenosine (m6A)-reader complex broadly involved in post-transcriptional regulation by impacting mRNA stability and translation (*48*). We therefore set out to test the hypothesis that this complex is implicated in ASDs through a circuit connecting transcription of ASD risk genes to the protein network that we have identified. We analyzed published RNA targets of IGF2BP1-3 identified in HEK cells (*48*) and found that targets of each protein individually, as well as their combined targets, are all significantly enriched for ASD risk genes identified through exome sequencing (*10*) (*P* = 3.9e-10 for the combined target list; **Figure 6A** and **Supplementary Table 8**); among these risk genes are also nine of the index genes in this study.

**Figure 6.**
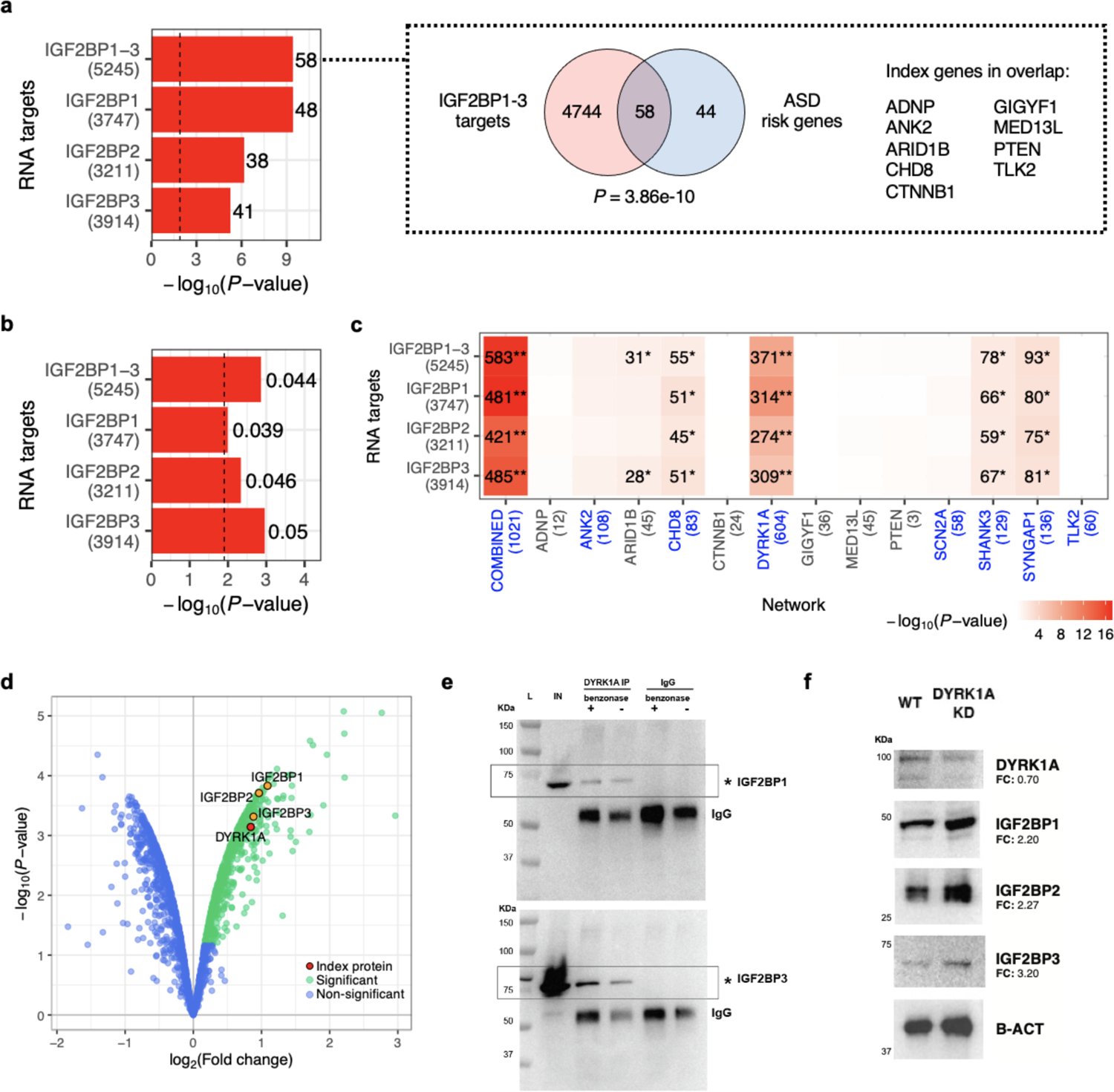
The IGF2BP1-3 complex interacts with ASD-related transcripts and proteins. **(a)** Left: overlap enrichment between RNA targets of IGF2BP1-3 (target counts in parentheses) and ASD risk genes (FDR ≤ 0.1 in exome sequencing data). ‘IGF2BP1-3’ indicates the combined target list (i.e., union of the targets for each IGF2BP). Enrichment P-values were calculated using one-tailed hypergeometric tests; vertical dashed line indicates Bonferroni-corrected (*P* < 0.05/4, adjusting for number of target lists) significance threshold. The number of genes in the overlap is shown to the right of each bar. Right: overlap between the IGF2BP1-3 combined targets and ASD risk genes. Index genes in the overlap are listed on the right. **(b)** Common variant risk enrichment of IGF2BP1-3 targets derived using ASD GWAS data and MAGMA. A vertical dashed line indicates Bonferroni-corrected (*P* < 0.05/4) significance threshold. The enrichment coefficient is shown to the right of each bar. **(c)** Overlap enrichment between IGF2BP1-3 targets and the ASD PPI network or index protein-specific sub-networks (compared to non-interactors). Enrichment P-values were calculated using one-tailed hypergeometric tests; gene counts in overlaps reaching nominal (*P* < 0.05) or Bonferroni (*P* < 0.05/4) significance are shown in the heat map followed by single or double asterisks, respectively. Networks that contain any of the IGF2BPs are highlighted in blue on the x-axis. **(d)** Volcano plot of an IP-MS experiment for DYRK1A in iNs, showing DYRK1A in red, significant interactors (log_2_ FC > 0 and FDR ≤ 0.1) in green, and non-interactors in blue; IGF2BP1-3, which were validated by western blot, are marked in orange. **(e)** Immunoblot on an DYRK1A IP with and without addition of 25U benzonase, as stated at the top. An asterisk marks the expected band of each protein named on the right. L=Ladder, IN=Input, IP=Immunoprecipitation, IgG=IgG control. Molecular weights are in KDa. **(f)** Immunoblot showing protein levels (named on the right) upon DYRK1A siRNA-mediated knockdown for 48 hours in iPSCs. B-actin is used as loading control. FC (fold change) was calculated as a ratio of the intensity of the bands detected in the WT vs. DYRK1A KD lanes. Un=Untreated, KD=knockdown.

Genetic risk enrichment analysis using ASD GWAS data (*49*) and MAGMA (*50*) also found the IGF2BP1-3 targets to be enriched for common variant risk of ASDs (*P* = 1.4e-3 for the combined target list; **Figure 6B** and **Supplementary Table 8**). Furthermore, we performed a conditional enrichment analysis to test whether genes in our combined PPI network and index protein-specific sub-networks are enriched for IGF2BP1-3 targets compared to the non-interactors detected in our neuronal proteomic data (**Figure 6C** and **Supplementary Table 8**). At a Bonferroni-corrected significance threshold (*P* < 0.05/56, adjusting for 14 networks and 4 IGF2BP target lists), the combined and DYRK1A networks both show significant enrichment for IGF2BP1-3 targets (*P* = 1.7e-17 and 7.6e-9 for the combined target list, respectively).

DYRK1A is a ubiquitously expressed protein kinase encoded by a gene localized in the Down syndrome critical region of chromosome 21 and is strongly associated with learning defects associated with Down syndrome (*51*) and ASDs (*52*). DYRK1A has been implicated in post-transcriptional regulation of different cellular processes involved in brain development and function, ranging from early embryogenesis through late aging (*53*). Thus, given the enriched overlap observed between the IGF2BP1-3 targets and the DYRK1A network, we hypothesized that DYRK1A might be part of an ASD-relevant feedback circuitry acting upstream of the IGF2BP1-3 complex. To test this hypothesis, we first verified that the interactions between DYRK1A and IGF2BP1-3 identified by IP-MS (**Figure 6D**) are not mediated by nucleic acids, hence indicating a direct regulatory relationship between DYRK1A and the IGF2BP1-3 complex (**Figure 6E**). Next, we performed siRNA knockdown of DYRK1A in iPSCs, and observed that partial knockdown (30%) was sufficient to trigger increased expression of IGF2BP1-3 (**Figure 6F**). Overall, our results suggest that the IGF2BP1-3 complex regulates an ASD-relevant transcriptional circuitry, and that the newly found interactions between DYRK1A and the complex might be key in modulating the expression of numerous ASD-relevant genes.

### The network is enriched for rare variant risks of ASDs and developmental disorders

To evaluate whether the combined PPI network and index protein-specific sub-networks are enriched for genetic risks of ASDs, we performed global and conditional enrichment analyses using rare variant association statistics derived from exome sequencing of ASD patients and controls (*10*). In the global analysis, we tested whether genes in the networks are more significantly associated with ASDs compared to other protein-coding genes in the genome. In the conditional analysis, we compared the association scores of the network genes against that of the non-interactors detected in our neuronal proteomic data. Five networks (combined, ANK2, CHD8, DYRK1A, SHANK3) reached Bonferroni significance (*P* < 0.05/14, adjusting for 14 networks) in the global analysis (**Supplementary Figure 10A**), and only the combined network was significant (*P* = 1.1e-5) in the conditional analysis (**Figure 7A** and **Supplementary Table 9**). Notably, the conditional enrichment was robust even after we removed known InWeb interactors from the combined network (*P* = 5.5e-5; **Supplementary Figure 10B**), supporting that this signal is not driven by the proteome of the neuronal cell model or the known interaction partners of the index proteins. Rather, the novel, neuron-specific interactions identified in this study are enriched for rare variant risk of ASDs over and above a background set of proteins expressed in iNs. We also performed analogous enrichment analyses using exome sequencing data for developmental disorders (DD) (*54*) and schizophrenia (SCZ) (*55*) and gnomAD (*56*) pLI scores, which represent the probability of a gene being loss-of-function intolerant (**Figure 7A**, **Supplementary Figure 10A,B** and **Supplementary Table 9**). In the conditional analysis, we observed that the combined network is significantly enriched for DD genes (*P* = 6.2e-7) and genes with high pLI scores (*P* = 2.6e-31); five index protein-specific sub-networks (ANK2, ARID1B, CHD8, DYRK1A, SYNGAP1) are also enriched for high pLI scores (*P* = 1.1e-12 to 3.4e-4). In contrast, there was no significant SCZ enrichment for any of the networks. Overall, these results indicate that the interactors that we have identified contribute to rare variant risks of both ASDs and DD more than one would expect for genes generally expressed in neurons. The interactors are also more likely to be intolerant to loss-of-function mutations, suggesting that they might be essential for cell fate specification or survival.

**Figure 7.**
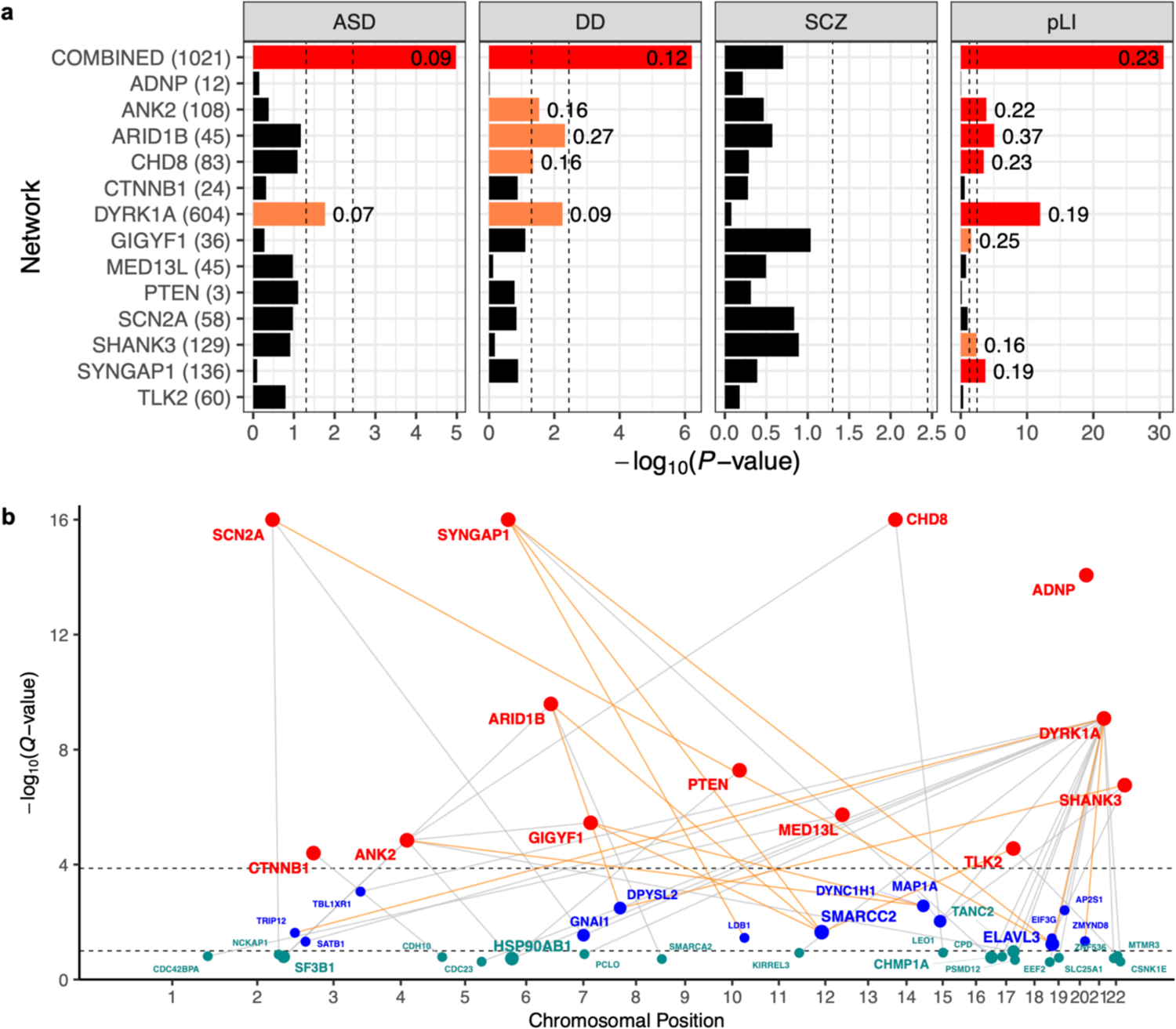
Genetic risk enrichment in the ASD PPI network. (a) Rare variant or pLI score enrichment of the network or index protein-specific sub-networks compared to non-interactors, derived using one-tailed Kolmogorov-Smirnov tests and gene-based association statistics from ASD, DD, or SCZ exome sequencing data or gnomAD pLI scores. The number of genes in each network is shown in parentheses on the y-axis. Left or right vertical dashed lines indicate nominal (*P* < 0.05) or Bonferroni-corrected (*P* < 0.05/14, adjusting for 14 networks) significance thresholds, respectively; nominally or Bonferroni-significant results are highlighted in orange or red, respectively, and labeled with the corresponding KS test statistics. **(b)** Social Manhattan plot of index genes and interactors that are suggestive ASD risk genes in exome sequencing data. Index genes are in red, FDR ≤ 0.1 risk genes are in blue, and FDR ≤ 0.25 risk genes are in cyan; size of the interactor nodes and their labels scale with the number of index genes linked to each interactor. Genes connected by gray lines indicate observed interactions in the ASD PPI network; interactions that have been validated by IP-WB are highlighted in orange.

Next, in order to assess the common variant risk enrichment of the PPI networks, we performed global and conditional analyses using ASD GWAS data (*49*) and MAGMA (*50*). Only the SYNGAP1 network reached nominal significance in both the global and conditional analyses (conditional *P* = 3.71e-3, which is just shy of the Bonferroni-corrected threshold of *P* < 0.05/14; **Supplementary Figure 10C** and **Supplementary Table 10**). We also repeated the analyses using GWAS data of other psychiatric disorders, including attention deficit hyperactivity disorder (ADHD) (*57*), bipolar disorder (BIP) (*58*), major depressive disorder (MDD) (*59*), and SCZ (*60*), as well as height (*61*) as an additional control trait. While several networks were nominally significant for some of these phenotypes, none reached Bonferroni significance (**Supplementary Figure 10C** and **Supplementary Table 10**). Given that the GWAS for ASDs and other psychiatric disorders had relatively modest sample sizes (∼46k to 77k samples, except for MDD which had 500k samples), repeating these analyses when better-powered GWAS become available in the future may allow us to assess whether the suggestive common variant enrichment results reported here reflect true biology related to psychiatric conditions.

### The network can prioritize additional ASD risk genes from genetic studies

Since the combined PPI network is enriched for rare variants associated with ASDs, we used it to prioritize additional ASD risk genes that did not meet stringent statistical significance cutoffs in the exome sequencing study (*10*). Specifically, we generated a ‘social Manhattan plot’ to highlight protein interactions in the network between index genes and other ASD risk genes with FDR ≤ 0.25 in the exome sequencing data (**Figure 7B**). Consistent with the rare variant enrichment results, the overlap between our network and these ASD risk genes is statistically significant (*P* = 4.9e-4), with DYRK1A being linked to the most risk genes out of all the index genes. In total, we prioritized 32 ASD risk genes (13 with FDR ≤ 0.1 and 19 with FDR > 0.1 and ≤ 0.25) that are connected to at least one index gene in the network. Interestingly, these prioritized genes have elevated gene expression throughout cortical development that mirrors the high-confidence, exome-wide significant genes identified from exome sequencing (**Supplementary Figure 10D**). To demonstrate that many of the interactions in the social Manhattan plot are robust and reproducible, we have successfully validated 14 of the interactions using forward and/or reverse IPs followed by western blotting (**Supplementary Table 5**), including multiple interactions involving SMARCC2, ELAVL3, DPYSL2, and DYNC1H1. While follow-up investigation on the prioritized genes is necessary to better validate and evaluate our prioritization approach, our analysis showcases how the ASD PPI network is a unique resource that can be used to corroborate findings from recent GWAS and sequencing studies of ASDs, both in terms of building an exhaustive list of ASD risk genes and generating hypotheses of how these genes might contribute to ASDs mechanistically.

## Discussion

ASDs are a heterogeneous set of heritable neurodevelopmental conditions with contributions from both rare and common genetic variants, and their genetic and biological complexity is reflected in the clinical diversity of ASD patients (*9*). In the past decade, rare variants with large effect sizes in protein-coding genes have been shown to significantly shift individual risk of ASDs during development and early childhood (*11*). In particular, a recent exome sequencing study identified 24 genes that contain an excess of rare deleterious mutations in ASD patients versus controls (*10*). However, how such ASD-associated mutations act singularly or in ensemble remains largely unknown (*14, 15*). For instance, there is evidence that mutations in several of these genes result in similar mild impairments in social and communication skills (*62*). In other cases, specific mutations are associated with altered brain size and other synaptopathies (*63, 64*), but it is still unclear whether their function may converge on a single or restricted number of pathways and networks.

In this study, we combined human genetics, iPSC technology, computational modeling, and interaction proteomics to shed light on how a subset of ASD risk genes (index genes) may functionally converge in disease-relevant and brain cell type-specific pathways. Starting with the 24 high-confidence ASD index genes identified by exome sequencing (*10*), we performed extensive screening of immunoreagents amenable for IP of their encoded proteins, and successfully carried out 26 IP-MS experiments of 13 index proteins (ADNP, ANK2, ARID1B, CHD8, CTNNB1, DYRK1A, GIGYF1, MED13L, PTEN, SCN2A, SHANK3, SYNGAP1, TLK2) in human iPSC-derived excitatory neurons. While excitatory neurons were challenging to obtain *in vitro* in the past (*65*), recent iPSC protocols have enabled large-scale production of homogeneous, electrophysiologically active, and synaptically connected iNs that resemble cortical excitatory neurons in approximately three weeks (*31, 66*), allowing to systematically execute interaction proteomics in an *in vitro* neuronal cell model of key relevance to ASDs according to recent genetic and transcriptomic studies (*67*). We consolidated the iN-derived IP-MS data into a combined PPI network consisting of the 13 index proteins and 1,021 interactor proteins that represents, to our knowledge, the most extensive cell type-specific ASD PPI network thus far. We found that over 90% of the interactions in the network consists of interactions that have not been reported in the literature but are nevertheless highly reproducible, as demonstrated by extensive validation through “forward” and “reverse” immunoprecipitations (91.5% and 82.6% validation rate, respectively). This finding reflects the potential for biological discovery of neuronal-specific interactome proteomics, as opposed to relying on publicly available PPI datasets that are mostly populated by data collected in highly proliferative or immortalized cell types (e.g., HEK cells, cancer cell lines) (*23, 24*). We also compared a subset of the iN-derived PPI data to those derived from human brain homogenates, as well as characterized the co-expression patterns of interacting proteins in iNs across orthogonal brain transcriptomic datasets, showing that a significant portion of the iN-derived PPIs recapitulate interactions and gene relationships observed in complex brain tissues. Furthermore, the lower statistical power in the brain IP-MS data (i.e., lower replicate correlations) supports our choice of executing interaction proteomics in more homogeneous, *in vitro* iNs instead of less abundant and non-renewable brain samples consisting of a complex mix of cell types. The neuronal relevance and specificity of the PPI network is further supported by its enrichment for genes expressed in brain tissues (*37*) and neuronal cell types (*30*). We also observed a significant overlap between the network and differentially expressed genes in the *postmortem* cortex of idiopathic ASD patients versus controls, specifically in layer 2/3 cortical excitatory neurons (*30*). This finding is of particular relevance given that such patients were not genetically characterized, suggesting that PPIs can be envisioned as a tool to stratify ASD patients in the future.

Our findings also illustrate how novel, brain-specific interactions within the PPI network can link ASD-associated genetic signals to relevant neuronal phenotypes. The *ANK2* gene, for instance, encodes for two major protein isoforms as a result of alternative splicing. One protein isoform has a molecular weight of 220 kDa and is expressed in multiple tissues, while the other (commonly referred to as giant ANK2) has a molecular weight of 440 kDa due to the inclusion of exon 37 and is expressed only in the nervous system (*40*). Three human ANK2 ASD mutations (P1843S, R2608 frameshift, and E3429V) are located in exon 37 and thus only affect giant ANK2, whereas others (R895*, R990*, Q3683fs) affect both isoforms (*68*). In particular, a mouse model for human ASD mutation of giant ANK2 shows increased axonal branching and ectopic connectivity in cultured neurons, as well as a transient increase in excitatory synapses during postnatal development (*39*). In order to identify isoform-specific PPIs that may contribute to ANK2 function in axon guidance, we designed a giant ANK2 KO iPSC model and compared ANK2 IPs performed in the wild-type versus giant ANK2 KO NPCs and iNs. Our results suggest that giant ANK2 and its PPIs are essential for proper localization of cytoskeletal components to cell projections and maintenance of neuronal morphology. We were also able to recapitulate cellular phenotypes observed in mice and concluded that they are the result of both primary loss-of-function of neuronal proteins (likely due to mis-localization), and secondary effects linked to severely impaired cell morphogenesis. This observation is particularly relevant in the light of recent efforts of utilizing synthetic oligonucleotides to either inhibit or restore specific transcripts (*69*).

Another example of how the PPI network can be used to reconcile genetic signals with specific molecular mechanisms leading to cellular phenotypes resides in the newly characterized interaction between PTEN and AKAP8L. *PTEN* is frequently mutated in cancers due to its antagonizing role in PI3K/AKT/mTOR signaling (*44*), but germline mutations of *PTEN* have also been described in a subset of patients with autism combined with macrocephaly (*45*). However, there is no consensus on whether PTEN-mediated cell proliferation defects are tied to the neuronal phenotypes observed in ASDs. We found that knockout of the pseudo-kinase AKAP8L, the most robust interaction partner of PTEN in iNs, phenocopies its cellular phenotype. This discovery in iNs, together with the recently reported interaction between AKAP8L and mTORC1 (*70*), indicate that AKAP8L could be implicated in ASDs by bridging PTEN to key players in the mTOR pathway in human neurons. A previous study of a family with autism and macrocephaly has also identified AKAP8L as a likely candidate gene affected by the disease-associated genomic alteration at 19p13.12 (*71*), further supporting the hypothesis that the PTEN-AKAP8L interaction may influence neurodevelopmental phenotypes through the mTOR signaling cascades. This observation also offers the opportunity to explore AKAP8L as a potential target for therapeutic interventions in the mTOR pathway.

Our approach of generating a combined PPI network of multiple ASD index proteins also allowed us to experimentally identify functional convergence of these proteins based on their shared interactions (*16, 18, 72*). The hypothesis that ASD risk genes may act in a small number of networks has been long debated (*15, 73*), and previous studies have only identified broad functional categories (e.g., “gene expression regulation”, “neuronal communication”, and “cytoskeleton”) to be enriched among known ASD risk genes (*10*). In this study, we identified a number of proteins involved in RNA metabolism as being highly interconnected to multiple index proteins. Intriguingly, we found three components of the same complex (i.e., IGF2BP1, IGF2BP2, and IGF2BP3) to be among the most recurrent proteins in the network, each interacting with 5-7 index proteins. Insulin-like growth factor 2 mRNA-binding proteins 1-3 (IGF2BP1-3) were recently characterized as part of a N6-methyladenosine (m6A)-reader complex broadly involved in post-transcriptional regulation by impacting mRNA stability and translation rates (*48*). Numerous studies have shown that mRNA transport, turnover, and translation of synaptic proteins are key to maintaining neuronal health, synaptic plasticity, learning, and memory (*74*), and m6A deposition has emerged as a critical regulatory mechanism for these functions in neurons and neural progenitors (*75–78*). Growing evidence has also linked FMRP (Fragile X mental retardation protein, named after the most common single-gene disorder associated with ASDs) and its role in synaptic protein regulation to its binding of m6A-modified transcripts (*79*). Although the IGF2BP1-3 complex has a well-documented role in modulating neural stem cell growth and division in *Drosophila* (*80*), there is no significant genetic evidence linking them to ASDs. However, in light of our findings, we hypothesize that similar to FMRP, the complex may be broadly involved in the regulation of ASD-relevant genes, and that its lack of genetic signals in ASDs may be due to the fact that at least two components of the complex, IGF2BP1 and IGF2BP3, are intolerant to LoF mutations in humans (pLI scores = 0.999, 0.239, and 1 for IGF2BP1-3, respectively) (*56*). To explore this hypothesis, we surveyed exome sequencing and GWAS data for ASDs and related disorders and found suggestive evidence (nominal *P* < 0.05) linking IGF2BP1 and IGF2BP3 to developmental disorders, schizophrenia, and/or ADHD (**Supplementary Table 11**), although larger genetic datasets will be needed to confirm if these are true associations. Furthermore, we analyzed published RNA targets of IGF2BP1-3 (*48*) and found that they are significantly enriched for ASD risk genes identified through exome sequencing, common variant risks of ASDs, as well as genes in our PPI network. In particular, we found the DYRK1A interactome to be strongly enriched for IGF2BP1-3 targets, and showed that partial knockdown of DYRK1A led to increased expression of IGF2BP1-3. All together, these findings suggest that the IGF2BP1-3 complex and DYRK1A might be part of a feedback mechanism reinforcing the expression of ASD-relevant transcripts, and offers opportunity for therapeutic strategies aimed at restoring IGF2BP1-3 levels upon modulation of DYRK1A (e.g., through the use of commercially available DYRK1A inhibitors (*81*)), or *vice versa*, employing antisense oligonucleotides to target the IGF2BP1-3 complex. Importantly, these findings also showcase how convergent signals in the PPI network can be used to identify ASD-relevant genes and pathways that large-scale genetic studies have yet to discover due to either early embryonic lethality or sampling/statistical power limitations.

Finally, we show that the network is indeed statistically enriched for rare variant risks of ASDs and developmental disorders captured in exome sequencing data (*10, 54*). Interestingly, the enrichment appears to be specific to these developmental phenotypes, as the same degree of enrichment was not observed for SCZ (*55*). Given these results, we integrated the network with ASD exome sequencing data in a ‘social Manhattan plot’ to demonstrate that it can be used to prioritize risk genes with suggestive significance in the genetic analysis, based on their interactions with ASD index proteins in the network. Although we did not observe an enrichment for common variant risk of ASDs in the network, it is possible that this is due to statistical power limitations of the most recent ASD GWAS; we expect that with more data in the future, our network will also enable the prioritization of risk genes from GWAS data. This expectation is supported by our findings in a parallel study (co-submitted manuscript), where we applied the same brain cell type-specific interaction proteomics framework to generate an iN-derived PPI network for SCZ risk genes prioritized from GWAS loci. As well-powered, multi-ancestry GWAS datasets are available for SCZ, we found that the SCZ PPI network is enriched for common variant risk of SCZ in both Europeans and East Asians, and that the network can complement conventional strategies such as statistical fine-mapping and eQTL co-localization analyses to prioritize additional risk genes from GWAS loci. Repeating similar analyses for ASDs when larger GWAS datasets become available in the future may allow us to better determine if rare and common variant risks of ASDs generally converge onto shared pathways represented in the ASD PPI network, as we have already seen specifically for the transcriptional circuit regulated by IGF2BP1-3.

In conclusion, we built a PPI network of 13 index proteins encoded by high-confidence ASD risk genes in human iPSC-derived excitatory neurons. This network, which contains > 90% newly characterized interactions, is enriched for genes expressed in brain tissues and neuronal cell types, transcriptionally perturbed genes in the brains of idiopathic ASD patients, as well as genes containing rare deleterious mutations associated with ASDs and developmental disorders. We performed follow-up experiments and analyses to demonstrate that the network contains rich information for uncovering ASD-related biology, both in terms of investigating the individual PPIs (e.g., giant ANK2-specific PPIs and PTEN-AKAP8L interaction) and the convergent signals (e.g., IGF2BP1-3 complex) in the network. Overall, our findings not only bring clarity on how certain risk genes contribute to ASDs, but also suggest convergent biology and shared pathways that may inform therapeutic intervention or patient stratification strategies. Going forward, the approach described in this study and the parallel SCZ study (co-submitted manuscript) may be adapted and expanded to generate PPI networks for additional disease risk genes in various brain cell types in order to compile a comprehensive cell type-specific PPI atlas for psychiatric disorders.

## Data Availability

Raw RNA-seq and IP-MS data will be available through GEO and MassIVE, respectively, upon publication of the manuscript.

## Acknowledgements

We would like to thank Hailiang Huang and Ruize Liu for fruitful discussions and analytical advice, and Steve Hyman for helping conceptualize this study and critically reading the manuscript. We thank the Broad Proteomics Platform, Whitehead Proteomics Core Facility, and Harvard Center for Mass Spectrometry for discussions and generation of IP-MS data.

## Author Contributions

GP and JR carried out tissue culture. GP, JMM, JR, and JCB tested antibodies and ran immunoblots. GP and KWL executed IP experiments. KWL and MAG ran MS experiments and analysis at CNCR. GP performed the rest of the experiments. KT performed bulk RNA-seq, IP-MS, GTEx, and BrainSpan analyses. JKTC performed comparisons of IP-MS data. YHH performed bulk RNA-seq, IP-MS, BrainSpan, and the rest of the analyses. ABS, NF, KCE, and KL supervised and managed the study. KCE and KL designed the study. KL initiated and led the study. GP, YHH, KT, NF, KCE, and KL wrote the manuscript with input from co-authors.

## References

1. E. B. Robinson, B. M. Neale, S. E. Hyman, Genetic research in autism spectrum disorders. Curr. Opin. Pediatr. 27, 685–91 (2015).

2. B. S. Abrahams, D. H. Geschwind, Advances in autism genetics: on the threshold of a new neurobiology. Nat. Rev. Genet. 9, 341–55 (2008).

3. T. Gaugler, L. Klei, S. J. Sanders, C. A. Bodea, A. P. Goldberg, A. B. Lee, M. Mahajan, D. Manaa, Y. Pawitan, J. Reichert, S. Ripke, S. Sandin, P. Sklar, O. Svantesson, A. Reichenberg, C. M. Hultman, B. Devlin, K. Roeder, J. D. Buxbaum, Most genetic risk for autism resides with common variation. Nat. Genet. 46, 881–5 (2014).

4. I. Iossifov, B. J. O’Roak, S. J. Sanders, M. Ronemus, N. Krumm, D. Levy, H. A. Stessman, K. T. Witherspoon, L. Vives, K. E. Patterson, J. D. Smith, B. Paeper, D. A. Nickerson, J. Dea, S. Dong, L. E. Gonzalez, J. D. Mandell, S. M. Mane, M. T. Murtha, C. A. Sullivan, M. F. Walker, Z. Waqar, L. Wei, A. J. Willsey, B. Yamrom, Y. Lee, E. Grabowska, E. Dalkic, Z. Wang, S. Marks, P. Andrews, A. Leotta, J. Kendall, I. Hakker, J. Rosenbaum, B. Ma, L. Rodgers, J. Troge, G. Narzisi, S. Yoon, M. C. Schatz, K. Ye, W. R. McCombie, J. Shendure, E. E. Eichler, M. W. State, M. Wigler, The contribution of de novo coding mutations to autism spectrum disorder. Nature. 515, 216–21 (2014).

5. J. Sebat, B. Lakshmi, D. Malhotra, J. Troge, C. Lese-Martin, T. Walsh, B. Yamrom, S. Yoon, A. Krasnitz, J. Kendall, A. Leotta, D. Pai, R. Zhang, Y.-H. Lee, J. Hicks, S. J. Spence, A. T. Lee, K. Puura, T. Lehtimäki, D. Ledbetter, P. K. Gregersen, J. Bregman, J. S. Sutcliffe, V. Jobanputra, W. Chung, D. Warburton, M.-C. King, D. Skuse, D. H. Geschwind, T. C. Gilliam, K. Ye, M. Wigler, Strong association of de novo copy number mutations with autism. Science. 316, 445–9 (2007).

6. S. J. Sanders, X. He, A. J. Willsey, A. G. Ercan-Sencicek, K. E. Samocha, A. E. Cicek, M. T. Murtha, V. H. Bal, S. L. Bishop, S. Dong, A. P. Goldberg, C. Jinlu, J. F. Keaney, L. Klei, J. D. Mandell, D. Moreno-De-Luca, C. S. Poultney, E. B. Robinson, L. Smith, T. Solli-Nowlan, M. Y. Su, N. A. Teran, M. F. Walker, D. M. Werling, A. L. Beaudet, R. M. Cantor, E. Fombonne, D. H. Geschwind, D. E. Grice, C. Lord, J. K. Lowe, S. M. Mane, D. M. Martin, E. M. Morrow, M. E. Talkowski, J. S. Sutcliffe, C. A. Walsh, T. W. Yu, Autism Sequencing Consortium, D. H. Ledbetter, C. L. Martin, E. H. Cook, J. D. Buxbaum, M. J. Daly, B. Devlin, K. Roeder, M. W. State, Insights into Autism Spectrum Disorder Genomic Architecture and Biology from 71 Risk Loci. Neuron. 87, 1215–1233 (2015).

7. D. J. Weiner, E. M. Wigdor, S. Ripke, R. K. Walters, J. A. Kosmicki, J. Grove, K. E. Samocha, J. I. Goldstein, A. Okbay, J. Bybjerg-Grauholm, T. Werge, D. M. Hougaard, J. Taylor, iPSYCH-Broad Autism Group, Psychiatric Genomics Consortium Autism Group, D. Skuse, B. Devlin, R. Anney, S. J. Sanders, S. Bishop, P. B. Mortensen, A. D. Børglum, G. D. Smith, M. J. Daly, E. B. Robinson, Polygenic transmission disequilibrium confirms that common and rare variation act additively to create risk for autism spectrum disorders. Nat. Genet. 49, 978–985 (2017).

8. E. B. Robinson, B. St Pourcain, V. Anttila, J. A. Kosmicki, B. Bulik-Sullivan, J. Grove, J. Maller, K. E. Samocha, S. J. Sanders, S. Ripke, J. Martin, M. V Hollegaard, T. Werge, D. M. Hougaard, iPSYCH-SSI-Broad Autism Group, B. M. Neale, D. M. Evans, D. Skuse, P. B. Mortensen, A. D. Børglum, A. Ronald, G. D. Smith, M. J. Daly, Genetic risk for autism spectrum disorders and neuropsychiatric variation in the general population. Nat. Genet. 48, 552–5 (2016).

9. D. H. Geschwind, Genetics of autism spectrum disorders. Trends Cogn. Sci. 15, 409–16 (2011).

10. F. K. Satterstrom, J. A. Kosmicki, J. Wang, M. S. Breen, S. De Rubeis, J. Y. An, M. Peng, R. Collins, J. Grove, L. Klei, C. Stevens, J. Reichert, M. S. Mulhern, M. Artomov, S. Gerges, B. Sheppard, X. Xu, A. Bhaduri, U. Norman, H. Brand, G. Schwartz, R. Nguyen, E. E. Guerrero, C. Dias, B. Aleksic, R. Anney, M. Barbosa, S. Bishop, A. Brusco, J. Bybjerg-Grauholm, A. Carracedo, M. C. Y. Chan, A. G. Chiocchetti, B. H. Y. Chung, H. Coon, M. L. Cuccaro, A. Curró, B. Dalla Bernardina, R. Doan, E. Domenici, S. Dong, C. Fallerini, M. Fernández-Prieto, G. B. Ferrero, C. M. Freitag, M. Fromer, J. J. Gargus, D. Geschwind, E. Giorgio, J. González-Peñas, S. Guter, D. Halpern, E. Hansen-Kiss, X. He, G. E. Herman, I. Hertz-Picciotto, D. M. Hougaard, C. M. Hultman, I. Ionita-Laza, S. Jacob, J. Jamison, A. Jugessur, M. Kaartinen, G. P. Knudsen, A. Kolevzon, I. Kushima, S. L. Lee, T. Lehtimäki, E. T. Lim, C. Lintas, W. I. Lipkin, D. Lopergolo, F. Lopes, Y. Ludena, P. Maciel, P. Magnus, B. Mahjani, N. Maltman, D. S. Manoach, G. Meiri, I. Menashe, J. Miller, N. Minshew, E. M. S. Montenegro, D. Moreira, E. M. Morrow, O. Mors, P. B. Mortensen, M. Mosconi, P. Muglia, B. M. Neale, M. Nordentoft, N. Ozaki, A. Palotie, M. Parellada, M. R. Passos-Bueno, M. Pericak-Vance, A. M. Persico, I. Pessah, K. Puura, A. Reichenberg, A. Renieri, E. Riberi, E. B. Robinson, K. E. Samocha, S. Sandin, S. L. Santangelo, G. Schellenberg, S. W. Scherer, S. Schlitt, R. Schmidt, L. Schmitt, I. M. W. Silva, T. Singh, P. M. Siper, M. Smith, G. Soares, C. Stoltenberg, P. Suren, E. Susser, J. Sweeney, P. Szatmari, L. Tang, F. Tassone, K. Teufel, E. Trabetti, M. P. del Trelles, C. A. Walsh, L. A. Weiss, T. Werge, D. M. Werling, E. M. Wigdor, E. Wilkinson, A. J. Willsey, T. W. Yu, M. H. C. Yu, R. Yuen, E. Zachi, E. Agerbo, T. D. Als, V. Appadurai, M. Bækvad-Hansen, R. Belliveau, A. Buil, C. E. Carey, F. Cerrato, K. Chambert, C. Churchhouse, S. Dalsgaard, D. Demontis, A. Dumont, J. Goldstein, C. S. Hansen, M. E. Hauberg, M. V. Hollegaard, D. P. Howrigan, H. Huang, J. Maller, A. R. Martin, J. Martin, M. Mattheisen, J. Moran, J. Pallesen, D. S. Palmer, C. B. Pedersen, M. G. Pedersen, T. Poterba, J. B. Poulsen, S. Ripke, A. J. Schork, W. K. Thompson, P. Turley, R. K. Walters, C. Betancur, E. H. Cook, L. Gallagher, M. Gill, J. S. Sutcliffe, A. Thurm, M. E. Zwick, A. D. Børglum, M. W. State, A. E. Cicek, M. E. Talkowski, D. J. Cutler, B. Devlin, S. J. Sanders, K. Roeder, M. J. Daly, J. D. Buxbaum, Large-Scale Exome Sequencing Study Implicates Both Developmental and Functional Changes in the Neurobiology of Autism. Cell. 180, 568–584.e23 (2020).

11. X. Jin, S. K. Simmons, A. Guo, A. S. Shetty, M. Ko, L. Nguyen, V. Jokhi, E. Robinson, P. Oyler, N. Curry, G. Deangeli, S. Lodato, J. Z. Levin, A. Regev, F. Zhang, P. Arlotta, In vivo Perturb-Seq reveals neuronal and glial abnormalities associated with autism risk genes. Science. 370 (2020), doi:10.1126/science.aaz6063.

12. S. De Rubeis, X. He, A. P. Goldberg, C. S. Poultney, K. Samocha, A. E. Cicek, Y. Kou, L. Liu, M. Fromer, S. Walker, T. Singh, L. Klei, J. Kosmicki, F. Shih-Chen, B. Aleksic, M. Biscaldi, P. F. Bolton, J. M. Brownfeld, J. Cai, N. G. Campbell, A. Carracedo, M. H. Chahrour, A. G. Chiocchetti, H. Coon, E. L. Crawford, S. R. Curran, G. Dawson, E. Duketis, B. A. Fernandez, L. Gallagher, E. Geller, S. J. Guter, R. S. Hill, J. Ionita-Laza, P. Jimenz Gonzalez, H. Kilpinen, S. M. Klauck, A. Kolevzon, I. Lee, I. Lei, J. Lei, T. Lehtimäki, C.-F. Lin, A. Ma’ayan, C. R. Marshall, A. L. McInnes, B. Neale, M. J. Owen, N. Ozaki, M. Parellada, J. R. Parr, S. Purcell, K. Puura, D. Rajagopalan, K. Rehnström, A. Reichenberg, A. Sabo, M. Sachse, S. J. Sanders, C. Schafer, M. Schulte-Rüther, D. Skuse, C. Stevens, P. Szatmari, K. Tammimies, O. Valladares, A. Voran, W. Li-San, L. A. Weiss, A. J. Willsey, T. W. Yu, R. K. C. Yuen, DDD Study, Homozygosity Mapping Collaborative for Autism, UK10K Consortium, E. H. Cook, C. M. Freitag, M. Gill, C. M. Hultman, T. Lehner, A. Palotie, G. D. Schellenberg, P. Sklar, M. W. State, J. S. Sutcliffe, C. A. Walsh, S. W. Scherer, M. E. Zwick, J. C. Barett, D. J. Cutler, K. Roeder, B. Devlin, M. J. Daly, J. D. Buxbaum, Synaptic, transcriptional and chromatin genes disrupted in autism. Nature. 515, 209–15 (2014).

13. D. Pinto, E. Delaby, D. Merico, M. Barbosa, A. Merikangas, L. Klei, B. Thiruvahindrapuram, X. Xu, R. Ziman, Z. Wang, J. A. S. Vorstman, A. Thompson, R. Regan, M. Pilorge, G. Pellecchia, A. T. Pagnamenta, B. Oliveira, C. R. Marshall, T. R. Magalhaes, J. K. Lowe, J. L. Howe, A. J. Griswold, J. Gilbert, E. Duketis, B. A. Dombroski, M. V De Jonge, M. Cuccaro, E. L. Crawford, C. T. Correia, J. Conroy, I. C. Conceição, A. G. Chiocchetti, J. P. Casey, G. Cai, C. Cabrol, N. Bolshakova, E. Bacchelli, R. Anney, S. Gallinger, M. Cotterchio, G. Casey, L. Zwaigenbaum, K. Wittemeyer, K. Wing, S. Wallace, H. van Engeland, A. Tryfon, S. Thomson, L. Soorya, B. Rogé, W. Roberts, F. Poustka, S. Mouga, N. Minshew, L. A. McInnes, S. G. McGrew, C. Lord, M. Leboyer, A. S. Le Couteur, A. Kolevzon, P. Jiménez González, S. Jacob, R. Holt, S. Guter, J. Green, A. Green, C. Gillberg, B. A. Fernandez, F. Duque, R. Delorme, G. Dawson, P. Chaste, C. Café, S. Brennan, T. Bourgeron, P. F. Bolton, S. Bölte, R. Bernier, G. Baird, A. J. Bailey, E. Anagnostou, J. Almeida, E. M. Wijsman, V. J. Vieland, A. M. Vicente, G. D. Schellenberg, M. Pericak-Vance, A. D. Paterson, J. R. Parr, G. Oliveira, J. I. Nurnberger, A. P. Monaco, E. Maestrini, S. M. Klauck, H. Hakonarson, J. L. Haines, D. H. Geschwind, C. M. Freitag, S. E. Folstein, S. Ennis, H. Coon, A. Battaglia, P. Szatmari, J. S. Sutcliffe, J. Hallmayer, M. Gill, E. H. Cook, J. D. Buxbaum, B. Devlin, L. Gallagher, C. Betancur, S. W. Scherer, Convergence of genes and cellular pathways dysregulated in autism spectrum disorders. Am. J. Hum. Genet. 94, 677–94 (2014).

14. D. H. Geschwind, Autism: many genes, common pathways? Cell. 135, 391–5 (2008).

15. J. M. Berg, D. H. Geschwind, Autism genetics: searching for specificity and convergence. Genome Biol. 13, 247 (2012).

16. N. Safari-Alighiarloo, M. Taghizadeh, M. Rezaei-Tavirani, B. Goliaei, A. A. Peyvandi, Protein-protein interaction networks (PPI) and complex diseases. Gastroenterol. Hepatol. from bed to bench. 7, 17–31 (2014).

17. S. Choobdar, M. E. Ahsen, J. Crawford, M. Tomasoni, T. Fang, D. Lamparter, J. Lin, B. Hescott, X. Hu, J. Mercer, T. Natoli, R. Narayan, DREAM Module Identification Challenge Consortium, A. Subramanian, J. D. Zhang, G. Stolovitzky, Z. Kutalik, K. Lage, D. K. Slonim, J. Saez-Rodriguez, L. J. Cowen, S. Bergmann, D. Marbach, Assessment of network module identification across complex diseases. Nat. Methods. 16, 843–852 (2019).

18. K. Lage, Protein-protein interactions and genetic diseases: The interactome. Biochim. Biophys. Acta-Mol. Basis Dis. 1842, 1971–1980 (2014).

19. A. Lundby, E. J. Rossin, A. B. Steffensen, M. R. Acha, C. Newton-Cheh, A. Pfeufer, S. N. Lynch, QT Interval International GWAS Consortium (QT-IGC), S.-P. Olesen, S. Brunak, P. T. Ellinor, J. W. Jukema, S. Trompet, I. Ford, P. W. Macfarlane, B. P. Krijthe, A. Hofman, A. G. Uitterlinden, B. H. Stricker, H. M. Nathoe, W. Spiering, M. J. Daly, F. W. Asselbergs, P. van der Harst, D. J. Milan, P. I. W. de Bakker, K. Lage, J. V Olsen, Annotation of loci from genome-wide association studies using tissue-specific quantitative interaction proteomics. Nat. Methods. 11, 868–74 (2014).

20. B. J. O’Roak, L. Vives, S. Girirajan, E. Karakoc, N. Krumm, B. P. Coe, R. Levy, A. Ko, C. Lee, J. D. Smith, E. H. Turner, I. B. Stanaway, B. Vernot, M. Malig, C. Baker, B. Reilly, J. M. Akey, E. Borenstein, M. J. Rieder, D. A. Nickerson, R. Bernier, J. Shendure, E. E. Eichler, Sporadic autism exomes reveal a highly interconnected protein network of de novo mutations. Nature. 485, 246–50 (2012).

21. B. M. Neale, Y. Kou, L. Liu, A. Ma’ayan, K. E. Samocha, A. Sabo, C.-F. Lin, C. Stevens, L.-S. Wang, V. Makarov, P. Polak, S. Yoon, J. Maguire, E. L. Crawford, N. G. Campbell, E. T. Geller, O. Valladares, C. Schafer, H. Liu, T. Zhao, G. Cai, J. Lihm, R. Dannenfelser, O. Jabado, Z. Peralta, U. Nagaswamy, D. Muzny, J. G. Reid, I. Newsham, Y. Wu, L. Lewis, Y. Han, B. F. Voight, E. Lim, E. Rossin, A. Kirby, J. Flannick, M. Fromer, K. Shakir, T. Fennell, K. Garimella, E. Banks, R. Poplin, S. Gabriel, M. DePristo, J. R. Wimbish, B. E. Boone, S. E. Levy, C. Betancur, S. Sunyaev, E. Boerwinkle, J. D. Buxbaum, E. H. Cook Jr, B. Devlin, R. a. Gibbs, K. Roeder, G. D. Schellenberg, J. S. Sutcliffe, M. J. Daly, Patterns and rates of exonic de novo mutations in autism spectrum disorders. Nature. 485, 242–245 (2012).

22. T. Li, A. Kim, J. Rosenbluh, H. Horn, L. Greenfeld, D. An, A. Zimmer, A. Liberzon, J. Bistline, T. Natoli, Y. Li, A. Tsherniak, R. Narayan, A. Subramanian, T. Liefeld, B. Wong, D. Thompson, S. Calvo, S. Carr, J. Boehm, J. Jaffe, J. Mesirov, N. Hacohen, A. Regev, K. Lage, GeNets: a unified web platform for network-based genomic analyses. Nat. Methods. 15, 543–546 (2018).

23. T. Li, R. Wernersson, R. B. Hansen, H. Horn, J. Mercer, G. Slodkowicz, C. T. Workman, O. Rigina, K. Rapacki, H. H. Stærfeldt, S. Brunak, T. S. Jensen, K. Lage, A scored human protein-protein interaction network to catalyze genomic interpretation. Nat. Methods. 14, 61–64 (2016).

24. G. Pintacuda, F. H. Lassen, Y.-H. H. Hsu, A. Kim, J. M. Martín, E. Malolepsza, J. K. Lim, N. Fornelos, K. C. Eggan, K. Lage, Genoppi is an open-source software for robust and standardized integration of proteomic and genetic data. Nat. Commun. 12, 2580 (2021).

25. N. N. Parikshak, R. Luo, A. Zhang, H. Won, J. K. Lowe, V. Chandran, S. Horvath, D. H. Geschwind, Integrative functional genomic analyses implicate specific molecular pathways and circuits in autism. Cell. 155, 1008–21 (2013).

26. C. L. Hartl, G. Ramaswami, W. G. Pembroke, S. Muller, G. Pintacuda, A. Saha, P. Parsana, A. Battle, K. Lage, D. H. Geschwind, Coexpression network architecture reveals the brain-wide and multiregional basis of disease susceptibility. Nat. Neurosci. 24, 1313– 1323 (2021).

27. X. Xu, A. B. Wells, D. R. O’Brien, A. Nehorai, J. D. Dougherty, Cell type-specific expression analysis to identify putative cellular mechanisms for neurogenetic disorders. J. Neurosci. 34, 1420–31 (2014).

28. I. Voineagu, X. Wang, P. Johnston, J. K. Lowe, Y. Tian, S. Horvath, J. Mill, R. M. Cantor, B. J. Blencowe, D. H. Geschwind, Transcriptomic analysis of autistic brain reveals convergent molecular pathology. Nature. 474, 380–4 (2011).

29. G. Ramaswami, H. Won, M. J. Gandal, J. Haney, J. C. Wang, C. C. Y. Wong, W. Sun, S. Prabhakar, J. Mill, D. H. Geschwind, Integrative genomics identifies a convergent molecular subtype that links epigenomic with transcriptomic differences in autism. Nat. Commun. 11, 4873 (2020).

30. D. Velmeshev, L. Schirmer, D. Jung, M. Haeussler, Y. Perez, S. Mayer, A. Bhaduri, N. Goyal, D. H. Rowitch, A. R. Kriegstein, Single-cell genomics identifies cell type–specific molecular changes in autism. Science (80-.). 364, 685–689 (2019).

31. R. Nehme, E. Zuccaro, S. D. Ghosh, C. Li, J. L. Sherwood, O. Pietilainen, L. E. Barrett, F. Limone, K. A. Worringer, S. Kommineni, Y. Zang, D. Cacchiarelli, A. Meissner, R. Adolfsson, S. Haggarty, J. Madison, M. Muller, P. Arlotta, Z. Fu, G. Feng, K. Eggan, Combining NGN2 Programming with Developmental Patterning Generates Human Excitatory Neurons with NMDAR-Mediated Synaptic Transmission. Cell Rep. 23, 2509– 2523 (2018).

32. Y. Zhang, C. Pak, Y. Han, H. Ahlenius, Z. Zhang, S. Chanda, S. Marro, C. Patzke, C. Acuna, J. Covy, W. Xu, N. Yang, T. Danko, L. Chen, M. Wernig, T. C. Südhof, Rapid single-step induction of functional neurons from human pluripotent stem cells. Neuron. 78, 785–98 (2013).

33. A. Comella-Bolla, J. G. Orlandi, A. Miguez, M. Straccia, M. García-Bravo, G. Bombau, M. Galofré, P. Sanders, J. Carrere, J. C. Segovia, J. Blasi, N. D. Allen, J. Alberch, J. Soriano, J. M. Canals, Human Pluripotent Stem Cell-Derived Neurons Are Functionally Mature In Vitro and Integrate into the Mouse Striatum Following Transplantation. Mol. Neurobiol. 57, 2766–2798 (2020).

34. H. J. Kang, Y. I. Kawasawa, F. Cheng, Y. Zhu, X. Xu, M. Li, A. M. M. Sousa, M. Pletikos, K. A. Meyer, G. Sedmak, T. Guennel, Y. Shin, M. B. Johnson, Ž. Krsnik, S. Mayer, S. Fertuzinhos, S. Umlauf, S. N. Lisgo, A. Vortmeyer, D. R. Weinberger, S. Mane, T. M. Hyde, A. Huttner, M. Reimers, J. E. Kleinman, N. Šestan, Spatio-temporal transcriptome of the human brain. Nature. 478, 483–489 (2011).

35. K. R. Maynard, L. Collado-Torres, L. M. Weber, C. Uytingco, B. K. Barry, S. R. Williams, J. L. Catallini, M. N. Tran, Z. Besich, M. Tippani, J. Chew, Y. Yin, J. E. Kleinman, T. M. Hyde, N. Rao, S. C. Hicks, K. Martinowich, A. E. Jaffe, Transcriptome-scale spatial gene expression in the human dorsolateral prefrontal cortex. Nat. Neurosci. 24, 425–436 (2021).

36. R. R. Stickels, E. Murray, P. Kumar, J. Li, J. L. Marshall, D. J. Di Bella, P. Arlotta, E. Z. Macosko, F. Chen, Highly sensitive spatial transcriptomics at near-cellular resolution with Slide-seqV2. Nat. Biotechnol. 39, 313–319 (2021).

37. H. K. Finucane, Y. A. Reshef, V. Anttila, K. Slowikowski, A. Gusev, A. Byrnes, S. Gazal, P. R. Loh, C. Lareau, N. Shoresh, G. Genovese, A. Saunders, E. Macosko, S. Pollack, J. R. B. Perry, J. D. Buenrostro, B. E. Bernstein, S. Raychaudhuri, S. McCarroll, B. M. Neale, A. L. Price, Heritability enrichment of specifically expressed genes identifies disease-relevant tissues and cell types. Nat. Genet. 50, 621–629 (2018).

38. F. Koopmans, P. van Nierop, M. Andres-Alonso, A. Byrnes, T. Cijsouw, M. P. Coba, L. N. Cornelisse, R. J. Farrell, H. L. Goldschmidt, D. P. Howrigan, N. K. Hussain, C. Imig, A. P. H. de Jong, H. Jung, M. Kohansalnodehi, B. Kramarz, N. Lipstein, R. C. Lovering, H. MacGillavry, V. Mariano, H. Mi, M. Ninov, D. Osumi-Sutherland, R. Pielot, K.-H. Smalla, H. Tang, K. Tashman, R. F. G. Toonen, C. Verpelli, R. Reig-Viader, K. Watanabe, J. van Weering, T. Achsel, G. Ashrafi, N. Asi, T. C. Brown, P. De Camilli, M. Feuermann, R. E. Foulger, P. Gaudet, A. Joglekar, A. Kanellopoulos, R. Malenka, R. A. Nicoll, C. Pulido, J. de Juan-Sanz, M. Sheng, T. C. Südhof, H. U. Tilgner, C. Bagni, À. Bayés, T. Biederer, N. Brose, J. J. E. Chua, D. C. Dieterich, E. D. Gundelfinger, C. Hoogenraad, R. L. Huganir, R. Jahn, P. S. Kaeser, E. Kim, M. R. Kreutz, P. S. McPherson, B. M. Neale, V. O’Connor, D. Posthuma, T. A. Ryan, C. Sala, G. Feng, S. E. Hyman, P. D. Thomas, A. B. Smit, M. Verhage, SynGO: An Evidence-Based, Expert-Curated Knowledge Base for the Synapse. Neuron. 103, 217–234.e4 (2019).

39. R. Yang, K. K. Walder-Christensen, N. Kim, D. Wu, D. N. Lorenzo, A. Badea, Y.-H. Jiang, H. H. Yin, W. C. Wetsel, V. Bennett, ANK2 autism mutation targeting giant ankyrin-B promotes axon branching and ectopic connectivity. Proc. Natl. Acad. Sci. U. S. A. 116, 15262–15271 (2019).

40. M. J. Gandal, P. Zhang, E. Hadjimichael, R. L. Walker, C. Chen, S. Liu, H. Won, H. van Bakel, M. Varghese, Y. Wang, A. W. Shieh, J. Haney, S. Parhami, J. Belmont, M. Kim, P. Moran Losada, Z. Khan, J. Mleczko, Y. Xia, R. Dai, D. Wang, Y. T. Yang, M. Xu, K. Fish, P. R. Hof, J. Warrell, D. Fitzgerald, K. White, A. E. Jaffe, PsychENCODE Consortium, M. A. Peters, M. Gerstein, C. Liu, L. M. Iakoucheva, D. Pinto, D. H. Geschwind, Transcriptome-wide isoform-level dysregulation in ASD, schizophrenia, and bipolar disorder. Science. 362 (2018), doi:10.1126/science.aat8127.

41. M. Ashburner, C. A. Ball, J. A. Blake, D. Botstein, H. Butler, J. M. Cherry, A. P. Davis, K. Dolinski, S. S. Dwight, J. T. Eppig, M. A. Harris, D. P. Hill, L. Issel-Tarver, A. Kasarskis, S. Lewis, J. C. Matese, J. E. Richardson, M. Ringwald, G. M. Rubin, G. Sherlock, Gene ontology: tool for the unification of biology. The Gene Ontology Consortium. Nat Genet. 25, 25–29 (2000).

42. The Gene Ontology Consortium, The Gene Ontology Resource: 20 years and still GOing strong. Nucleic Acids Res. 47, D330–D338 (2019).

43. E. F. Spence, D. J. Kanak, B. R. Carlson, S. H. Soderling, The Arp2/3 Complex Is Essential for Distinct Stages of Spine Synapse Maturation, Including Synapse Unsilencing. J. Neurosci. 36, 9696–709 (2016).

44. A. Carracedo, P. P. Pandolfi, The PTEN-PI3K pathway: of feedbacks and cross-talks. Oncogene. 27, 5527–41 (2008).

45. F. Zahedi Abghari, Y. Moradi, M. Akouchekian, PTEN gene mutations in patients with macrocephaly and classic autism: A systematic review. Med. J. Islam. Repub. Iran. 33, 10 (2019).

46. A. K. Tilot, T. W. Frazier, C. Eng, Balancing Proliferation and Connectivity in PTEN-associated Autism Spectrum Disorder. Neurotherapeutics. 12, 609–19 (2015).

47. C.-J. Chen, M. Sgritta, J. Mays, H. Zhou, R. Lucero, J. Park, I.-C. Wang, J. H. Park, B. A. Kaipparettu, L. Stoica, P. Jafar-Nejad, F. Rigo, J. Chin, J. L. Noebels, M. Costa-Mattioli, Therapeutic inhibition of mTORC2 rescues the behavioral and neurophysiological abnormalities associated with Pten-deficiency. Nat. Med. 25, 1684–1690 (2019).

48. H. Huang, H. Weng, W. Sun, X. Qin, H. Shi, H. Wu, B. S. Zhao, A. Mesquita, C. Liu, C. L. Yuan, Y.-C. Hu, S. Hüttelmaier, J. R. Skibbe, R. Su, X. Deng, L. Dong, M. Sun, C. Li, S. Nachtergaele, Y. Wang, C. Hu, K. Ferchen, K. D. Greis, X. Jiang, M. Wei, L. Qu, J.-L. Guan, C. He, J. Yang, J. Chen, Recognition of RNA N6-methyladenosine by IGF2BP proteins enhances mRNA stability and translation. Nat. Cell Biol. 20, 285–295 (2018).

49. J. Grove, S. Ripke, T. D. Als, M. Mattheisen, R. K. Walters, H. Won, J. Pallesen, E. Agerbo, O. A. Andreassen, R. Anney, S. Awashti, R. Belliveau, F. Bettella, J. D. Buxbaum, J. Bybjerg-Grauholm, M. Bækvad-Hansen, F. Cerrato, K. Chambert, J. H. Christensen, C. Churchhouse, K. Dellenvall, D. Demontis, S. De Rubeis, B. Devlin, S. Djurovic, A. L. Dumont, J. I. Goldstein, C. S. Hansen, M. E. Hauberg, M. V. Hollegaard, S. Hope, D. P. Howrigan, H. Huang, C. M. Hultman, L. Klei, J. Maller, J. Martin, A. R. Martin, J. L. Moran, M. Nyegaard, T. Nærland, D. S. Palmer, A. Palotie, C. B. Pedersen, M. G. Pedersen, T. dPoterba, J. B. Poulsen, B. S. Pourcain, P. Qvist, K. Rehnström, A. Reichenberg, J. Reichert, E. B. Robinson, K. Roeder, P. Roussos, E. Saemundsen, S. Sandin, F. K. Satterstrom, G. Davey Smith, H. Stefansson, S. Steinberg, C. R. Stevens, P. F. Sullivan, P. Turley, G. B. Walters, X. Xu, N. R. Wray, M. Trzaskowski, E. M. Byrne, A. Abdellaoui, M. J. Adams, T. M. Air, T. F. M. Andlauer, S. A. Bacanu, A. T. F. Beekman, T. B. Bigdeli, E. B. Binder, D. H. R. Blackwood, J. Bryois, H. N. Buttenschøn, N. Cai, E. Castelao, T. K. Clarke, J. R. I. Coleman, L. Colodro-Conde, B. Couvy-Duchesne, N. Craddock, G. E. Crawford, G. Davies, I. J. Deary, F. Degenhardt, E. M. Derks, N. Direk, C. V. Dolan, E. C. Dunn, T. C. Eley, V. Escott-Price, F. F. H. Kiadeh, H. K. Finucane, A. J. Forstner, J. Frank, H. A. Gaspar, M. Gill, F. S. Goes, S. D. Gordon, L. S. Hall, T. F. Hansen, S. Herms, I. B. Hickie, P. Hoffmann, G. Homuth, C. Horn, J. J. Hottenga, M. Ising, R. Jansen, E. Jorgenson, J. A. Knowles, I. S. Kohane, J. Kraft, W. W. Kretzschmar, J. Krogh, Z. Kutalik, Y. Li, P. A. Lind, D. J. MacIntyre, D. F. MacKinnon, R. M. Maier, W. Maier, J. Marchini, H. Mbarek, P. McGrath, P. McGuffin, S. E. Medland, D. Mehta, C. M. Middeldorp, E. Mihailov, Y. Milaneschi, L. Milani, F. M. Mondimore, G. W. Montgomery, S. Mostafavi, N. Mullins, M. Nauck, B. Ng, M. G. Nivard, D. R. Nyholt, P. F. O’Reilly, H. Oskarsson, M. J. Owen, J. N. Painter, R. E. Peterson, E. Pettersson, W. J. Peyrot, G. Pistis, D. Posthuma, J. A. Quiroz, J. P. Rice, B. P. Riley, M. Rivera, S. S. Mirza, R. Schoevers, E. C. Schulte, L. Shen, J. Shi, S. I. Shyn, E. Sigurdsson, G. C. B. Sinnamon, J. H. Smit, D. J. Smith, F. Streit, J. Strohmaier, K. E. Tansey, H. Teismann, A. Teumer, W. Thompson, P. A. Thomson, T. E. Thorgeirsson, M. Traylor, J. Treutlein, V. Trubetskoy, A. G. Uitterlinden, D. Umbricht, S. Van der Auwera, A. M. van Hemert, A. Viktorin, P. M. Visscher, Y. Wang, B. T. Webb, S. M. Weinsheimer, J. Wellmann, G. Willemsen, S. H. Witt, Y. Wu, H. S. Xi, J. Yang, F. Zhang, V. Arolt, B. T. Baune, K. Berger, D. I. Boomsma, S. Cichon, U. Dannlowski, E. J. C. de Geus, J. R. DePaulo, E. Domenici, K. Domschke, T. Esko, H. J. Grabe, S. P. Hamilton, C. Hayward, A. C. Heath, K. S. Kendler, S. Kloiber, G. Lewis, Q. S. Li, S. Lucae, P. A. F. Madden, P. K. Magnusson, N. G. Martin, A. M. McIntosh, A. Metspalu, B. Müller-Myhsok, M. M. Nöthen, M. C. O’Donovan, S. A. Paciga, N. L. Pedersen, B. W. J. H. Penninx, R. H. Perlis, D. J. Porteous, J. B. Potash, M. Preisig, M. Rietschel, C. Schaefer, T. G. Schulze, J. W. Smoller, H. Tiemeier, R. Uher, H. Völzke, M. M. Weissman, C. M. Lewis, D. F. Levinson, G. Breen, M. Agee, B. Alipanahi, A. Auton, R. K. Bell, K. Bryc, S. L. Elson, P. Fontanillas, N. A. Furlotte, B. S. Hromatka, K. E. Huber, A. Kleinman, N. K. Litterman, M. H. McIntyre, J. L. Mountain, E. S. Noblin, C. A. M. Northover, S. J. Pitts, J. F. Sathirapongsasuti, O. V. Sazonova, J. F. Shelton, S. Shringarpure, J. Y. Tung, V. Vacic, C. H. Wilson, K. Stefansson, D. H. Geschwind, M. Nordentoft, D. M. Hougaard, T. Werge, O. Mors, P. B. Mortensen, B. M. Neale, M. J. Daly, A. D. Børglum, Identification of common genetic risk variants for autism spectrum disorder. Nat. Genet. 51, 431–444 (2019).

50. C. A. de Leeuw, J. M. Mooij, T. Heskes, D. Posthuma, MAGMA: generalized gene-set analysis of GWAS data. PLoS Comput. Biol. 11, e1004219 (2015).

51. A. Duchon, Y. Herault, DYRK1A, a Dosage-Sensitive Gene Involved in Neurodevelopmental Disorders, Is a Target for Drug Development in Down Syndrome. Front. Behav. Neurosci. 10, 104 (2016).

52. T. Dang, W. Y. Duan, B. Yu, D. L. Tong, C. Cheng, Y. F. Zhang, W. Wu, K. Ye, W. X. Zhang, M. Wu, B. B. Wu, Y. An, Z. L. Qiu, B. L. Wu, Autism-associated Dyrk1a truncation mutants impair neuronal dendritic and spine growth and interfere with postnatal cortical development. Mol. Psychiatry. 23, 747–758 (2018).

53. P. Fernández-Martínez, C. Zahonero, P. Sánchez-Gómez, DYRK1A: the double-edged kinase as a protagonist in cell growth and tumorigenesis. Mol. Cell. Oncol. 2, e970048.

54. J. Kaplanis, K. E. Samocha, L. Wiel, Z. Zhang, K. J. Arvai, R. Y. Eberhardt, G. Gallone, S. H. Lelieveld, H. C. Martin, J. F. McRae, P. J. Short, R. I. Torene, E. de Boer, P. Danecek, E. J. Gardner, N. Huang, J. Lord, I. Martincorena, R. Pfundt, M. R. F. Reijnders, A. Yeung, H. G. Yntema, Deciphering Developmental Disorders Study, L. E. L. M. Vissers, J. Juusola, C. F. Wright, H. G. Brunner, H. V Firth, D. R. FitzPatrick, J. C. Barrett, M. E. Hurles, C. Gilissen, K. Retterer, Evidence for 28 genetic disorders discovered by combining healthcare and research data. Nature. 586, 757–762 (2020).

55. T. Singh, B. M. Neale, M. J. Daly, Exome sequencing identifies rare coding variants in 10 genes which confer substantial risk for schizophrenia. *medRxiv* [preprint]. https://doi.org/10.1101/2020.09.18.20192815 (2020).

56. K. J. Karczewski, L. C. Francioli, G. Tiao, B. B. Cummings, J. Alföldi, Q. Wang, R. L. Collins, K. M. Laricchia, A. Ganna, D. P. Birnbaum, L. D. Gauthier, H. Brand, M. Solomonson, N. A. Watts, D. Rhodes, M. Singer-Berk, E. M. England, E. G. Seaby, J. A. Kosmicki, R. K. Walters, K. Tashman, Y. Farjoun, E. Banks, T. Poterba, A. Wang, C. Seed, N. Whiffin, J. X. Chong, K. E. Samocha, E. Pierce-Hoffman, Z. Zappala, A. H. O’Donnell-Luria, E. V. Minikel, B. Weisburd, M. Lek, J. S. Ware, C. Vittal, I. M. Armean, L. Bergelson, K. Cibulskis, K. M. Connolly, M. Covarrubias, S. Donnelly, S. Ferriera, S. Gabriel, J. Gentry, N. Gupta, T. Jeandet, D. Kaplan, C. Llanwarne, R. Munshi, S. Novod, N. Petrillo, D. Roazen, V. Ruano-Rubio, A. Saltzman, M. Schleicher, J. Soto, K. Tibbetts, C. Tolonen, G. Wade, M. E. Talkowski, Genome Aggregation Database Consortium, B. M. Neale, M. J. Daly, D. G. MacArthur, The mutational constraint spectrum quantified from variation in 141,456 humans. Nature. 581, 434–443 (2020).

57. D. Demontis, R. K. Walters, J. Martin, M. Mattheisen, T. D. Als, E. Agerbo, G. Baldursson, R. Belliveau, J. Bybjerg-Grauholm, M. Bækvad-Hansen, F. Cerrato, K. Chambert, C. Churchhouse, A. Dumont, N. Eriksson, M. Gandal, J. I. Goldstein, K. L. Grasby, J. Grove, O. O. Gudmundsson, C. S. Hansen, M. E. Hauberg, M. V Hollegaard, D. P. Howrigan, H. Huang, J. B. Maller, A. R. Martin, N. G. Martin, J. Moran, J. Pallesen, D. S. Palmer, C. B. Pedersen, M. G. Pedersen, T. Poterba, J. B. Poulsen, S. Ripke, E. B. Robinson, F. K. Satterstrom, H. Stefansson, C. Stevens, P. Turley, G. B. Walters, H. Won, M. J. Wright, ADHD Working Group of the Psychiatric Genomics Consortium (PGC), Early Lifecourse & Genetic Epidemiology (EAGLE) Consortium, 23andMe Research Team, O. A. Andreassen, P. Asherson, C. L. Burton, D. I. Boomsma, B. Cormand, S. Dalsgaard, B. Franke, J. Gelernter, D. Geschwind, H. Hakonarson, J. Haavik, H. R. Kranzler, J. Kuntsi, K. Langley, K.-P. Lesch, C. Middeldorp, A. Reif, L. A. Rohde, P. Roussos, R. Schachar, P. Sklar, E. J. S. Sonuga-Barke, P. F. Sullivan, A. Thapar, J. Y. Tung, I. D. Waldman, S. E. Medland, K. Stefansson, M. Nordentoft, D. M. Hougaard, T. Werge, O. Mors, P. B. Mortensen, M. J. Daly, S. V Faraone, A. D. Børglum, B. M. Neale, Discovery of the first genome-wide significant risk loci for attention deficit/hyperactivity disorder. Nat. Genet. 51, 63–75 (2019).

58. E. A. Stahl, G. Breen, A. J. Forstner, A. McQuillin, S. Ripke, V. Trubetskoy, M. Mattheisen, Y. Wang, J. R. I. Coleman, H. A. Gaspar, C. A. de Leeuw, S. Steinberg, J. M. W. Pavlides, M. Trzaskowski, E. M. Byrne, T. H. Pers, P. A. Holmans, A. L. Richards, L. Abbott, E. Agerbo, H. Akil, D. Albani, N. Alliey-Rodriguez, T. D. Als, A. Anjorin, V. Antilla, S. Awasthi, J. A. Badner, M. Bækvad-Hansen, J. D. Barchas, N. Bass, M. Bauer, R. Belliveau, S. E. Bergen, C. B. Pedersen, E. Bøen, M. P. Boks, J. Boocock, M. Budde, W. Bunney, M. Burmeister, J. Bybjerg-Grauholm, W. Byerley, M. Casas, F. Cerrato, P. Cervantes, K. Chambert, A. W. Charney, D. Chen, C. Churchhouse, T.-K. Clarke, W. Coryell, D. W. Craig, C. Cruceanu, D. Curtis, P. M. Czerski, A. M. Dale, S. de Jong, F. Degenhardt, J. Del-Favero, J. R. DePaulo, S. Djurovic, A. L. Dobbyn, A. Dumont, T. Elvsåshagen, V. Escott-Price, C. C. Fan, S. B. Fischer, M. Flickinger, T. M. Foroud, L. Forty, J. Frank, C. Fraser, N. B. Freimer, L. Frisén, K. Gade, D. Gage, J. Garnham, C. Giambartolomei, M. G. Pedersen, J. Goldstein, S. D. Gordon, K. Gordon-Smith, E. K. Green, M. J. Green, T. A. Greenwood, J. Grove, W. Guan, J. Guzman-Parra, M. L. Hamshere, M. Hautzinger, U. Heilbronner, S. Herms, M. Hipolito, P. Hoffmann, D. Holland, L. Huckins, S. Jamain, J. S. Johnson, A. Juréus, R. Kandaswamy, R. Karlsson, J. L. Kennedy, S. Kittel-Schneider, J. A. Knowles, M. Kogevinas, A. C. Koller, R. Kupka, C. Lavebratt, J. Lawrence, W. B. Lawson, M. Leber, P. H. Lee, S. E. Levy, J. Z. Li, C. Liu, S. Lucae, A. Maaser, D. J. MacIntyre, P. B. Mahon, W. Maier, L. Martinsson, S. McCarroll, P. McGuffin, M. G. McInnis, J. D. McKay, H. Medeiros, S. E. Medland, F. Meng, L. Milani, G. W. Montgomery, D. W. Morris, T. W. Mühleisen, N. Mullins, H. Nguyen, C. M. Nievergelt, A. N. Adolfsson, E. A. Nwulia, C. O’Donovan, L. M. O. Loohuis, A. P. S. Ori, L. Oruc, U. Ösby, R. H. Perlis, A. Perry, A. Pfennig, J. B. Potash, S. M. Purcell, E. J. Regeer, A. Reif, C. S. Reinbold, J. P. Rice, F. Rivas, M. Rivera, P. Roussos, D. M. Ruderfer, E. Ryu, C. Sánchez-Mora, A. F. Schatzberg, W. A. Scheftner, N. J. Schork, C. Shannon Weickert, T. Shehktman, P. D. Shilling, E. Sigurdsson, C. Slaney, O. B. Smeland, J. L. Sobell, C. Søholm Hansen, A. T. Spijker, D. St Clair, M. Steffens, J. S. Strauss, F. Streit, J. Strohmaier, S. Szelinger, R. C. Thompson, T. E. Thorgeirsson, J. Treutlein, H. Vedder, W. Wang, S. J. Watson, T. W. Weickert, S. H. Witt, S. Xi, W. Xu, A. H. Young, P. Zandi, P. Zhang, S. Zöllner, eQTLGen Consortium, BIOS Consortium, R. Adolfsson, I. Agartz, M. Alda, L. Backlund, B. T. Baune, F. Bellivier, W. H. Berrettini, J. M. Biernacka, D. H. R. Blackwood, M. Boehnke, A. D. Børglum, A. Corvin, N. Craddock, M. J. Daly, U. Dannlowski, T. Esko, B. Etain, M. Frye, J. M. Fullerton, E. S. Gershon, M. Gill, F. Goes, M. Grigoroiu-Serbanescu, J. Hauser, D. M. Hougaard, C. M. Hultman, I. Jones, L. A. Jones, R. S. Kahn, G. Kirov, M. Landén, M. Leboyer, C. M. Lewis, Q. S. Li, J. Lissowska, N. G. Martin, F. Mayoral, S. L. McElroy, A. M. McIntosh, F. J. McMahon, I. Melle, A. Metspalu, P. B. Mitchell, G. Morken, O. Mors, P. B. Mortensen, B. Müller-Myhsok, R. M. Myers, B. M. Neale, V. Nimgaonkar, M. Nordentoft, M. M. Nöthen, M. C. O’Donovan, K. J. Oedegaard, M. J. Owen, S. A. Paciga, C. Pato, M. T. Pato, D. Posthuma, J. A. Ramos-Quiroga, M. Ribasés, M. Rietschel, G. A. Rouleau, M. Schalling, P. R. Schofield, T. G. Schulze, A. Serretti, J. W. Smoller, H. Stefansson, K. Stefansson, E. Stordal, P. F. Sullivan, G. Turecki, A. E. Vaaler, E. Vieta, J. B. Vincent, T. Werge, J. I. Nurnberger, N. R. Wray, A. Di Florio, H. J. Edenberg, S. Cichon, R. A. Ophoff, L. J. Scott, O. A. Andreassen, J. Kelsoe, P. Sklar, Bipolar Disorder Working Group of the Psychiatric Genomics Consortium, Genome-wide association study identifies 30 loci associated with bipolar disorder. Nat. Genet. 51, 793– 803 (2019).

59. D. M. Howard, M. J. Adams, T.-K. Clarke, J. D. Hafferty, J. Gibson, M. Shirali, J. R. I. Coleman, S. P. Hagenaars, J. Ward, E. M. Wigmore, C. Alloza, X. Shen, M. C. Barbu, E. Y. Xu, H. C. Whalley, R. E. Marioni, D. J. Porteous, G. Davies, I. J. Deary, G. Hemani, K. Berger, H. Teismann, R. Rawal, V. Arolt, B. T. Baune, U. Dannlowski, K. Domschke, C. Tian, D. A. Hinds, 23andMe Research Team, Major Depressive Disorder Working Group of the Psychiatric Genomics Consortium, M. Trzaskowski, E. M. Byrne, S. Ripke, D. J. Smith, P. F. Sullivan, N. R. Wray, G. Breen, C. M. Lewis, A. M. McIntosh, Genome-wide meta-analysis of depression identifies 102 independent variants and highlights the importance of the prefrontal brain regions. Nat. Neurosci. 22, 343–352 (2019).

60. Schizophrenia Working Group of the Psychiatric Genomics Consortium, Biological insights from 108 schizophrenia-associated genetic loci. Nature. 511, 421–7 (2014).

61. L. Yengo, J. Sidorenko, K. E. Kemper, Z. Zheng, A. R. Wood, M. N. Weedon, T. M. Frayling, J. Hirschhorn, J. Yang, P. M. Visscher, Meta-analysis of genome-wide association studies for height and body mass index in ∼700 000 individuals of European ancestry. Hum. Mol. Genet. 27, 3641–3649 (2018).

62. M. Losh, D. Childress, K. Lam, J. Piven, Defining key features of the broad autism phenotype: a comparison across parents of multiple- and single-incidence autism families. Am. J. Med. Genet. B. Neuropsychiatr. Genet. **147B**, 424–33 (2008).

63. R. Keller, R. Basta, L. Salerno, M. Elia, Autism, epilepsy, and synaptopathies: a not rare association. Neurol. Sci. 38, 1353–1361 (2017).

64. R. M. Busch, S. Srivastava, O. Hogue, T. W. Frazier, P. Klaas, A. Hardan, J. A. Martinez-Agosto, M. Sahin, C. Eng, Developmental Synaptopathies Consortium, Neurobehavioral phenotype of autism spectrum disorder associated with germline heterozygous mutations in PTEN. Transl. Psychiatry. 9, 253 (2019).

65. D. V Hansen, J. L. R. Rubenstein, A. R. Kriegstein, Deriving excitatory neurons of the neocortex from pluripotent stem cells. Neuron. 70, 645–60 (2011).

66. V. Busskamp, N. E. Lewis, P. Guye, A. H. M. Ng, S. L. Shipman, S. M. Byrne, N. E. Sanjana, J. Murn, Y. Li, S. Li, M. Stadler, R. Weiss, G. M. Church, Rapid neurogenesis through transcriptional activation in human stem cells. Mol. Syst. Biol. 10, 760 (2014).

67. F. B. Russo, A. Brito, A. M. de Freitas, A. Castanha, B. C. de Freitas, P. C. B. Beltrão-Braga, The use of iPSC technology for modeling Autism Spectrum Disorders. Neurobiol. Dis. 130, 104483 (2019).

68. V. Bennett, K. Walder, Evolution in action: giant ankyrins awake. Dev. Cell. 33, 1–2 (2015).

69. T. C. Roberts, R. Langer, M. J. A. Wood, Advances in oligonucleotide drug delivery. Nat. Rev. Drug Discov. 19, 673–694 (2020).

70. C. H. Melick, D. Meng, J. L. Jewell, A-kinase anchoring protein 8L interacts with mTORC1 and promotes cell growth. J. Biol. Chem. 295, 8096–8105 (2020).

71. R. A. Nebel, J. Kirschen, J. Cai, Y. J. Woo, K. Cherian, B. S. Abrahams, Reciprocal Relationship between Head Size, an Autism Endophenotype, and Gene Dosage at 19p13.12 Points to AKAP8 and AKAP8L. PLoS One. 10, e0129270 (2015).

72. V. Janjić, N. Pržulj, Biological function through network topology: a survey of the human diseasome. Brief. Funct. Genomics. 11, 522–32 (2012).

73. M. J. Gandal, J. R. Haney, N. N. Parikshak, V. Leppa, G. Ramaswami, C. Hartl, A. J. Schork, V. Appadurai, A. Buil, T. M. Werge, C. Liu, K. P. White, CommonMind Consortium, PsychENCODE Consortium, iPSYCH-BROAD Working Group, S. Horvath, D. H. Geschwind, Shared molecular neuropathology across major psychiatric disorders parallels polygenic overlap. Science. 359, 693–697 (2018).

74. Y. Joo, D. R. Benavides, Local Protein Translation and RNA Processing of Synaptic Proteins in Autism Spectrum Disorder. Int. J. Mol. Sci. 22 (2021), doi:10.3390/ijms22062811.

75. P. J. Batista, B. Molinie, J. Wang, K. Qu, J. Zhang, L. Li, D. M. Bouley, E. Lujan, B. Haddad, K. Daneshvar, A. C. Carter, R. A. Flynn, C. Zhou, K.-S. Lim, P. Dedon, M. Wernig, A. C. Mullen, Y. Xing, C. C. Giallourakis, H. Y. Chang, m(6)A RNA modification controls cell fate transition in mammalian embryonic stem cells. Cell Stem Cell. 15, 707–19 (2014).

76. S. Geula, S. Moshitch-Moshkovitz, D. Dominissini, A. A. Mansour, N. Kol, M. Salmon-Divon, V. Hershkovitz, E. Peer, N. Mor, Y. S. Manor, M. S. Ben-Haim, E. Eyal, S. Yunger, Y. Pinto, D. A. Jaitin, S. Viukov, Y. Rais, V. Krupalnik, E. Chomsky, M. Zerbib, I. Maza, Y. Rechavi, R. Massarwa, S. Hanna, I. Amit, E. Y. Levanon, N. Amariglio, N. Stern-Ginossar, N. Novershtern, G. Rechavi, J. H. Hanna, Stem cells. m6A mRNA methylation facilitates resolution of naïve pluripotency toward differentiation. Science. 347, 1002–6 (2015).

77. C.-X. Wang, G.-S. Cui, X. Liu, K. Xu, M. Wang, X.-X. Zhang, L.-Y. Jiang, A. Li, Y. Yang, W.-Y. Lai, B.-F. Sun, G.-B. Jiang, H.-L. Wang, W.-M. Tong, W. Li, X.-J. Wang, Y.-G. Yang, Q. Zhou, METTL3-mediated m6A modification is required for cerebellar development. PLoS Biol. 16, e2004880 (2018).

78. K.-J. Yoon, F. R. Ringeling, C. Vissers, F. Jacob, M. Pokrass, D. Jimenez-Cyrus, Y. Su, N.-S. Kim, Y. Zhu, L. Zheng, S. Kim, X. Wang, L. C. Doré, P. Jin, S. Regot, X. Zhuang, S. Canzar, C. He, G.-L. Ming, H. Song, Temporal Control of Mammalian Cortical Neurogenesis by m6A Methylation. Cell. 171, 877–889.e17 (2017).

79. B. M. Edens, C. Vissers, J. Su, S. Arumugam, Z. Xu, H. Shi, N. Miller, F. Rojas Ringeling, G.-L. Ming, C. He, H. Song, Y. C. Ma, FMRP Modulates Neural Differentiation through m6A-Dependent mRNA Nuclear Export. Cell Rep. 28, 845–854.e5 (2019).

80. T. J. Samuels, A. I. Järvelin, D. Ish-Horowicz, I. Davis, Imp/IGF2BP levels modulate individual neural stem cell growth and division through myc mRNA stability. Elife. 9 (2020), doi:10.7554/eLife.51529.

81. Y. Ogawa, Y. Nonaka, T. Goto, E. Ohnishi, T. Hiramatsu, I. Kii, M. Yoshida, T. Ikura, H. Onogi, H. Shibuya, T. Hosoya, N. Ito, M. Hagiwara, Development of a novel selective inhibitor of the Down syndrome-related kinase Dyrk1A. Nat. Commun. 1, 86 (2010).

82. F. Krueger, Trim Galore!: a wrapper tool around Cutadapt and FastQC to consistently apply quality and adapter trimming to FastQ files. https://github.com/FelixKrueger/TrimGalore.

83. D. Kim, J. M. Paggi, C. Park, C. Bennett, S. L. Salzberg, Graph-based genome alignment and genotyping with HISAT2 and HISAT-genotype. Nat. Biotechnol. 37, 907–915 (2019).

84. S. Anders, P. T. Pyl, W. Huber, HTSeq--a Python framework to work with high-throughput sequencing data. Bioinformatics. 31, 166–9 (2015).

85. A. D. Yates, P. Achuthan, W. Akanni, J. Allen, J. Allen, J. Alvarez-Jarreta, M. R. Amode, I. M. Armean, A. G. Azov, R. Bennett, J. Bhai, K. Billis, S. Boddu, J. C. Marugán, C. Cummins, C. Davidson, K. Dodiya, R. Fatima, A. Gall, C. G. Giron, L. Gil, T. Grego, L. Haggerty, E. Haskell, T. Hourlier, O. G. Izuogu, S. H. Janacek, T. Juettemann, M. Kay, I. Lavidas, T. Le, D. Lemos, J. G. Martinez, T. Maurel, M. McDowall, A. McMahon, S. Mohanan, B. Moore, M. Nuhn, D. N. Oheh, A. Parker, A. Parton, M. Patricio, M. P. Sakthivel, A. I. Abdul Salam, B. M. Schmitt, H. Schuilenburg, D. Sheppard, M. Sycheva, M. Szuba, K. Taylor, A. Thormann, G. Threadgold, A. Vullo, B. Walts, A. Winterbottom, A. Zadissa, M. Chakiachvili, B. Flint, A. Frankish, S. E. Hunt, G. IIsley, M. Kostadima, N. Langridge, J. E. Loveland, F. J. Martin, J. Morales, J. M. Mudge, M. Muffato, E. Perry, M. Ruffier, S. J. Trevanion, F. Cunningham, K. L. Howe, D. R. Zerbino, P. Flicek, Ensembl 2020. Nucleic Acids Res. 48, D682–D688 (2020).

86. M. D. Robinson, D. J. McCarthy, G. K. Smyth, edgeR: a Bioconductor package for differential expression analysis of digital gene expression data. Bioinformatics. 26, 139– 40 (2010).

87. T. Stuart, A. Butler, P. Hoffman, C. Hafemeister, E. Papalexi, W. M. Mauck, Y. Hao, M. Stoeckius, P. Smibert, R. Satija, Comprehensive Integration of Single-Cell Data. Cell. 177, 1888–1902.e21 (2019).

88. S. Tyanova, T. Temu, P. Sinitcyn, A. Carlson, M. Y. Hein, T. Geiger, M. Mann, J. Cox, The Perseus computational platform for comprehensive analysis of (prote)omics data. Nat. Methods. 13, 731–740 (2016).

89. M. E. Ritchie, B. Phipson, D. Wu, Y. Hu, C. W. Law, W. Shi, G. K. Smyth, limma powers differential expression analyses for RNA-sequencing and microarray studies. Nucleic Acids Res. 43, e47 (2015).

90. T. P. Quinn, M. F. Richardson, D. Lovell, T. M. Crowley, propr: An R-package for Identifying Proportionally Abundant Features Using Compositional Data Analysis. Sci. Rep. 7, 16252 (2017).

91. M. A. Skinnider, J. W. Squair, L. J. Foster, Evaluating measures of association for single-cell transcriptomics. Nat. Methods. 16, 381–386 (2019).

92. 1000 Genomes Project Consortium, A. Auton, L. D. Brooks, R. M. Durbin, E. P. Garrison, H. M. Kang, J. O. Korbel, J. L. Marchini, S. McCarthy, G. A. McVean, G. R. Abecasis, A global reference for human genetic variation. Nature. 526, 68–74 (2015).

93. G. Teo, G. Liu, J. Zhang, A. I. Nesvizhskii, A. C. Gingras, H. Choi, SAINTexpress: Improvements and additional features in Significance Analysis of INTeractome software. J. Proteomics. 100, 37–43 (2014).

94. P. Feist, A. B. Hummon, Proteomic challenges: sample preparation techniques for microgram-quantity protein analysis from biological samples. Int. J. Mol. Sci. 16, 3537– 63 (2015).

95. E. L. Huttlin, R. J. Bruckner, J. Navarrete-Perea, J. R. Cannon, K. Baltier, F. Gebreab, M. P. Gygi, A. Thornock, G. Zarraga, S. Tam, J. Szpyt, B. M. Gassaway, A. Panov, H. Parzen, S. Fu, A. Golbazi, E. Maenpaa, K. Stricker, S. Guha Thakurta, T. Zhang, R. Rad, J. Pan, D. P. Nusinow, J. A. Paulo, D. K. Schweppe, L. P. Vaites, J. W. Harper, S. P. Gygi, Dual proteome-scale networks reveal cell-specific remodeling of the human interactome. Cell. 184, 3022–3040.e28 (2021).

96. H. Moriya, Quantitative nature of overexpression experiments. Mol. Biol. Cell. 26, 3932–9 (2015).

97. G. Gou, A. Roca-Fernandez, M. Kilinc, E. Serrano, R. Reig-Viader, Y. Araki, R. L. Huganir, C. de Quintana-Schmidt, G. Rumbaugh, À. Bayés, SynGAP splice variants display heterogeneous spatio-temporal expression and subcellular distribution in the developing mammalian brain. J. Neurochem. 154, 618–634 (2020).

98. K. Kammers, R. N. Cole, C. Tiengwe, I. Ruczinski, Detecting Significant Changes in Protein Abundance. EuPA open proteomics. 7, 11–19 (2015).

99. E. L. Huttlin, L. Ting, R. J. Bruckner, F. Gebreab, M. P. Gygi, J. Szpyt, S. Tam, G. Zarraga, G. Colby, K. Baltier, R. Dong, V. Guarani, L. P. Vaites, A. Ordureau, R. Rad, B. K. Erickson, M. Wühr, J. Chick, B. Zhai, D. Kolippakkam, J. Mintseris, R. A. Obar, T. Harris, S. Artavanis-Tsakonas, M. E. Sowa, P. De Camilli, J. A. Paulo, J. W. Harper, S. P. Gygi, The BioPlex Network: A Systematic Exploration of the Human Interactome. Cell. 162, 425–440 (2015).

