## Supplementary Materials for "Interaction studies of risk proteins in human induced neurons reveal convergent biology and novel mechanisms underlying autism spectrum disorders"

**This PDF file includes:**

Materials & Methods

Supplementary Text 1 to 3

Supplementary Figures 1 to 10

Captions for Supplementary Tables 1 to 12

**Other Supplementary Materials for this manuscript include the following:**

Supplementary Tables 1 to 12 (Excel format)

#### Materials & Methods

##### Experimental Model Details

##### Human brain samples

Post-mortem brain tissue was obtained from the Netherlands Brain Bank (NBB) at the Netherlands Institute for Neuroscience (NIN), Amsterdam. Brain tissue was collected from two adult donors with written informed consent for brain autopsy and the use of the material and clinical information for research purposes has been obtained by the NBB. PMD was less than 12 hours.

##### Cell lines

Glutamatergic patterned induced neurons (iNs) were differentiated from a male induced pluripotent stem cell (iPSC) line (iPS hDFn 83/22 iNgn2#9 [iPS3] from (*31*)) by conditional expression of the neuralizing transcription factor NGN2, as described previously (*24*, *31*). A plate was coated with GelTrex (LifeTechnologies, A1413301) adhesion matrix (1:100 in DMEM/F:12, Life Technologies, Inc., 11320033) and cells were seeded at a density of 40,000 cells $cm^{-2}$ in Stemflex media (Life Technologies, Inc., A3349401) containing Geneticin as selective antibiotic (Life Technologies, Inc., 10131027) (1:400). Once the cells achieved 60% confluency, the monolayer was detached using Accutase (Life Technologies, Inc., A11105) and transferred onto new plates for cell expansion. Human embryonic kidney 293 (HEK293) cells were grown as monolayers in DMEM media (Life Technologies, 11320033) supplemented with 10% FBS (R&D Systems, S11195). Once the cells achieved 60% confluency, the monolayer was detached using Trypsin (Promega, V5111) and transferred onto new plates for cell expansion. Both iNs and HEK293 cells were incubated at 37°C and 5% $CO_2$.

#### Methods Details

##### Differentiation of iNs

hDFn cells, stably expressing TetO-Ngn2-Neo and reverse tetracycline-controlled transactivator (rtTA), were plated at a density of 40,000 cells $cm^{-2}$ with rock inhibitor Y27632 (Stemgent, 04-0012). Day 1 cells were differentiated in N2 media (Life Technologies, Inc.) supplemented with 10 μM SB431542 (Tocris, 1614), 2 μM XAV939 (Stemgent, 04-00046) and 100 nM LDN-193189 (Stemgent, 04-0074) along with doxycycline hyclate (2 μg $mL^{-1}$). Day 2 media was N2+SB/XAV/LDN/doxycycline hyclate and differentiation media were as previously described. On day 3, cell differentiation was continued in neurobasal media (Life Technologies, Inc.) supplemented with B27 (50X, Thermo Scientific), brain-derived neurotrophic factor (BDNF), ciliary neurotrophic factor (CTNF), glial cell-derived neurotrophic factor (GDNF) (R&D Systems 248-BD/CF, 257-NT/CF, and 212-GD/CF at 10 ng $mL^{-1}$) and doxycycline hyclate (2 μg $mL^{-1}$).

##### Protein extraction and immunoblotting

Total protein extract was obtained by harvesting cells and either processing them immediately or snap-freezing them on dry ice for storage at -80°C. In both cases, cell pellets were washed with PBS and resuspended in 10x packed cell volume (PCV) IP lysis buffer (Thermo Scientific), with freshly added Halt protease and phosphatase inhibitors (Thermo Scientific). After a 20 min incubation time at 4°C, cells were collected by centrifugation (16,200 g, 20 min, 4°C) and resuspended in 3x PCV lysis buffer. The concentration of the samples was quantified using the Thermo BCA protein assay and when not used immediately, samples were stored at -80°C. Samples for immunoblotting were diluted in 6x SMASH buffer (50 mM Tris HCl pH 6.8, 10%

glycerol, 2% SDS, 0.02% bromophenol blue, 1% b-mercaptoethanol), boiled for 10 min at 95°C, separated on a NuPAGE 4-12% Bis-Tris Protein Gel (Invitrogen), and transferred onto a PVDF membrane (Life Technologies) by wet transfer (100 V for 2 hours). Membranes were blocked by incubation for one hour at room temperature in 10 mL TBS and 0.1% Tween (TBST) with 5% w/v BioRad Blotting-grade Blocker. Blots were incubated overnight at 4°C with the primary antibody, washed 3 times for 10 min with TBST and incubated for 45 min with secondary antibody conjugated to horseradish peroxidase. After washing 3 times for 10 min with TBST, bands were visualized using SuperSignal™ West Femto Maximum Sensitivity Substrate (Thermo Scientific). "Forward" IP western validations included membranes loaded with immunoprecipitate of the index protein which was incubated overnight at 4°C with the primary antibody of the interactor protein, and "reverse" IP western validations included membranes loaded with immunoprecipitate of the interactor protein which was incubated overnight at 4°C with the primary antibody of the index protein. All antibodies used in this study (including those that were not IP-competent) are listed in **Supplementary Tables 1**, **2**, and **5**.

**Mouse cortex preparation**

Mouse cortices were isolated from p0 pups of C57BL/6 background, cut into small pieces, and flash frozen. Lysis was performed by adding Pierce IP lysis buffer (Pierce) to frozen pieces and mechanically dissociating them by vigorous pipetting. All subsequent steps were the same as those for iNs.

**Human brain preparation**

For human post-mortem brain samples, 0.6 g of tissue was homogenized in 34 mL extraction buffer (0.5% n-Dodecyl β-D-maltoside, 150 mM NaCl, 50 mM Hepes, pH 7.4) with a glass homogenizer (PotterS from B. Braun) set at 900 rpm for 12 strokes. The homogenate was centrifuged twice at 16,000g for 10 min. All subsequent steps were the same as those for iNs.

**Immunoprecipitations**

For each individual experiment, 1-2 mg of protein extract from the same cell differentiation batch was incubated at 4°C overnight in the presence of 1-2 µg of the relevant antibody. On the next day, 50 µL of Protein G beads (Pierce) were added to each sample and incubated at 4°C for 4 hours. Flow-through was collected and beads were washed once with 1 mL lysis buffer (Pierce) supplemented with Halt protease and phosphatase inhibitors (Thermo Scientific), and twice with PBS. Beads were resuspended in 60 µL of PBS and 10% of the volume was employed for immunoblotting, after being boiled in 6xSMASH buffer (50 mM Tris HCl pH 6.8, 10% Glycerol, 2% SDS, 0.02% bromophenol blue, 1% b-mercaptoethanol) for 10 min at 95ºC. The remaining volume was stored at -80°C and subsequently used for mass spectrometry analysis.

**Mass spectrometry**

**Supplementary Table 2** provides an overview of where liquid chromatography-tandem mass spectrometry (LC-MS/MS) for each IP-MS dataset was performed; MS protocols for each facility/site are described in more detail below.

*Broad Proteomics Platform (Broad):* Proteins in IP samples were digested on beads using 90 µL of digestion buffer (2 M urea / 50 mM Tris buffer with 1 mM DTT and 5 µg/mL Trypsin) for 1 hr, shaking at 1000 rpm. The suspension was then transferred to a new tube, and the beads were washed twice with 60 µL of wash buffer (2 M urea / 50 mM Tris buffer). The wash buffer was added to the suspension with digestion. The digestion and wash process was repeated a second time pooling the suspensions with the suspensions from the first round. The pooled solution was reduced using 4 mM DTT for 30 min at 25°C shaking at 1000 rpm. The proteins were then alkylated using 10 mM iodoacetamide and incubating for 45 min at 25°C shaking at 1000 rpm and protected from light. Proteins were then digested with 0.5 µg of trypsin overnight at 25°C shaking at 700 rpm. The next day proteins were quenched using 40 µL of 10% formic acid and desalted using an Oasis Cartridge. Samples were vacuum dried and labeled with iTRAQ4 kits (Sciex). Each iTRAQ 4-plex consisted of 2 replicate experimental IPs using an antibody against an index protein (labeled with iTRAQ reagent labels 114 and 115) and 2 replicate control IPs using a control IgG antibody (labeled with 116 and 117). Reconstituted peptides were separated on an online nanoflow EASY-nLC 1000 UHPLC system (Thermo Scientific) and analyzed on a benchtop Orbitrap Q Exactive Plus (DYRK1A, PTEN) or Q Exactive HF (SCN2A, SHANK3) mass spectrometer (Thermo Scientific). The peptide samples were injected onto a capillary column (Picofrit with 10 µm tip opening / 75 µm diameter, New Objective, PF360-75-10-N-5) packed in-house with 20 cm C18 silica material (1.9 µm ReproSil-Pur C18-AQ medium, Dr. Maisch GmbH, r119.aq). The UHPLC setup was connected with a custom-fit micro-adapting tee (360 µm, IDEX Health & Science, UH-753), and capillary columns were heated to 50 °C in column heater sleeves (Phoenix-ST) to reduce back-pressure during UHPLC separation. Injected peptides were separated at a flow rate of 200 nL/min with a linear 196 min (SCN2A, SHANK3, PTEN) or 234 min (DYRK1A) gradient from 94% solvent A (3% acetonitrile, 0.1% formic acid) to 30% (DYRK1A) or 40% (SCN2A, SHANK3, PTEN) solvent B (90% acetonitrile, 0.1%

formic acid), followed by a linear 8 or 9 min gradient to 60% (DYRK1A) or 65% (SCN2A, SHANK3, PTEN) solvent B and a 1 or 3 min ramp to 90% B. The Q Exactive instrument was operated in the data-dependent mode acquiring HCD MS/MS scans (R=17.5K –QE+ or R=45K-QE-HF) after each MS1 scan (R=70K-QE+ or R=60K-QE-HF) on the 12(QE+) or 16(QE-HF) most abundant ions using an MS1 ion target of 3x106 ions and an MS2 target of 5x104 ions. The maximum ion time utilized for the MS/MS scans was 120 ms; the HCD-normalized collision energy was set to 27; the dynamic exclusion time was set to 20s, and the peptide match preferred and isotope exclusion functions were enabled.

*Whitehead Proteomics Core Facility (Whitehead):* IP samples on beads were resuspended in 100 µL of 100 mM triethylammonium bicarbonate (TEAB), reduced using 2 µL of 50 mM tris(2-carboxyethyl)phosphine for 60 min at 60°C, and alkylated using 1 µL of 2% S-methyl methanethiosulfonate in isopropanol for 10 min at room temperature. Proteins were then digested with 250 ng of trypsin overnight at 37°C with gentle shaking. iTRAQ4 reagents (Sciex) were resuspended in 50 µL isopropanol and added to each sample followed by vortex and spin; the samples in each 4-plex consisted of 2 replicate experimental IPs using an antibody against an index protein (labeled with iTRAQ reagent labels 114 and 115) and 2 replicate control IPs using a control IgG antibody (labeled with 116 and 117). The samples were combined and incubated at room temperature for 2 hr, and then washed, extracted, and concentrated by solid phase extraction using Waters Sep-Pak Plus C18 cartridges. Organic solvent was removed and the sample volumes were reduced to 80 µL via speed vacuum. The labeled peptides were subjected to basic (high pH) reversed phase high performance liquid chromatography (HPLC) with fraction collection using Shimadzu LC-20AD pumps and a FRC-10A fraction collector. Samples were loaded on a 10 cm x 2.1 mm column packed with 2.6 micron Aeris PEPTIDE XB-C18 media (Phenomenex). The initial gradient condition was isocratic 1% buffer A (20 mM ammonium formate in water, pH = 10) at 150 µL $min^{-1}$, with increasing buffer B (acetonitrile) concentrations to 16.7% B at 20.5 min, 30% B at 31 min, and 45% B at 36 min. The column was washed with high percent B and re-equilibrated between analytical runs for a total cycle time of ~55 min. Sixteen 450 µL fractions (fx) were collected, combined into eight samples (fx1+2, fx3+9, fx4+10, fx5+11, fx6+12, fx7+13, fx8+14, fx15+16), then reduced to 20 µL via speed vacuum. The combined samples were subjected to reversed phase HPLC using Thermo EASY-nLC 1200 pumps and autosampler, followed by mass spectrometry using a Thermo Q Exactive HF-X Hybrid Quadrupole-Orbitrap mass spectrometer and a nanoflow configuration. Samples were loaded on a 6 cm x 100 micron column packed with 10 micron ODS-A C18 material (YMC), washed with 4 µL total volume to trap and wash peptides, then eluted onto the analytical column packed with 1.7 micron Aeris C18 material (Phenomenex) in a fritted 14 cm x 75 micron fused silica tubing pulled to a 5 micron tip. The initial gradient condition was 1% buffer A (1% formic acid in water) at 300 nL $min^{-1}$, with increasing buffer B (1% formic acid in acetonitrile) concentrations to 6% B at 1 min, 21% B at 42.5 min, 36% B at 63.15 min, and 50% B at 73 min. The column was washed with high percent B and re-equilibrated between analytical runs for a total cycle time of ~97 min. The mass spectrometer was operated in a data-dependent acquisition mode where the 20 most abundant peptides detected in the Orbitrap using full scan mode with a resolution of 60,000 were subjected to daughter ion fragmentation using a resolution of 15,000. A running list of parent ions was tabulated to an exclusion list to increase the number of peptides analyzed throughout the chromatographic run.

*Center for Neurogenomics and Cognitive Research (CNCR):* IP samples were treated and digested by two protocols. For in-gel digestion, proteins were briefly separated by SDS-PAGE electrophoresis. Each sample was split in three fractions covering protein masses of >70kDa, 70-40 kDa and <40 kDa, and proteins were digested by trypsin overnight at 37ºC. Filter-aided sample preparation (FASP) protocol was used for human brain post-mortem samples. Sample was solubilized in 2% SDS, 100 mM Tris (pH = 8.8) and transferred to the Microcon-30 filter tube (Millipore). After serial washes with 8M urea and 50 mM $NH_4HCO_3$, proteins were digested by trypsin overnight at 37ºC and collected as a single fraction. The resulting peptides were analyzed by the TripleTOF 5600+ mass spectrometer coupled to an Ultimate 3000 LC system (Dionex, Thermo Scientific). The mass spectrometer was run in micro-mode. Peptides were fractionated on a 200 mm Alltima C18 column (300 µm i.d., 3 µm particle size) at a flow rate of 5 µL $min^{-1}$. For in-gel digestion protocol, the tryptic peptides from each gel slice were analyzed with a 1 hr LC gradient. For FASP protocol, a 2 hr LC gradient was used. The eluted peptides were electro-sprayed with a micro-spray needle voltage of 5500 V. The MS survey scan range was 350–1250 m/z acquired for 250 ms. The top 20 precursor ions were selected for 90 ms per MS/MS acquisition, with a threshold of 90 counts. Dynamic exclusion was 10 s. Rolling CID function was activated, with an energy spread of 5 eV.

*Harvard Center for Mass Spectrometry (Harvard):* IP samples were stored in PBS buffer on beads. PBS was removed and samples were dissolved in 50 µL of 50 mM TEAB, followed by trypsin (Promega) digestion for 3 hr at 38 °C. Digested samples were dried to 20 µL and 10 µL of each sample was injected in the mass spectrometer. LC-MS/MS was performed on a Lumos Tribrid Orbitrap Mass Spectrometer (Thermo Scientific) equipped with Ultimate 3000 (Thermo Scientific) nano-high-performance liquid chromatography. Peptides were separated onto a 150-µm inner diameter microcapillary trapping column, packed with ~2 cm of C18 Reprosil resin (5 µm, 100 Å, Dr. Maisch GmbH, Germany), followed by separation on a 50-cm analytical column (PharmaFluidics, Ghent, Belgium). Separation was achieved by applying a gradient from 5 to 27% acetonitrile in 0.1% formic acid for > 90 min at 200 nL $min^{-1}$. Electrospray ionization was enabled by applying a voltage of 2 kV using a home-made electrode junction at the end of the microcapillary column and sprayed from metal tips (PepSep, Denmark). MS survey scan was performed in the Orbitrap, in a range of 400–1800 m/z at a resolution of 60,000, followed by the selection of the 20 most intense ions (TOP20) for CID-MS2 fragmentation in the ion trap using a precursor isolation width window of 2 m/z, automatic gain control setting of 10,000, and a maximum ion accumulation of 100 ms. Singly charged ion species were not subjected to collision-induced dissociation fragmentation. Normalized collision energy was set to 35 V and an activation time of 10 ms. Ions within a 10 ppm m/z window around ions selected for MS2 were excluded from further selection for fragmentation for 60 s.

**RNA extraction and quantification**

Cell culture medium was rapidly removed from cells cultured on 150mm Petri dishes and 5 mL of Trizol reagent (Life Technologies) was added. After total RNA extraction, samples were treated with DNase using the Ambion DNA-free DNase Treatment kit (Life Technologies) according to the manufacturer's instructions, and resuspended in water. Samples were subsequently centrifuged at 13,000 rpm for 20 min at 4°C. Pellets were washed in 75% ethanol and resuspended in water. 1 µL RNA was diluted 1:100 in water and quality-checked using the Agilent RNA 6000 Pico kit

(Agilent technologies) according to the manufacturer's instructions, and ran on a 2100 Bioanalyzer Instrument (Agilent).

**RNA-seq library preparation and sequencing**

Libraries for RNA-seq were constructed using TruSeq Stranded Total RNA Library Prep Kit (Illumina), with the incorporation of dUTP in the second strand synthesis, and sequenced at the UMass Medical School. Sequencing was performed in paired-end reads (2 × 100 bp) using a HiSeq 4000 system (Illumina).

**Generation of CRISPR/Cas9 edited cell lines**

All transgenic iPSC cells in this study are stable expressing lines. To generate stable iPSC cell lines, cells were electroporated using the NEON transfection system and reagents (Life Technologies), according to the manufacturer's instructions. RNP complexes were generated using EnGen Spy Cas9 NLS (NEB, M0646T), in combination with a single sgRNA for AKAP8L (Hs.Cas9.AKAP8L.1.AA, IDT), and a mix of 2 sgRNAs for ANK2 (CD.Cas9.VYBY9746.AA and CD.Cas9.YTSN3051.AA, IDT). After 24 hr, cells were transferred to 90 mm gelatinized Petri dishes. Cells were grown for 10-12 days, with medium being changed every day. Individual iPSC colonies were picked and expanded for further screening.

**siRNA knock-down**

Silencer Select siRNA against DYRK1A were purchased from Thermo Scientific (4390824). Transfection into cells was performed on coverslips with RNAiMAX Transfection Reagent

(Invitrogen 13778100), according to the manufacturer's instructions. KD were assessed 48 hr after transfection.

##### Genomic DNA extraction

Cells from a confluent 150 mm gelatinized Petri dish were harvested and resuspended in 1-3 mL of lysis buffer (10 mM NaCl, 10 mM Tris-HCl pH 7.5, 10 mM EDTA-NaOH pH 8.0, 0.5% Sodium lauroyl sarcosinate) with proteinase K (Thermo Scientific EO0491) added to a final concentration of 200 µg $mL^{-1}$. Samples were incubated overnight at 55°C. 1/25 volume of 5 M NaCl and 2.5 volume of ice-cold 100% ethanol were added. After mixing, a visible white cloud of DNA was extracted using a bent pipette tip and transferred to a clean tube containing 1 mL 70% ethanol. DNA was pelleted (16,100 g, 5 min, 4°C) and air-dried. Subsequently the pellet was resuspended in 300-400 µL 10 mM Tris pH 8.5 and the concentration measured by Nanodrop.

#### Quantification and Statistical Analysis

##### RNA-seq data analysis

Starting with RNA-seq data in FASTQ format, we ran Trim Galore (*82*) (v0.6.7) to trim adapters and low-quality reads using "--stringency 7" and other default settings for Illumina paired-end reads. Sequence alignment was performed using HISAT2 (*83*) (v2.2.1) with the "--rna-strandness FR" setting; index files for the GRCh38 reference genome with transcripts were downloaded from the HISAT2 website (https://daehwankimlab.github.io/hisat2/download/). The htseq-count script in Htseq (*84*) (v0.13.5) was used to generate gene counts from aligned reads with "--stranded yes --minaqual 1" and other default settings; GRCh38 GTF gene annotation file was obtained from Ensembl (*85*) (release 84). The EdgeR package (*86*) (v3.34.0) was used to remove low-count genes (min.count=5 in filterByExpr function) before calculating normalized gene counts (method="TMM" in calcNormFactors function).

**scRNA-seq data t-SNE plots**

Single-cell RNA sequencing (scRNA-seq) data in Velmeshev *et al.* (*30*) were downloaded from the UCSC Cell Browser (https://cells.ucsc.edu/?ds=autism#). The gene expression matrix (exprMatrix.tsv.gz), cell meta annotations (meta.tsv), dimensionality reduction coordinates (tSNE.coords.tsv.gz), and the Seurat R package (*87*) (v3.2.0) were used to generate t-SNE plots showing either index gene expression or cell type annotations across cell clusters.

**Protein quantification from raw IP-MS data**

Software and methods used by each MS facility/site to generate protein-level quantification reports from raw IP-MS data are described below; **Supplementary Table 2** links each IP-MS dataset we analyzed to each facility.

*Broad:* Mass spectra were analyzed using Spectrum Mill (v7.0; https://proteomics.broadinstitute.org). For peptide identification, MS/MS spectra were searched against a sequence database for the UniProt human reference proteome, including isoforms, with a set of common laboratory contaminant proteins appended (2017: 65,068 entries). Search parameters included: ESI-QEXACTIVE-HCD scoring parameters, trypsin enzyme specificity with a maximum of two missed cleavages, 40% minimum matched peak intensity, ± 20 ppm precursor mass tolerance, ± 20 ppm product mass tolerance. Carbamidomethylation of cysteines and iTRAQ4 full labeling of lysines and peptide n-termini were set as fixed modifications.

Allowed variable modifications were oxidation of methionine (M), acetyl (ProtN-term), and deamidated (N), with a precursor MH+ shift range of -18 to 64 Da. Identities interpreted for individual spectra were automatically designated as valid by optimizing score and delta rank1-rank2 score thresholds separately for each precursor charge state in each LC-MS/MS while allowing a maximum target-decoy-based false discovery rate (FDR) of 1.0% at the spectrum level. Identified peptides were organized into protein groups and subgroups (isoforms and family members) with Spectrum Mill's subgroup specific option enabled, so that peptides shared between subgroups are ignored when using reporter ion intensities to perform protein-level quantitation.

*Whit*ehead: Mass spectra were analyzed using PEAKS Studio (v8.5; Bioinformatics Solutions). Peptide and protein identification was performed by searching against the UniProt human reference proteome, including isoforms (Swiss-Prot/TrEMBL, release 2019_01) together with a set of common contaminants. An FDR threshold of 1% was used for identification of peptides and proteins. Relative ratios of the iTRAQ 4-plex reporter ions were used for protein-level quantitation.

*CNCR:* Mass spectra were analyzed using MaxQuant (v1.6.1.0 or v1.6.3.4; https://www.maxquant.org). Peptide and protein identification was performed by searching against the UniProt human database, and label-free quantitation was performed to generate protein-level quantification reports. MaxQuant versions and settings used for each dataset can be found in the deposited parameters file linked to each dataset (**Supplementary Table 12**).

*Harvard:* Mass spectra were analyzed using Proteome Discoverer (v2.4; Thermo Scientific). Assignment of MS/MS spectra was performed using the Sequest HT algorithm by searching the data against the UniProt human reference proteome, including isoforms (Swiss-Prot, release 2019_01) as well as other known contaminants such as human keratins and common laboratory contaminants. Sequest HT searches were performed using a 10 ppm precursor ion tolerance and requiring each peptide's N/C termini to adhere with trypsin protease specificity, while allowing up to two missed cleavages. CID-MS2 spectra were searched with 0.5 Da ion tolerance for fragmentation. Methionine oxidation (+15.99492 Da) was set as variable modification. An MS2 spectra assignment FDR of 1% was applied to both proteins and peptides using the Percolator target-decoy database search. Label-free quantitation was performed to generate protein-level quantification reports.

**IP-MS data processing and analysis**

*Data processing:* Starting with protein-level quantification reports generated by the MS analytical software described above, we generally performed the following data processing procedures for each IP-MS dataset: (1) $\log_2$ transformation and median normalization of protein intensity values in each bait (i.e., index protein) or IgG control IP sample; (2) removing contaminants, proteins supported by < 1 unique peptide, and proteins detected in < 2 bait samples; (3) imputing missing protein intensity values in each sample by randomly sampling from a normal distribution with mean of $\mu - 1.8\sigma$ and standard deviation of $0.3\sigma$, where $\mu$ and $\sigma$ are the mean and standard deviation of the observed intensity values in the sample (*88*); (4) calculating $\log_2$ FC for each replicate pair of bait vs. IgG control samples. However, due to differences in MS protocols, we modified these steps for specific datasets: for data generated at Broad and Whitehead, few missing values were present due to the labeled quantification approach, therefore imputation was not performed and only proteins with no missingness across all bait and control samples were included for analysis;

for data generated at CNCR, instead of comparing bait samples against IgG controls to calculate $\log_2$ FC, the median of several negative IPs (i.e., bait samples in which the bait protein itself was not detected; **Supplementary Table 12**) from the same MS run were used as controls.

*Genoppi analysis:* We analyzed each processed IP-MS dataset using the Genoppi R package (*24*) (v1.0), including: (1) calculating Pearson's correlation of $\log_2$ FC values between replicates; (2) calculating average $\log_2$ FC, and corresponding P-value and false discovery rate (FDR), for each protein across replicates using a one-sample moderated t-test (*89*); (3) identifying statistically significant ($\log_2$ FC $> 0$ and FDR $\leq 0.1$) interactors of the bait; (4) assessing overlap enrichment between the identified interactors and known interactors from the InWeb database (*23*), as curated in the Genoppi R package. We QC'ed each dataset using two criteria: the replicate $\log_2$ FC correlation must be $> 0.6$ and the bait protein itself must be significant ($\log_2$ FC $> 0$ and FDR $\leq$ 0.1); datasets that failed to meet these criteria were excluded from further analysis. All IP-MS analysis results are provided in **Supplementary Table 2**.

*Defining non-interactors:* For each QC'ed IP-MS dataset derived from iNs, besides identifying significant interactors as described above, we also defined a matching set of "non-interactors" (**Supplementary Table 3**) to serve as proxy for the background iN proteome in conditional enrichment analyses. We applied different definitions for datasets generated by the labeled vs. label-free quantification approaches: for labeled datasets generated at Broad and Whitehead, we considered all non-significant proteins (i.e., $\log_2$ FC $\leq 0$ or FDR $> 0.1$) in Genoppi analysis to be non-interactors; for label-free datasets generated at CNCR, we included both non-significant proteins in Genoppi analysis and other proteins in the MS quantification report that were detected in < 2 bait samples (which were filtered out prior to Genoppi analysis).

*Generating PPI networks:* We consolidated all QC'ed iN-derived IP-MS datasets into a combined PPI network; we also combined the datasets for the same index proteins into index protein-specific networks. The interactors in each network are proteins that show up as significant interactors in ≥ 1 source IP-MS datasets contributing to the network; the matching "non-interactors" are proteins that show up as non-interactors in ≥ 1 source dataset but never as interactors in any source dataset. The full lists of interactors and non-interactors associated with each PPI network are provided in **Supplementary Table 3**; we note that in this table, we removed all index proteins from both the interactor and non-interactor lists for the combined PPI network to avoid biasing downstream enrichment analyses.

**Co-expression analysis**

Pairwise co-expression between each index gene and all other protein-coding genes were estimated using data from four independent studies, including (1) Stickels *et al.* (*36*): spatial transcriptomics in mouse neocortex (Puck_190921_19.digital_expression.txt.gz retrieved from: https://singlecell.broadinstitute.org/single_cell/study/SCP815/highly-sensitive-spatial-transcriptomics-at-near-cellular-resolution-with-slide-seqv2#study-download); (2) Maynard *et al.* (*35*): spatial transcriptomics in human dorsolateral prefrontal cortex (count matrix retrieved from spatialLIBD R package: https://github.com/LieberInstitute/HumanPilot); (3) Velmeshev *et al.* (*30*): single-cell RNA-seq in human cortex (rawMatrix.zip retrieved from: https://cells.ucsc.edu/?ds=autism); and (4) BrainSpan: bulk RNA-seq across human brain regions and developmental stages ("RNA-Seq Gencode v10 summarized to genes" dataset retrieved from: https://www.brainspan.org/static/download.html). We used two different methods for estimating co-expression to account for different statistical properties in these datasets. For the spatial transcriptomic datasets with very sparse gene expression matrices, we reasoned that the binary presence/absence of genes across physical locations would be the most informative, and therefore performed one-tailed Fisher's exact tests to calculate the significance of co-occurrence for each gene pair across locations. For the other datasets, we calculated a proportionality metric, ρ, for each gene pair using the propr R package (*90*) (v4.2.6); this metric is analogous to conventional correlation measures but has been shown to be better at capturing functional associations between genes in RNA-seq data (*91*). After calculating either the Fisher's exact P-values or the proportionality ρ values for all gene pairs involving each index gene, we then performed rank-based inverse normal transformation to convert the values into co-expression Z-scores (where a positive score indicates that a gene has higher than average co-expression with the index gene compared to the rest of the genome). We performed two-tailed Wilcoxon rank-sum tests to assess if the co-expression Z-scores between index genes and their interactors are significantly different from the scores between the index genes and other gene groups including non-interactors detected in IP-MS, known interactors from InWeb, and all protein-coding genes.

**BrainSpan analysis**

Gene expression in four regions of the frontal cortex (dorsolateral prefrontal cortex, medial prefrontal cortex, ventrolateral prefrontal cortex, orbital frontal cortex) across 10 developmental stages were obtained from the BrainSpan exon microarray dataset (*34*) (www.brainspan.org). At each developmental stage, we calculated the median and standard error of the expression values for various gene sets, limiting to genes with available BrainSpan data: (1) ASD risk genes reaching FWER ≤ 0.05, FDR ≤ 0.1, and FDR ≤ 0.25 in Satterstrom *et al.* (*10*); (2) index and interactor genes in our combined network; (3) SCN2A, SHANK3, and SYNGAP1 interactors found only in iNs, only in brain homogenates, and shared by both sample types; (4) overlap between the FDR ≤ 0.25 genes from Satterstrom *et al.* and interactor genes in the combined network; (5) random genes sampled from the BrainSpan dataset for comparison against the other gene sets.

**Tissue enrichment analyses using GTEx data**

Gene tissue specificity scores in all (GTEx.tstat.tsv) and brain region (GTEx_brain.tstat.tsv) GTEx tissues, as described in Finucane *et al.* (*37*), were downloaded from: https://data.broadinstitute.org/alkesgroup/LDSCORE/LDSC_SEG_ldscores/tstats/. Following the definition in Finucane *et al.*, we defined tissue-specific genes as the 10% of genes with the highest scores in each tissue. One-tailed hypergeometric tests were performed to assess the overlap enrichment between interactors in the combined PPI network vs. the tissue-specific genes in all tissues or the brain region tissues. In each test, the "population" was defined as all genes found in the GTEx data; within this population, "success" was the interactors in the network, "sample" was the tissue-specific genes, and "success in sample" was the overlap.

**Cell type enrichment analyses using scRNA-seq data**

We downloaded scRNA-seq data reported in Velmeshev *et al.* (*30*) from the UCSC Cell Browser (https://cells.ucsc.edu/?ds=autism). To define "commonly expressed genes" in each of the 17 annotated cell types, we used the gene expression matrix (exprMatrix.tsv.gz) and the cell meta annotations (meta.tsv) to identify genes that have non-zero expression (i.e., $\log_2$(UMI count) > 0) in > 50% of cells in a cell type. Separately, we obtained differentially expressed genes (DEGs) between ASD patients vs. controls in each cell type from Data S4 of Velmeshev *et al*. One-tailed hypergeometric tests were performed to assess the overlap enrichment between interactors in the combined PPI network vs. either the commonly expressed genes or the DEGs in each cell type. In each test, the "population" was defined as either all genes found in the scRNA-seq dataset (for the global enrichment tests) or the subset of genes that are interactors or non-interactors in the network (for the conditional enrichment tests). Within this population, "success" was the interactors in the network, "sample" was the commonly expressed genes or the DEGs in a cell type, and "success in sample" was the overlap.

**SynGO analysis**

We performed SynGO gene set enrichment analysis for genes in the combined PPI network by inputting their gene symbols into the SynGO browser (*38*) (https://syngoportal.org; dataset release: 20180731). The SynGO "Biological Processes" annotations and the "brain expressed" background set were used in the analysis.

**GO analysis for ANK2 WT- and KO-specific interactors**

We identified ANK2 "WT-specific" and "KO-specific" interactors by analyzing ANK2 IP-MS experiments performed in wild-type (WT) and giant ANK2 knockout (KO) neural progenitor cells. Specifically, the WT-specific interactors are proteins that are significant ($\log_2$ FC > 0 and FDR ≤ 0.1) in the ANK2 IP in WT cells (compared to IgG controls) but not in the ANK2 IP in KO cells; and vice versa for the KO-specific interactors. We performed GO term enrichment analysis using the GO cellular component (CC) annotations provided in the Genoppi R package (*24*) (v1.0), which were originally downloaded from the Gene Ontology Consortium (*41*, *42*) (2020-03-23 release; http://current.geneontology.org/products/pages/downloads.html). One-tailed hypergeometric tests were performed to assess the overrepresentation of CC terms among the WT- or KO-specific interactors. In each test, the "population" contained genes detected in ≥ 2 ANK2 IP replicates in WT cells or ≥ 2 ANK2 IP replicates in KO cells. Within this population, "success" was the genes annotated with a CC term, "sample" was the WT- or KO-specific interactors, and "success in sample" was the overlap.

**Enrichment analyses for IGF2BP1-3 targets**

RNA targets of IGF2BP1, IGF2BP2, and IGF2BP3 were obtained from Supplementary Table 1 of Huang *et* al. (*48*); these targets were identified by both RIP and PAR-CLIP methods in HEK293T cells. First, we performed one-tailed hypergeometric tests to assess the overlap enrichment between target genes of each IGF2BP or their combined target list vs. the ASD risk genes reaching FDR ≤ 0.1 in Satterstrom *et al.* (*10*). In each test, the "population" was defined as all autosomal genes in Table S2 of Satterstrom *et al.*; within this population, "success" was the target genes, "sample" was the ASD risk genes, and "success in sample" was the overlap. Second, to test whether the IGF2BP1-3 targets are enriched for polygenic risk of ASDs, we performed a global MAGMA analysis as described in **Common variant enrichment analysis**, except the "gene sets" tested here were the IGF2BP1-3 targets instead of the PPI network genes. Finally, we performed another set of one-tailed hypergeometric tests to assess the overlap enrichment between the IGF1BP1-3 targets vs. interactors in the combined or index protein-specific PPI networks. In each test, the "population" contained genes that are interactors or non-interactors in a network; within this population, "success" was the target genes, "sample" was the interactors in the network, and "success in sample" was the overlap.

**Rare variant enrichment analysis**

Gene-based rare variant association scores were extracted from exome sequencing studies of ASD ("qval_dnccPTV" column in Table S2 of Satterstrom *et al.* (*10*)), DD ("denovoWEST_p_full" column in Table S2 of Kaplanis *et al.* (*54*)), and SCZ ("P meta" column in Table S5 of Singh *et al.* (*55*)). In the global enrichment analysis, for each PPI network and each trait, a one-tailed Kolmorgov-Smirnov test was performed to compare the score distribution of the interactor genes against the rest of the genome (with available score data), testing the alternative hypothesis that the cumulative distribution function of the interactors lies above that of other genes (i.e., the interactors have smaller/more significant scores). In the conditional enrichment analysis, an analogous one-tailed Kolmorgov-Smirnov test was performed to compare the scores of the interactors vs. the non-interactors associated with each PPI network.

**pLI score enrichment analysis**

Gene pLI scores were obtained from the gnomAD (*56*) (v2.1.1) "pLoF Metrics by Gene TSV" dataset. For each PPI network, a one-tailed Kolmorgov-Smirnov test was performed to compare the pLI scores of the interactor genes against other genes in the pLI dataset (for the global enrichment analysis) or against the non-interactors (for the conditional enrichment analysis), testing the alternative hypothesis that the cumulative distribution function of the interactors lies below that of other genes (i.e., the interactors have higher pLI scores).

**Common variant enrichment analysis**

GWAS summary statistics for ASD (*49*), ADHD (*57*), BIP (*58*), MDD (*59*), and SCZ (*60*) were downloaded from the Psychiatric Genomics Consortium (www.med.unc.edu/pgc/download-results/). GWAS summary statistics for height (*61*) were downloaded from the GIANT consortium (https://portals.broadinstitute.org/collaboration/giant/index.php/GIANT_consortium_data_files). All of these GWAS were conducted using individuals of European (EUR) ancestry. The following analyses were performed using MAGMA (*50*) (v1.09) to calculate the polygenic risk enrichment of each PPI network for each GWAS phenotype. First, variants in the 1000 Genomes (*92*) (phase 3) EUR panel were mapped to protein-coding genes in the Ensembl (*85*) GRCh37 database with a flanking window of ± 50 kb; variants in the major histocompatibility complex region (chr6:28.5M-33.4M) were excluded. Next, for each GWAS dataset, gene-based P-values were calculated using the SNP-wise Mean model and linkage disequilibrium information estimated from the 1000 Genomes EUR panel. Finally, for each GWAS dataset and each PPI network, the gene-set analysis model in MAGMA was used to compare the interactor genes in the network against the rest of the protein-coding genome (for the global enrichment tests) or the non-interactor genes (for the conditional enrichment tests), computing a one-tailed P-value to support the alternative hypothesis that the interactors are more strongly associated with the GWAS phenotype.

**Social Manhattan plot**

ASD risk genes with FDR ≤ 0.25 were obtained from Table S2 of Satterstrom *et al.* (*10*), along with their GRCh38 genomic positions and Q-values. The combined PPI network was intersected with the ASD gene list to identify the subset of genes that are either an index gene or an interactor in the network. The genomic positions and -$\log_{10}$ Q-values of these genes were plotted in a

Manhattan plot, with visual links between the genes to indicate PPIs in the network. For visualization, the minimum Q-value was set to 1e-16 in the plot.

**Supplementary Text 1. Factors contributing to the large number of novel interactions in our IP-MS data.**

Across the 26 IP-MS datasets we analyzed, > 90% of the protein interactions have not been reported in InWeb. The large percentage of novel interactions can readily be explained by a combination of the brain cell-type-specificity of the data we report here and several additional deliberate design choices in our experimental protocol (which substantially differs from some common experimental approaches underlying data in InWeb). The design choices in question include: i) immunoprecipitation of endogenous [i.e., non-tagged and non-overexpressed] protein isoforms expressed in neurons; ii) IPs performed in a salt/detergent environment optimized for identifying inclusive protein networks instead of narrowly defined core protein complexes; and iii) IPs performed exclusively in neurons and not other 'generic' cell types. In more detail:

1. We performed IPs of endogenously expressed proteins using commercially available immunoreagents to avoid artifacts associated with tagged overexpression systems (e.g., due to excess of tagged proteins, biochemical interference of the tag with the interactions of the bait protein, or arbitrary selection of a specific protein isoform for tagging and overexpressing). This design choice allowed us to study relevant proteins in neurons, which are difficult-to-transfect cell types, without biases for previously annotated isoforms. Additionally, we did not perform any cross-linking prior to immunoprecipitation, which usually drives enrichment of the 'bait' protein.

2. Our experimental strategy was also designed for identifying protein networks not limited to stringently defined core protein complexes. This means that we are deliberately including both first- and second-order interactions in our data. We achieved this by titrating detergent

composition during co-IP washes (NP-40 is 0.01% v/v) and by maintaining low salt concentrations (150mM) to preserve electrostatic interactions. Indeed, it was not our aim to resolve stringent binary interactions or core molecular complexes, which is the scope of many large-scale protein interaction screens in the literature that constitute a significant fraction of the InWeb dataset. These studies, for the most part, use yeast two-hybrid assays and pull-downs of tagged proteins in overexpression systems. Rather, our aims were to map the brain cell-type-specific cellular networks anchored in index proteins linked to ASDs, use these inclusive datasets to model pathways and networks in the disease, and integrate with genetic datasets that are otherwise difficult to interpret mechanistically.

3. To further illustrate the importance of cell-type-specificity in our approach, we compared the interactors identified in our study vs. those already published in InWeb in more detail (see table below). In general, we found a lower percentage of overlap with InWeb for index proteins that have more neuron-specific functions and expression (e.g., 0.74% for SYNGAP1), and a higher percentage of overlap for index proteins with more ubiquitous expression across tissues and cell types (e.g., 54.2% for CTNNB1). This indicates that most of the 'novel' interactions in our data are driven by neuronal genes that are underrepresented in InWeb, which is generally enriched for genes that are more amenable to biochemical assays in proliferative, non-neuronal cell models (e.g., HEK cells). These observations are in agreement with results from our previously published Genoppi paper (*24*), in which we applied a similar IP-MS protocol to identify interactors for four unrelated proteins (BCL2, TDP-43, MDM2, and PTEN) across four neuronal or cancer cell lines: we found that only ~17% of the interactions were reported in InWeb and that they clustered according to the origin of the cell line used in the IPs, with a

clear set of cell-type-specific interaction partners both in neurons and in various cancer cell lines.

| Gene | # of interactors in this study | # of interactors in InWeb | # in overlap | overlap/this study % | overlap/InWeb % |
|---|---|---|---|---|---|
| ADNP | 12 | 53 | 1 | 8.33 | 1.89 |
| ***ANK2**** | ***108*** | ***119*** | ***8*** | ***7.41*** | ***6.72*** |
| ***ARID1B**** | ***45*** | ***89*** | ***13*** | ***28.89*** | ***14.61*** |
| CHD8 | 83 | 129 | 1 | 1.20 | 0.78 |
| ***CTNNB1**** | ***24*** | ***755*** | ***13*** | ***54.17*** | ***1.72*** |
| ***DYRK1A**** | ***604*** | ***358*** | ***39*** | ***6.46*** | ***10.89*** |
| GIGYF1 | 36 | 20 | 0 | 0.00 | 0.00 |
| ***MED13L**** | ***45*** | ***172*** | ***11*** | ***24.44*** | ***6.40*** |
| PTEN | 3 | 272 | 1 | 33.33 | 0.37 |
| SCN2A | 58 | 17 | 1 | 1.72 | 5.88 |
| SHANK3 | 129 | 71 | 3 | 2.33 | 4.23 |
| SYNGAP1 | 136 | 36 | 1 | 0.74 | 2.78 |
| TLK2 | 60 | 26 | 1 | 1.67 | 3.85 |

* Bold italic entries marked by an asterisk indicate nominally significant ($P < 0.05$) overlap between interactors in this study vs. InWeb

Finally, we also assessed whether the computational approach (Genoppi/limma) we used to analyze the IP-MS data contributes significantly to the large portion of novel interactions we identified. In the Genoppi paper, we delved into this issue by analyzing the IP-MS data using both Genoppi and an alternative established method, SAINTexpress (*93*), and found that on average ~85% of the interactors identified by Genoppi were also identified by SAINTexpress, indicating strong agreement between the two methods. This shows that our specific computational method for analyzing the data is not driving the number of new interactions reported in this study.

In conclusion, most newly reported interactions in the current study likely result from: i) an experimental design that aims at capturing endogenous protein interactions; ii) a deliberately inclusive IP protocol to identify neuronal networks and pathways rather than exclusively stringent core molecular complexes [which is the data type typically reported in InWeb]; and iii) an experimental system to capture brain cell-type-specific protein interactions that have not been studied in previous literature.

**Supplementary Text 2. Comparison of MS protocols and IP-MS datasets generated across MS sites.**

All MS facilities that generated data for this study employ a bottom–up approach whereby proteins are identified from peptides upon enzymatic digestion of the samples and subsequent comparison against standard databases of human proteins. However, there are several well-documented and facility-specific technical variables in the sample preparation and downstream workflow for MS that could impact the structure of the final data and the population of proteins detected by MS (*94*). Among these are i) removal of contaminants; ii) digestion strategies; iii) post-labeling; and iv) fractionation and separation of digested samples. As detailed in **Material and Methods**, each MS facility employed a different combination of these variables, yielding different, yet highly complementary, datasets. Therefore, we reasoned that by jointly analyzing these datasets to build an inclusive PPI network, we could capture complementary biological signals that would not be possible using a single MS platform due to the technical variables employed by each platform.

Indeed, each facility analyzed their raw MS spectra using in-house pipelines, software, and settings that have been optimized and tested for their specific workflow. Therefore we decided to use the protein-level quantification reports from each facility as the starting point of our analysis, as data in these reports have already been processed into similar formats regardless of differences in the upstream MS pipeline. We streamlined the downstream data processing and analysis pipeline as much as possible, and were able to identify PPIs that are consistent across a variety of metrics regardless of facility and protocol (see below). We note that this scenario is not uncommon for researchers who work with external facilities or with published datasets, and we hope our approach can serve as a reference for others looking to aggregate data generated from different MS methods.

We have performed several types of analyses to compare IP-MS data generated across facilities to support that our approach to integrating data across facilities is sound:

1. First, we compared various summary statistics of the 26 IP-MS datasets that were used to generate our combined iN-derived PPI network, including the $log_2$ FC correlation between replicates, number of analyzed proteins and significant interactors, $log_2$ FC and FDR of the index protein, and the same statistics for the interactors (**Supplementary Fig. 3A**). Most statistics are comparable across facilities, with a few exceptions: the Broad facility detected more proteins compared to the other two sites, while the CNCR facility consistently reported larger $log_2$ FC values due to using a label-free quantification approach.

2. Second, we compared all analyzed proteins and significant interactors derived from IPs for the same index protein, resulting in 17 pairwise comparisons involving IPs of 9 index proteins (**Supplementary Table 4** and **Supplementary Fig. 3B**). As expected, we observed that the 7 IP pairs from the same facility have better overlap between analyzed proteins (median overlap % = 59.6%) compared to the 10 IP pairs from different facilities (median overlap % = 36.2%). Among proteins analyzed in both IPs, there is significant $log_2$ FC correlation between all IP pairs, although the correlations for pairs from the same facility (median correlation = 0.798) are higher than pairs from different facilities (median correlation = 0.560). Similarly, IP pairs from the same facility also have better overlap between identified interactors (median overlap % = 76.8%) compared to those from different facilities (median overlap % = 51.9%).

These results not only reflect the methodological differences between facilities, but also suggest that proteins detected in each IP (i.e., technical variance between identical experiments) is a major factor driving the differences in interactors identified across IPs (even for IPs repeated at the same facility). To further assess whether the data structure and protein detection differences across facilities and IPs influence the quality and biological validity of the interactions identified in our data, we used both experimental and computational approaches to compare the interactors identified by different facilities:

1. First, we compared the validation rates of a subset of the interactors in forward and reverse IP experiments followed by western blotting (IP-WB; **Supplementary Table 5**). Overall, interactors identified by all three facilities have close to ~90% validation rates in forward IP-WB experiments (see table below), which agree with the 10% FDR cutoff we applied to define significant interactors. The validation rates in reverse IP-WB experiments are more variable across facilities, ranging from 66.7% for CNCR to 100% for Whitehead. However, given the relatively small sample size (23 reverse IPs in total) connatured with biochemical validations, these differences are within reasonable expectations. Combining results across both forward and reverse IP-WB, we observed >80% validation rate for all three facilities, with Whitehead having the best rate (96.3%), followed by CNCR (87.2%) and Broad (83.3%).

| **Facility** | **Forward validation rate** | **Reverse validation rate** | **Combined validation rate** |
|---|---|---|---|
| Broad | 21/24 (87.5%) | 11/13 (84.6%) | 25/30 (83.3%) |
| Whitehead | 23/24 (95.8%) | 5/5 (100%) | 26/27 (96.3%) |
| CNCR | 31/34 (91.2%) | 6/9 (66.7%) | 34/39 (87.2%) |
| Combined | 65/71 (91.5%) | 19/23 (82.6%) | 72/81 (88.9%) |

2. When we calculated the overlap enrichment between index protein interactors in each IP and known interactors in InWeb, we observed no significant difference between the facilities (**Supplementary Fig. 3A**). In terms of the percentages of 'novel' interactions identified by each facility (i.e., interactions not found in InWeb nor in the BioPlex 3.0 HEK293T dataset (*95*)), CNCR has the smallest percentage (89.5%, or 375/419), followed by Broad (94.5%, or 715/757) and Whitehead (95.8%, or 230/240).

3. We also explored the co-expression patterns of index protein-interactor gene pairs identified by the three facilities across four independent brain transcriptomic datasets (**Supplementary Fig. 3C**). We found that all three sets of gene pairs show comparable co-expression; that is, no set consistently has higher co-expression scores compared to the other sets. Importantly, interactors identified by all facilities are generally more likely to be co-expressed with the index proteins compared to all protein-coding genes, and in some cases to InWeb interactors.

Overall, our findings indicate that there are nuanced differences in terms of data structures and proteins detected between the IP-MS datasets generated at the three facilities, likely due upstream MS protocol and/or analytical differences. However, the significant PPIs that we identified using our downstream analytical pipeline have similar quality and degree of biological relevance regardless of facility origin, suggesting these data are complementary to each other.

Besides subsetting interactions by facility, we also grouped them by the number of IP datasets that identified each interaction to assess whether 'recurrent' interactions in our data (i.e., identified in >1 IP) may be more reproducible or biologically relevant compared to interactions that were only identified in a single IP:

1. In our forward and reverse IP-WB validation experiments, the subset of interactions detected in only 1 vs. in multiple IPs have very similar validation rates (see table below). We also extended the concept of 'recurrence' to look at interactions that have been previously reported in InWeb or BioPlex, and found 'novel' interactions in our data to have robust validation rates of ~90% that are comparable to the 'recurrent' interactions reported in previous literature. Together, these results demonstrate that interactions identified in only 1 IP in our data have comparable reproducibility in IP-WB validations as interactions that were detected in multiple IPs or previously reported interactions.

| **Interaction type** | **Forward validation rate** | **Reverse validation rate** | **Combined validation rate** |
|---|---|---|---|
| Found in 1 IP | 43/47 (91.5%) | 13/16 (81.3%) | 46/52 (88.5%) |
| Found in >1 IP | 22/24 (91.7%) | 6/7 (85.7%) | 26/29 (89.7%) |
| InWeb | 16/17 (94.1%) | 3/3 (100%) | 16/17 (94.1%) |
| non-InWeb, 1 IP | 34/37 (91.9%) | 11/14 (78.6%) | 37/42 (88.1%) |
| non-InWeb, >1 IP | 15/17 (88.2%) | 5/6 (83/3%) | 19/22 (86.4%) |
| BioPlex | 7/8 (87.5%) | 2/2 (100%) | 7/8 (87.5%) |
| non-BioPlex, 1 IP | 39/42 (92.9%) | 12/15 (80%) | 42/47 (89.4%) |
| non-BioPlex, >1 IP | 19/21 (90.5%) | 5/6 (83.3%) | 23/26 (88.5%) |

2. Related to above, we also calculated the percentages of 'novel' (i.e., non-InWeb and non-BioPlex) interactions identified in only 1 IP vs. in multiple IPs. We found that 93.6% (1103/1178) of interactions identified in only 1 IP are novel, compared to 87.6% (134/153) and 100% (12/12) for those found in 2 or 3 IPs. Therefore, the most recurrent interactions

identified in our data are actually all novel interactions that have not been reported in previous literature.

3. We plotted the co-expression scores of index protein-interactor gene pairs identified in 1, 2, or 3 IPs across brain transcriptomic datasets and found these gene pairs to show comparable co-expression across three out of the four tested datasets (**Supplementary Fig. 3D**). The only exception is the BrainSpan dataset, in which gene pairs identified in only 1 IP actually have higher co-expression than those identified in 2 or 3 IPs. Again, these results suggest that interactions identified in only 1 vs. multiple IPs are both biologically relevant in terms of capturing gene relationships in complex brain tissues.

4. Finally, for SCN2A, SHANK3, and SYNGAP1 interactors, we compared their overlap with interactors detected in *postmortem* brain homogenates (**Fig. 2G** and **Supplementary Fig. 6C**). Both iN interactors detected in only 1 vs. in > 1 IP show significant overlap with those detected in brain samples (see table below). However, iN interactors identified in > 1 IP have more significant overlap (*P* = 7.9E-14) compared to those identified in only 1 IP (*P* = 3.4E-3). This trend may be due the fact that the 'strongest' interactions are more likely to be detected both across IPs performed at different MS sites and in more heterogeneous brain samples that consist of a mix of cell types, while 'looser' interactions (that are still biologically valid and important to ASDs) may only be detectable in more homogeneous cell populations like the iNs. However, the other analyses described above indicate that both types of interactions are reproducible and biologically relevant. In the future, additional IPs performed using tissues from different brain regions may allow us to further investigate this issue.

| **Interactor type** | **# in iN only** | **# in brain only** | **# in overlap** | **Overlap *P**** |
|---|---|---|---|---|

| | | | | |
|---|---|---|---|---|
| SCN2A, 1 iN IP | 52 | 22 | 1 | 0.23 |
| SCN2A, >1 iN IP | 5 | 23 | 0 | 1 |
| SHANK3, 1 iN IP | 74 | 36 | 11 | 4.1E-10 |
| SHANK3, >1 iN IP | 25 | 28 | 19 | 3.6E-21 |
| SYNGAP1, 1 iN IP | 100 | 27 | 5 | 2.1E-3 |
| SYNGAP1, >1 iN IP | 26 | 27 | 5 | 1.2E-5 |
| Combined, 1 iN IP | 210 | 81 | 19 | 3.4E-3 |
| Combined, >1 iN IP | 53 | 76 | 24 | 7.9E-14 |

*Overlap enrichment P-values were calculated using one-tailed hypergeometric tests and proteins detected in both iN and brain as the total population

In conclusion, in both the experimental and computational analyses described above, we did not observe consistent quality differences between i) interactors identified by different MS facilities and ii) interactors identified in only 1 vs. multiple IPs. That is, we did not find clear evidence suggesting that the more inclusive dataset of the ‘union’ of interactors across IPs would be inherently more ‘noisy’ than a more restricted set of interactors identified by looking at the intersection of interactors across IPs. Therefore, since many of our downstream analyses rely on having large numbers of network genes to perform well-powered enrichment tests, we made a design choice to study the most inclusive network for each protein by taking the union of all of its interactors across IPs. However, we note that we also provide all the necessary data (**Supplementary Tables 2** and **3**) for other researchers to explore the ‘intersection’ (or other approaches to defining subsets) of interactors based on their specific research goals.

**Supplementary Text 3. Discussion on the $\log_2$ FC cutoff used to define index protein interactors.**

In proteomic data, the distribution of observed fold changes (FCs) is directly dependent on the experimental design in a given study. That is, FC is a relative, context-dependent measure and often not directly comparable between studies. For example, as discussed in **Supplementary Text 1**, many previous interaction proteomics studies have aimed at identifying core molecular complexes through tagging and overexpressing proteins for immunoprecipitation, mostly in "generic" cell types that are easy to transfect. In these studies, it may be more common to detect interactions at high FCs (>2) due to overexpression of proteins (*96*). In contrast, we purposely designed our IP experiments to pull down endogenously expressed protein isoforms in neurons, some of which may have relatively low expression despite their functional importance (e.g., SYNGAP1 (*97*)); we also titrated our IP conditions to preserve larger and more inclusive networks of interacting proteins rather than restricting to narrowly defined core protein complexes that capture more limited biological insights. These design choices directly impact the measured protein abundances and FCs observed in our experiments and may result in relatively low FCs. In addition, differences in MS methods would also be a major factor determining the FC observed in the experiment. For example, data generated using isobaric labeling tend to have lower FCs due to ratio compression compared to those generated with a label-free approach. This is indeed what we observed in this study, where interactors identified using isobaric labeling MS have a median $\log_2$ FC of ~1, compared to a median of ~5 for those from label-free data (**Supplementary Fig. 3A**).

Therefore, we chose not to use an arbitrary FC cutoff when defining significant interactions as it would not be consistent across studies and protocols. Instead, we defined all proteins with $\log_2$ FC $> 0$ and FDR $\leq 0.1$ as significant interactors of the index protein in each IP-MS experiment.

However, we note that the statistical method that we used to calculate significance (i.e., the moderated t-test implemented in the Genoppi/limma R package) is inherently designed so that proteins with larger FC (and thus larger sample variance) are more likely to be significant compared to proteins with smaller FC (*98*). Similar concepts were implemented in several other established approaches for mapping PPIs that implicitly consider FC information, but do not specifically use an FC cutoff (e.g., ComPASS-Plus used in BioPlex (*99*)).

In addition, we performed validation experiments and computational analyses to support the high quality and biological relevance of the identified interactors regardless of their FCs:

1. **Reverse IP experiments validated interactions across a wide range of FCs.** To assess the quality and reproducibility of our PPI data, we have performed a significant amount of validation experiments including both forward and reverse IPs, testing interactions identified with $\log_2$ FCs of 0.31 to 18.1 (**Fig. 2F** and **Supplementary Table 5**). In particular, we performed reverse IPs using a panel of interactors as baits, and successfully detected the original index proteins in 19 out of 23 reverse IPs that showed bait enrichment (82.6% validation rate). Given that we do not expect all true interactions to be detectable bidirectionally, these results support a relatively low false positive rate (~17%) in our data that is in general agreement with the 10% FDR cutoff we applied to identify significant interactions. We also specifically tested whether there was any association between FCs in the original IP-MS data and validation outcomes in the reverse IPs and did not observe any significant relationship; in fact, we were able to validate interactors identified at $\log_2$ FCs of 0.54 to 13.2 (**Supplementary Table 5**).

2. **Additional validation using known protein interaction data.** We found that interactors for five of the 13 index proteins are enriched for known InWeb interactors, including some gold standard interactors that have relatively low FCs in our data (**Supplementary Tables 2** and **3**). For example, in the ANK2 IP highlighted in **Supplementary Fig. 2B**, eight known InWeb interactors were identified as significant interactors in our data with $\log_2$ FCs of ~0.5 to 2. The relatively low FCs here are certainly not reflective of these interactors being false positives, rather they reflect our experimental design and the isobaric labeling MS protocol used to generate the data.

3. **Orthogonal support using brain co-expression data.** We used human and mouse brain co-expression data to show that on average, transcripts of the interactors are more likely to be co-expressed (using bulk or single-cell RNA-seq data) or co-localized (using spatial transcriptomic data) with that of the index proteins compared to i) the non-interactors [i.e., non-significant proteins with $\log_2$ FC $\leq 0$ or FDR $> 0.1$ in IP-MS]; ii) known interactors in InWeb; and iii) all protein-coding genes (**Fig. 2H** and **Supplementary Fig. 7C**). Another key observation is that when we grouped interactors based on different $\log_2$ FC thresholds, we did not observe a positive association between their FCs and co-expression scores in any of the four tested expression datasets (**Supplementary Fig. 3E**). This suggests that the interactions identified at lower FCs could have as much biological relevance as those identified at higher FCs in terms of representing pairwise functional gene relationships in complex brain tissues.

In conclusion, FCs are strongly dependent on the experimental design and our deliberate choices in this study would not result in very high FC values. We also note that, since FCs are context-dependent, there is no consensus in the literature about what a relevant or appropriate global FC cutoff would be in IP-MS experiments. However, the experimental and computational validations that we have described above strongly support the high quality of our data despite using an inclusive FC cutoff to define interactors across our experiments. Furthermore, our results indicate that there are no obvious quality differences between the interactors we identified across a range of different FCs.

#### Supplementary Figures

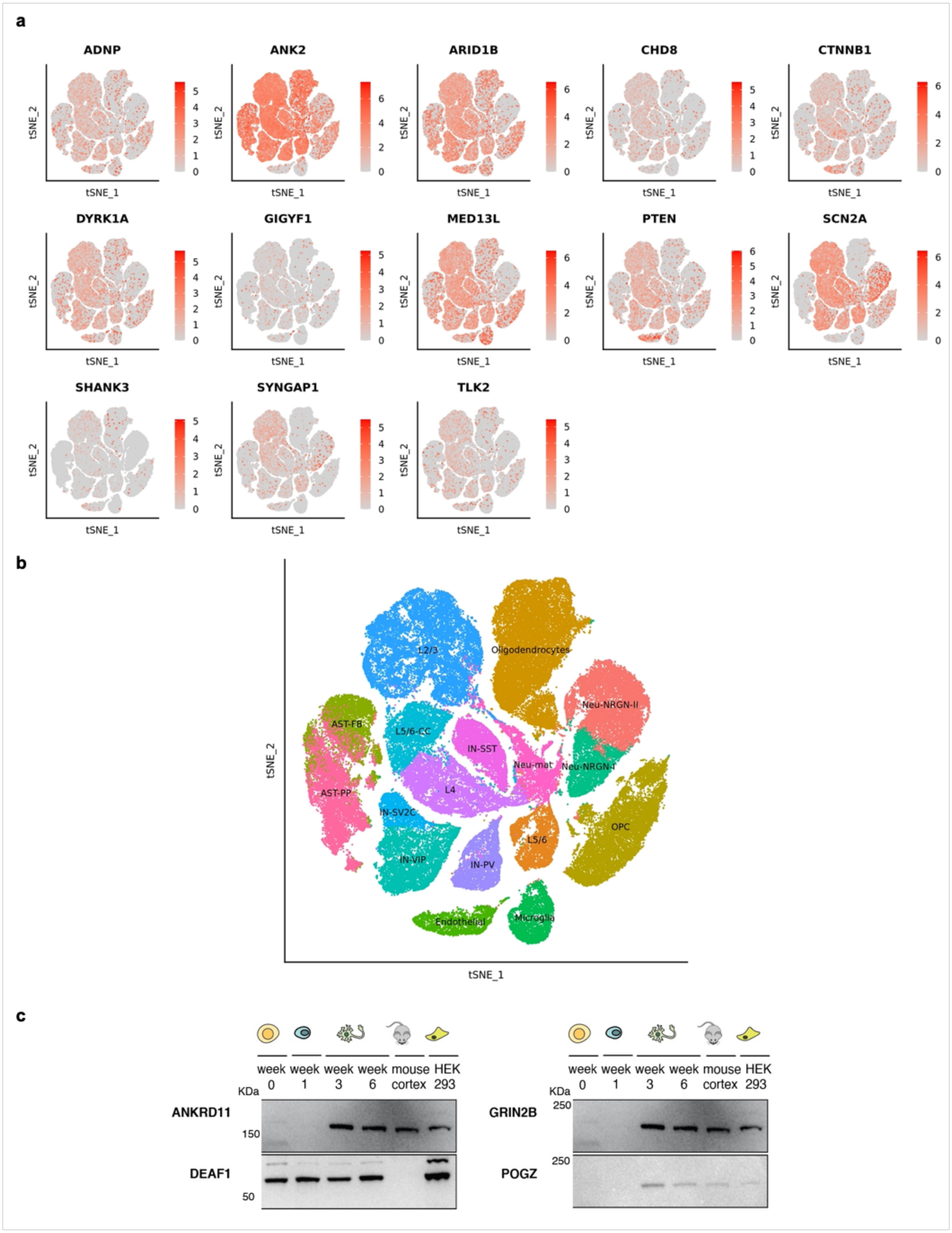

**Supplementary Figure 1. Expression of ASD index genes and proteins. (a)** t-SNE plots showing gene expression in *post mortem* cortex of ASD patients and controls, for the 13 index genes with successful IP experiments in this study. **(b)** Cell type annotations for the t-SNE plots shown in (a). Cell type abbreviations: AST-FB and AST-PP, fibrous and protoplasmic astrocytes; OPC, oligodendrocyte precursor cells; IN-PV, IN-SST, IN-SV2C, and IN-VIP, parvalbumin, somatostatin, SV2C, and VIP interneurons; L2/3 and L4, layer 2/3 and layer 4 excitatory neurons; L5/6 and L5/6-CC, layer 5/6 corticofugal projection and cortico-cortical projection neurons; Neu-mat, maturing neurons; Neu-NRGN-I and Neu-NRGN-II, NRGN-expression neurons. **(c)** Expression of four index proteins in differentiating iPSCs, mouse cortex, and HEK293 cells detected by western blot. Molecular weights (KDa) are marked on the left of each blot. Immunoprecipitations using the same antibodies were unsuccessful.

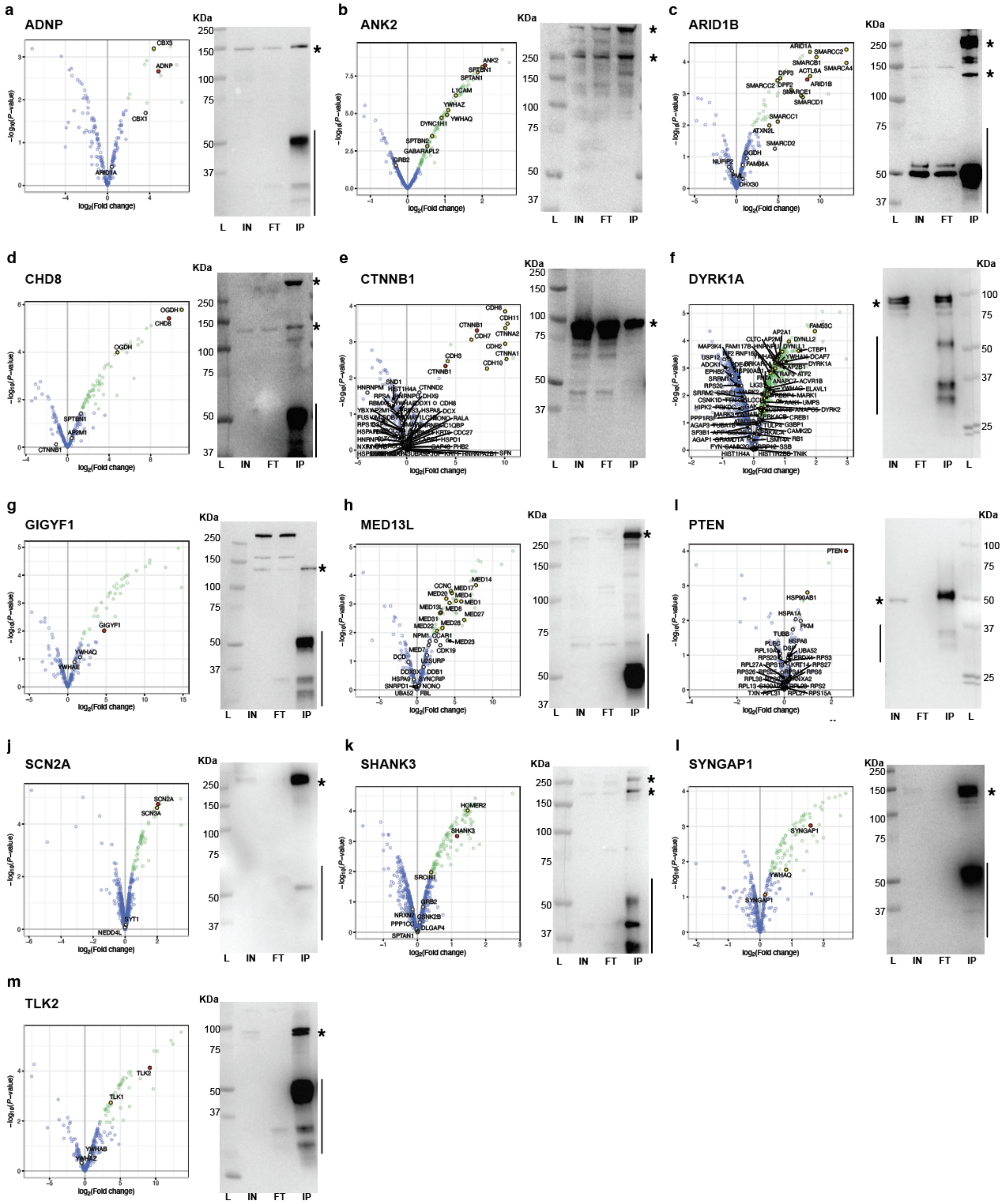

**Supplementary Figure 2. Representative IP-MS data and IP western blots for the 13 index proteins. (a)-(m)** Left: volcano plot of an IP-MS experiment in iNs, showing index protein in red, significant interactors ($\log_2$ FC > 0 and FDR ≤ 0.1) in green, and non-interactors in blue; known

InWeb interactors identified as interactors or non-interactors in the experiment are highlighted in yellow or white, respectively. Right: western blot on an IP of the index protein, whose main isoform(s) are marked with asterisks; IgG heavy and light chains are marked with a vertical line when appropriate; molecular weights are in KDa; L=Ladder, IN=Input, FT=Flow-through, IP=Immunoprecipitation. Noticeably, immunoprecipitation led to enrichment of the signal of full length ARID1B (250KDa), CHD8 (290KDa) and SYNGAP1 (150KDa) in the IP, compared to the starting material (Input).

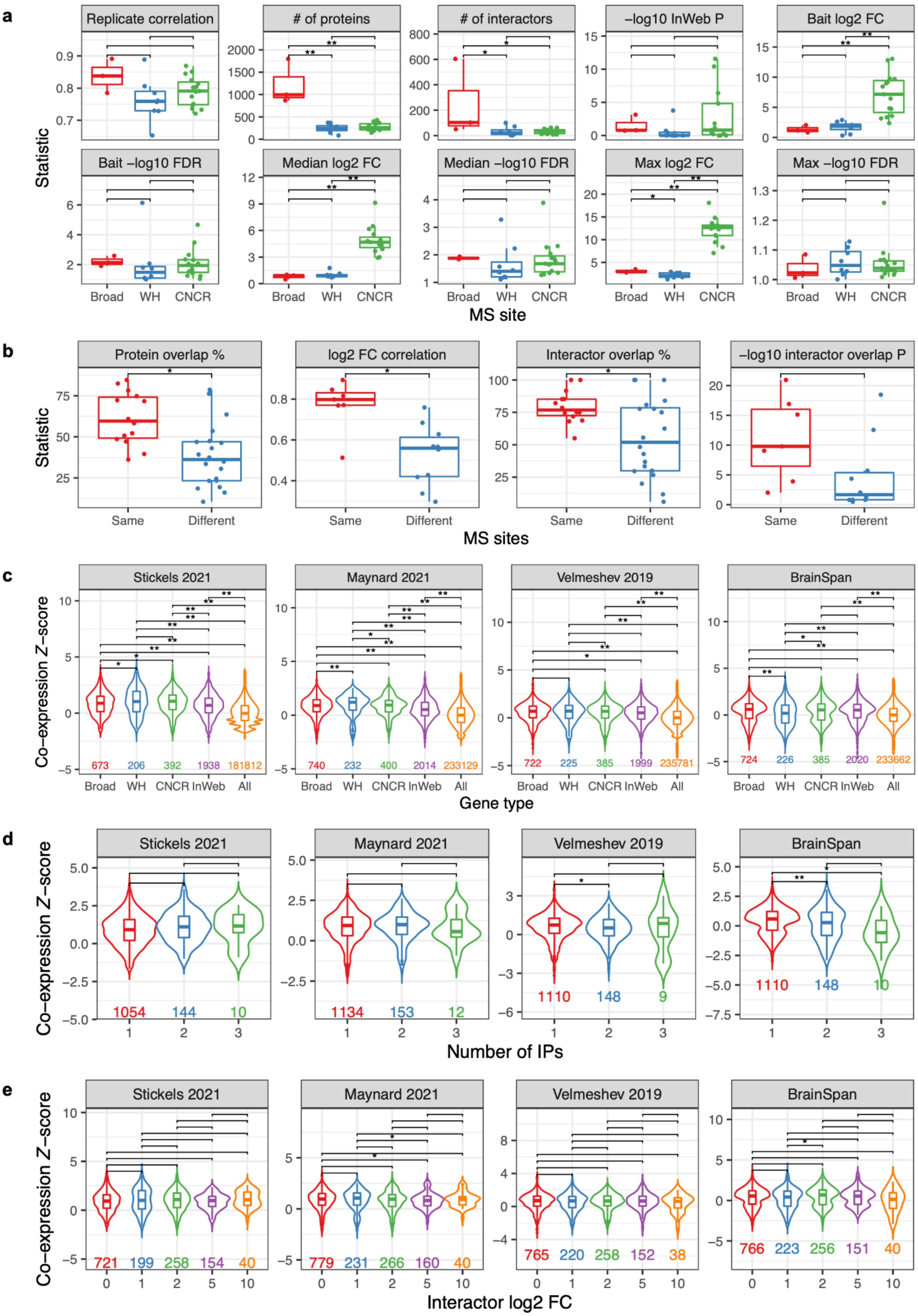
a
Replicate correlation
### of proteins
### of interactors
−log10 InWeb P
Bait log2 FC
Bait −log10 FDR
Median log2 FC
Median −log10 FDR
Max log2 FC
Max −log10 FDR
Statistic
Broad
WH
CNCR
MS site
b
Protein overlap %
log2 FC correlation
Interactor overlap %
−log10 interactor overlap P
Statistic
Same
Different
MS sites
c
Stickels 2021
Maynard 2021
Velmeshev 2019
BrainSpan
Co−expression Z−score
673 206 392 1938 181812
740 232 400 2014 233129
722 225 385 1999 235781
724 226 385 2020 233662
Broad WH CNCR InWeb All
Gene type
d
Stickels 2021
Maynard 2021
Velmeshev 2019
BrainSpan
Co−expression Z−score
1054 144 10
1134 153 12
1110 148 9
1110 148 10
Number of IPs
e
Stickels 2021
Maynard 2021
Velmeshev 2019
BrainSpan
Co−expression Z−score
721 199 258 154 40
779 231 266 160 40
765 220 258 152 38
766 223 256 151 40
0 1 2 5 10
Interactor log2 FC

**Supplementary Figure 3. Comparison of IP-MS datasets across MS sites. (a)** Summary statistics of the 26 QC'ed IP-MS datasets in iNs grouped by MS site. Replicate correlation, $\log_2$ FC correlation between replicates; # of proteins, number of proteins analyzed in Genoppi; # of interactors, number of significant interactors; -log10 InWeb P, $-\log_{10}$ of overlap enrichment P-value between significant interactors and known InWeb interactors; Bait log2 FC or Bait -log10 FDR, $\log_2$ FC or $-\log_{10}$ FDR of the index protein; Median log2 FC, Median -log10 FDR, Max log2 FC, or Max -log10 FDR, median or maximum of the corresponding statistics for significant interactors. **(b)** Comparison of IP pairs of the same index protein, grouped by pairs from the same site vs. different sites. Protein overlap %, percentage of analyzed proteins that overlap with the other IP; log2 FC correlation, $\log_2$ FC correlation of proteins analyzed in both IPs; Interactor overlap %, percentage of interactors that overlap with the other IP; -log10 interactor overlap P, -$\log_{10}$ of overlap enrichment P-value between interactors identified in both IPs. **(c)** Pairwise co-expression Z-scores between index genes and their interactors identified by each MS site (Broad, WH, or CNCR), known InWeb interactors (InWeb), and all protein-coding genes (All). Scores were calculated from spatial transcriptomic datasets in mouse neocortex (Stickels 2021) and human dorsolateral prefrontal cortex (Maynard 2021), single-cell RNA-seq dataset in human cortex (Velmeshev 2019), and the BrainSpan RNA-seq dataset. Number of gene pairs plotted for each gene type is indicated towards the bottom of the plot. **(d)** Pairwise co-expression Z-scores between index genes and their interactors identified by one, two, or three IP-MS datasets. **(e)** Pairwise co-expression Z-scores between index genes and their interactors identified at different $\log_2$ FC thresholds. In all panels, two-tailed Wilcoxon rank-sum tests were performed to compare statistics between groups; single or double asterisks indicate nominally ($P < 0.05$) or Bonferroni-significant ($P < 0.05$/number of pairwise comparisons, as indicated by horizontal brackets) difference, respectively.

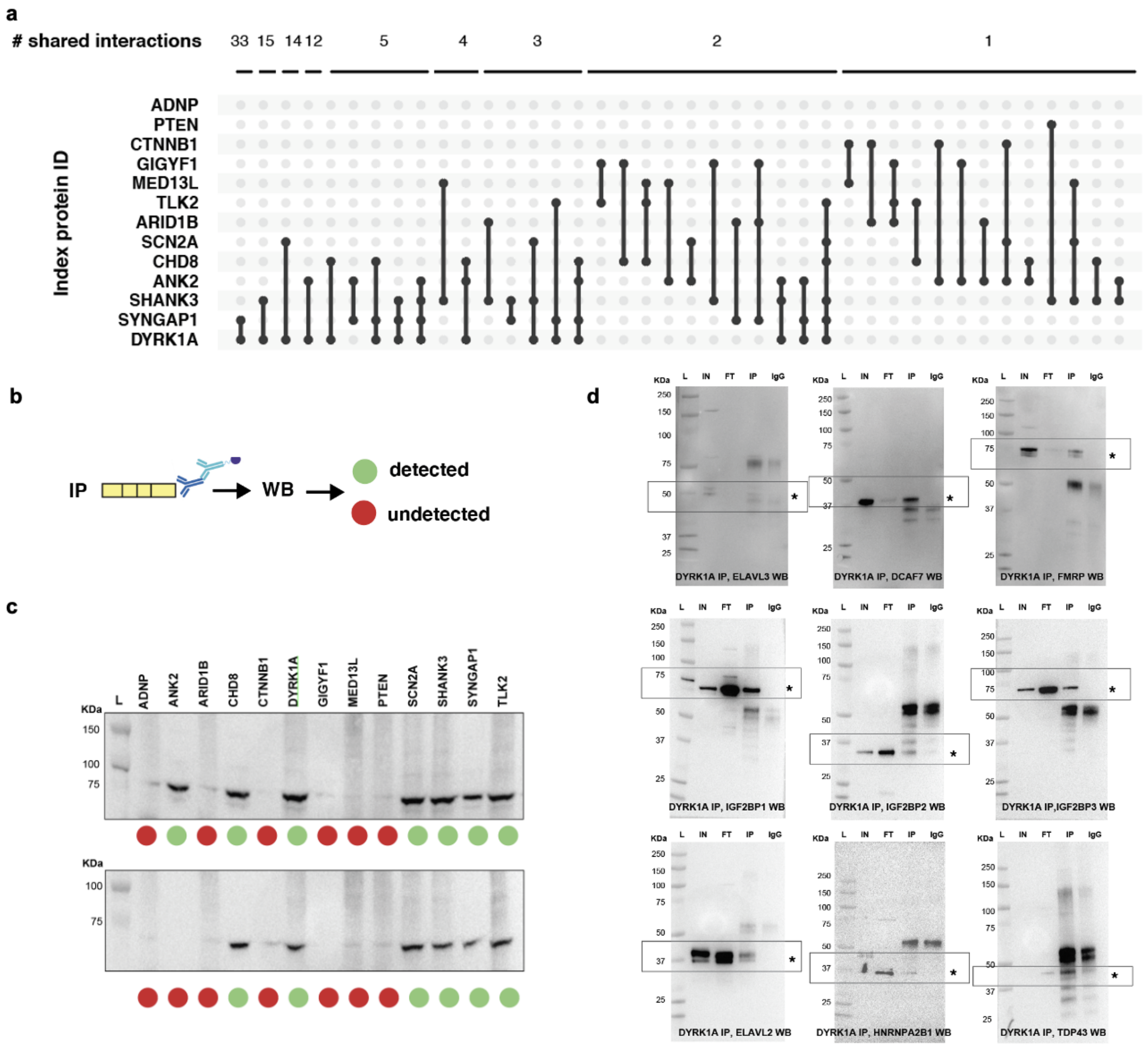


**Supplementary Figure 4. Convergence and IP-WB validation of the ASD PPI network. (a)** Summary of interactors shared between index proteins in the network. **(b)** Schematic of systematic western blot (WB) validation of interactions identified by IP-MS. **(c)** Western blots of two recurrent interactors, IGF2BP1 and IGF2BP3, on IP of each index protein (named at the top) in iNs. The main isoform of IGF2BP1 or IGF2BP3 is marked with an asterisk. Green dots represent observed enrichment of IGF2BP1 or IGF2BP3 by MS in each IP, whereas red dots represent absence of enrichment. **(d)** Western blots of ELAVL3, DCAF7, FMR1 (FMRP), IGF2BP1, IGF2BP2, IGF2BP3, ELAVL2, HNRNPA2B1, and TARDBP (TDP43) on an IP of DYRK1A in iNs. Due to low levels of expression of neuronal TDP-43, we could not detect signal in the Input prior to enrichment by IP. Molecular weights are in KDa. L=Ladder, IN=Input, FT=Flow-through, IP=Immunoprecipitation, IgG=IgG control. An asterisk marks the expected band(s) of each named protein. Boxes highlight the molecular weight where expected signal should be compared across lanes.

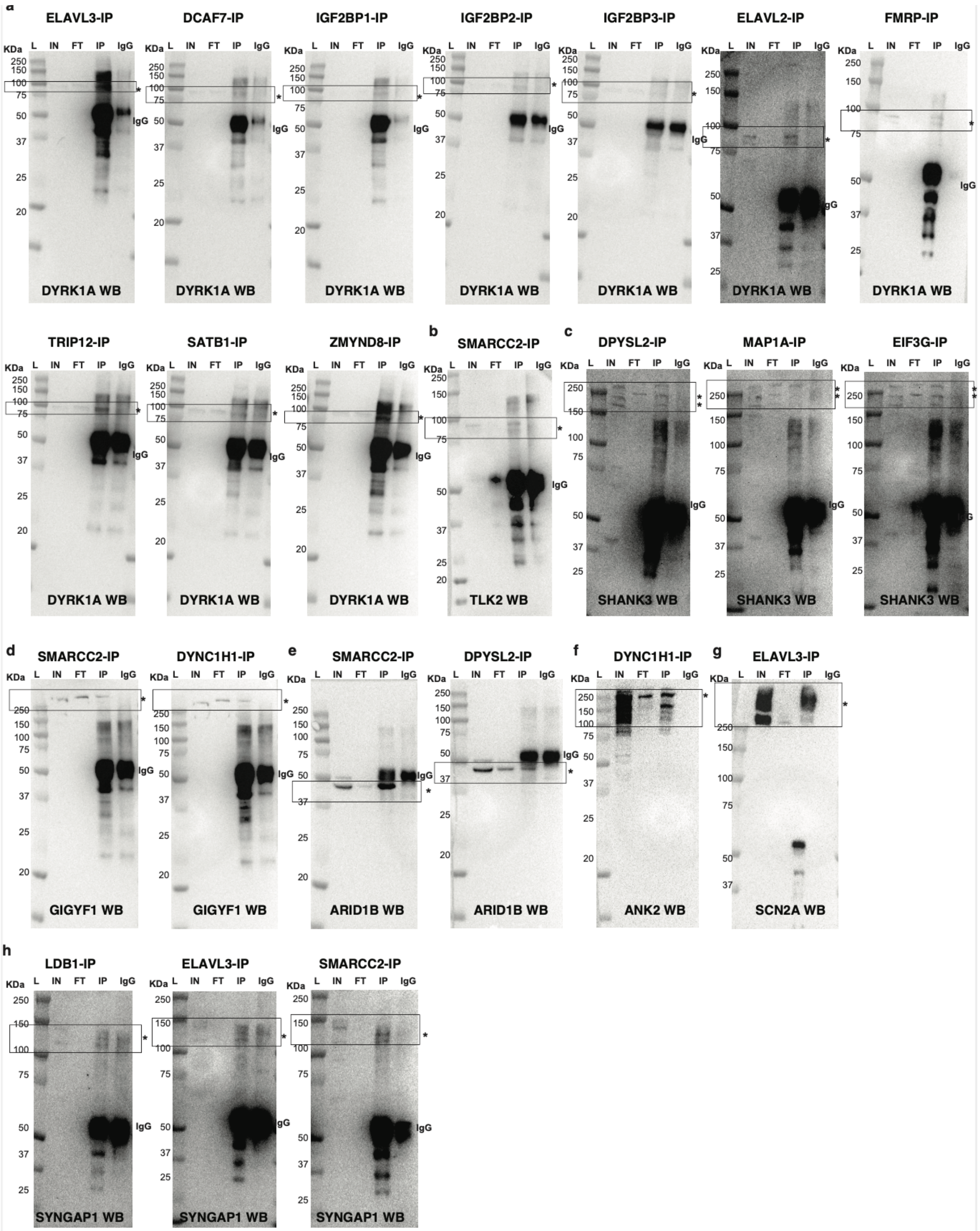
a
ELAVL3-IP
DCAF7-IP
IGF2BP1-IP
IGF2BP2-IP
IGF2BP3-IP
ELAVL2-IP
FMRP-IP
KDa
L
IN
FT
IP
IgG
DYRK1A WB
TRIP12-IP
SATB1-IP
ZMYND8-IP
b
SMARCC2-IP
TLK2 WB
c
DPYSL2-IP
MAP1A-IP
EIF3G-IP
SHANK3 WB
d
SMARCC2-IP
DYNC1H1-IP
GIGYF1 WB
e
SMARCC2-IP
DPYSL2-IP
ARID1B WB
f
DYNC1H1-IP
ANK2 WB
g
ELAVL3-IP
SCN2A WB
h
LDB1-IP
ELAVL3-IP
SMARCC2-IP
SYNGAP1 WB

**Supplementary Figure 5. Western blot analysis on reverse IPs of selected interactors to detect the presence of index proteins.** (a) DYRK1A western blots; (b) TLK2 western blots; (c) SHANK3 western blots; (d) GIGYF1 western blots; (e) ARID1B western blots; (f) ANK2 western blots; (g) SCN2A western blot; (h) SYNGAP1 western blots. Due to low levels of expression, we could only detect a dim signal in the input for DYRK1A and SYNGAP1 prior to enrichment by IP. Molecular weights are in KDa. L=Ladder, IN=Input [10% of the protein lysate utilized for the IP]; FT=supernatant [10% of unbound lysate]; IP=immunoprecipitate; IgG=IgG control. An asterisk marks the expected band(s) of each named protein. Boxes highlight the molecular weight where expected signal should be compared across lanes. The name of the immunoprecipitated protein is on top of each blot.

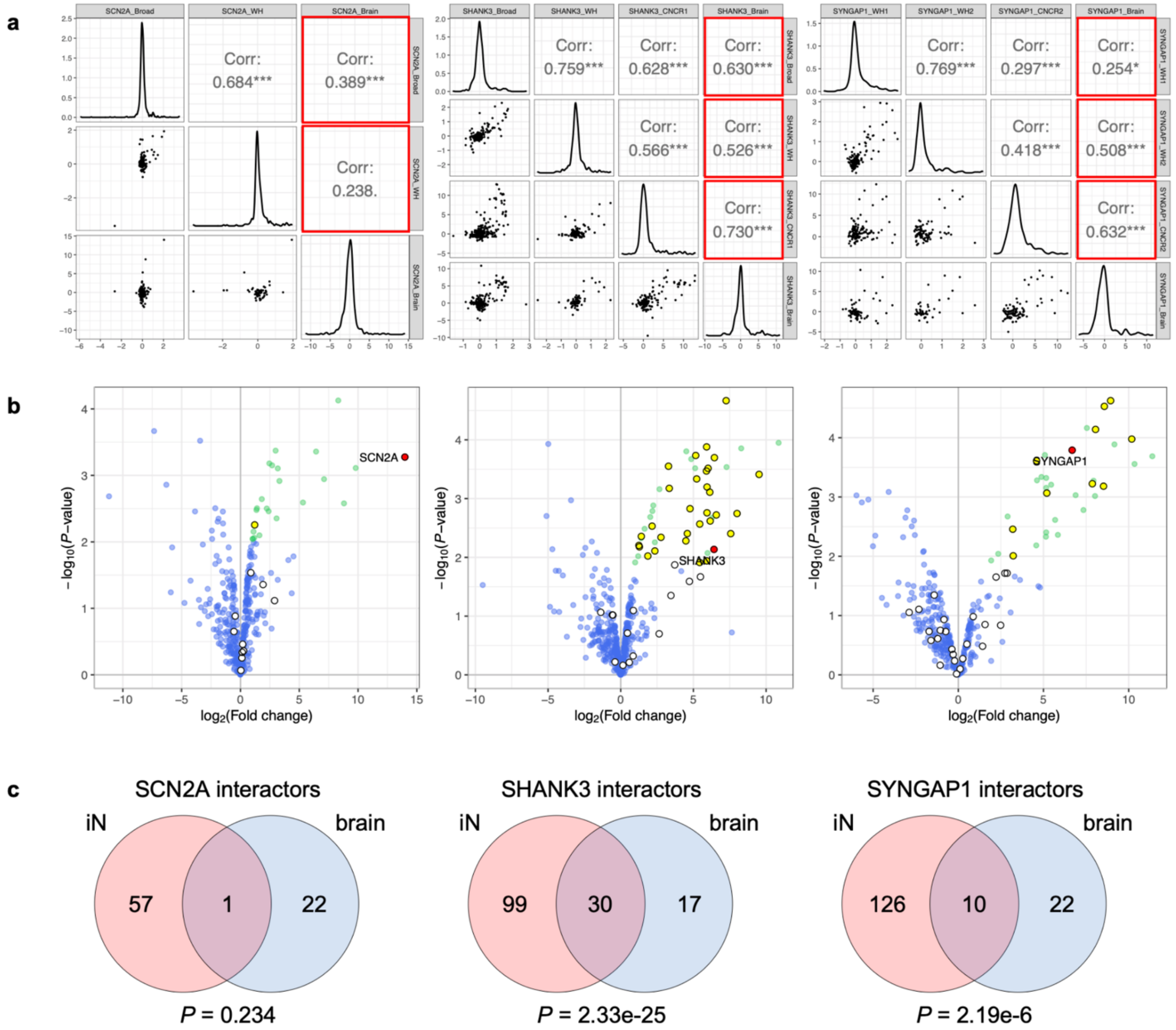


**Supplementary Figure 6. Agreement between iN-derived PPIs and brain IP-MS data. (a)** $Log_2$ FC correlation of proteins analyzed across SCN2A (left), SHANK3 (middle), and SYNGAP1 (right) IP-MS experiments in iNs and human brain homogenates (indicated by 'Brain' suffix in IP name). Triple asterisks indicate Pearson's correlations with $P < 0.001$; correlations with between iN vs. brain data are boxed in red. **(b)** Volcano plots of the brain IP-MS data from (a), showing index protein in red, significant interactors ($\log_2$ FC $> 0$ and FDR $\leq 0.1$) in green, and non-interactors in blue. Interactors or non-interactors that were identified as interactors in the analogous iN-derived IPs are highlighted in yellow or white, respectively. **(c)** Venn diagrams showing the overlap between the interactors found in iNs vs. brain homogenates. Overlap enrichment P-values were calculated using one-tailed hypergeometric tests that only considered the population of proteins detected in both iN and brain IPs.

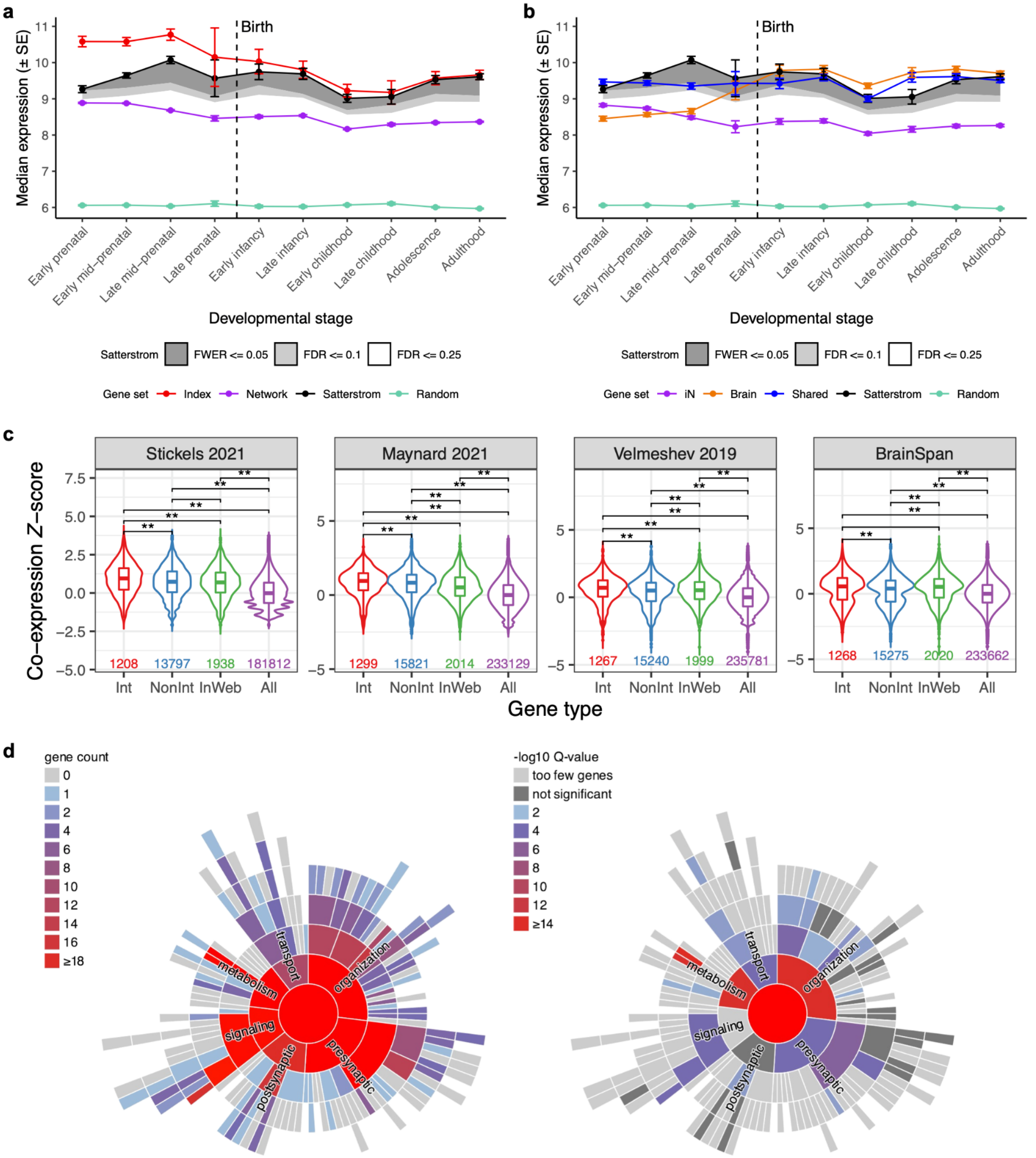


**Supplementary Figure 7. Tissue, cell type, and SynGO enrichment of the ASD PPI network. (a)** Frontal cortex RNA expression of gene sets across ten developmental stages. Median expression and standard error (SE) of each gene set were derived from the BrainSpan exon microarray dataset. Gene sets plotted include: Index, index genes used to generate the PPI network;

Network, interactor genes in the network (excluding index genes); Satterstrom, FWER $\leq$ 0.05 genes in Satterstrom *et al.*; Random, random genes from the BrainSpan dataset. Shaded regions indicate median expression of genes reaching FWER $\leq$ 0.05, FDR $\leq$ 0.1, or FDR $\leq$ 0.25 in Satterstrom *et al.*, with darker grey indicating greater significance. **(b)** Frontal cortex RNA expression of SCN2A, SHANK3, and SYNGAP1 interactors identified only in iN-derived IP-MS data (iN), only in brain-derived IP-MS data (Brain), or in both (Shared). Other plotted gene sets are the same as described in (a). **(c)** Pairwise co-expression Z-scores between index genes and their interactors (Int), non-interactors (NonInt), known InWeb interactors (InWeb), and all protein-coding genes (All). Scores were calculated from spatial transcriptomic datasets in mouse neocortex (Stickels 2021) and human dorsolateral prefrontal cortex (Maynard 2021), single-cell RNA-seq dataset in human cortex (Velmeshev 2019), and the BrainSpan RNA-seq dataset. Single or double asterisks indicate nominally ($P$ < 0.05) or Bonferroni-significant ($P$ < 0.05/6, adjusting for 6 pairwise comparisons) difference in score distribution, respectively, as calculated by two-tailed Wilcoxon rank-sum tests. Number of gene pairs plotted for each gene type is indicated towards the bottom of the plot. **(d)** SynGO analysis of synaptic genes in the PPI network; the sunburst plots show the number (left) and the enrichment (right) of genes distributed over a range of biological processes of the synapse.

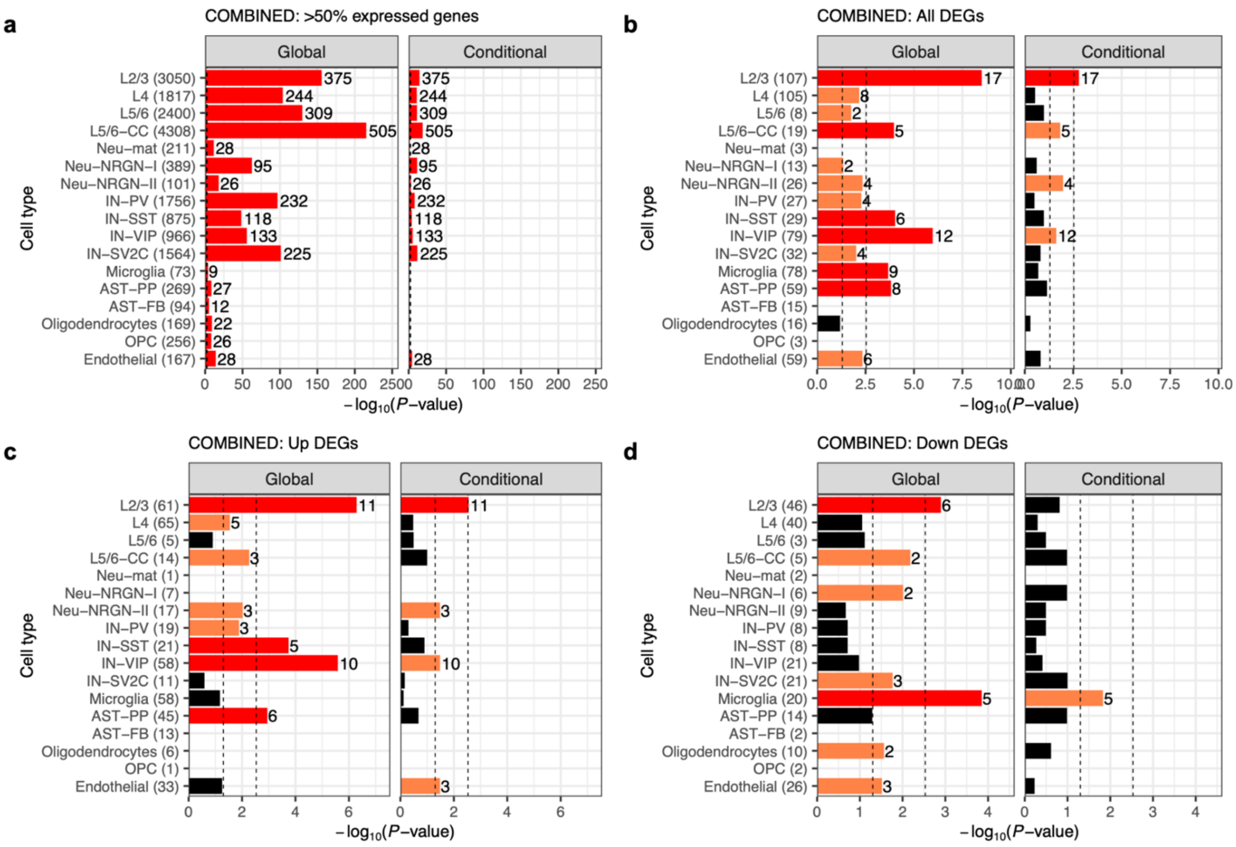


**Supplementary Figure 8. Cell type enrichment of the ASD PPI network. (a)** Global or conditional overlap enrichment between the network and genes expressed in > 50% of cells in each cell type in *post mortem* cortex of ASD patients and controls. **(a)-(c)** Global or conditional overlap enrichment between the network and all (a), up- (b) or down-regulated (c) differentially expressed genes in each cell type in *post mortem* cortex of ASD patients compared to controls. Nominally ($P$ < 0.05) or Bonferroni-significant ($P$ < 0.05/17, adjusting for 17 cell types) results are shown in orange or red, respectively. Cell type abbreviations: AST-FB and AST-PP, fibrous and protoplasmic astrocytes; OPC, oligodendrocyte precursor cells; IN-PV, IN-SST, IN-SV2C, and IN-VIP, parvalbumin, somatostatin, SV2C, and VIP interneurons; L2/3 and L4, layer 2/3 and layer 4 excitatory neurons; L5/6 and L5/6-CC, layer 5/6 corticofugal projection and cortico-cortical projection neurons; Neu-mat, maturing neurons; Neu-NRGN-I and Neu-NRGN-II, NRGN-expression neurons.

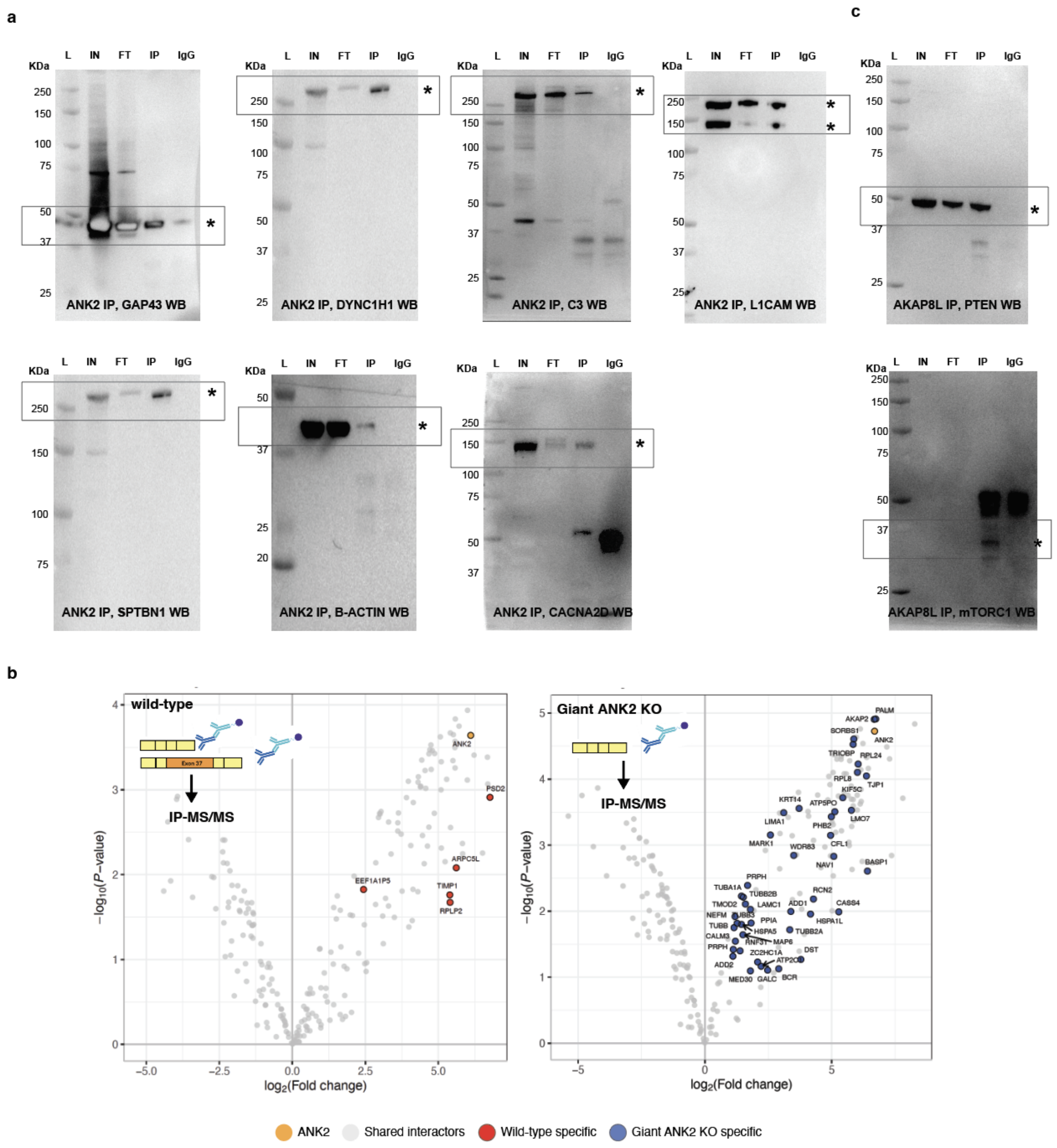


**Supplementary Figure 9. Follow-up validations and experiments for the ANK2 PPIs and the PTEN-AKAP8L interaction. (a)** Western blots of GAP43, DYNC1H1, C3, L1CAM, SPBTN1, ACTB (B-ACTIN), and CACNA2D1 (CACNA2D) on an IP of ANK2 in iNs. IN=Input, FT=Flow-through, IP=Immunoprecipitation, IgG=IgG control, H+L Igg=Heavy and light IgG chains. An asterisk marks the expected band of each named protein. **(b)** Volcano plots of IP-MS experiments for ANK2 in wild-type (left) and giant ANK2 KO (right) NPCs. ANK2 is shown in orange, wild-type-specific interactors (i.e., $\log_2$ FC > 0 and FDR ≤ 0.1 in wild-type data only) are in red, and giant ANK2 KO-specific interactors (i.e., $\log_2$ FC > 0 and FDR ≤ 0.1 in KO data only) are in blue. **(c)** Western blots of PTEN (above) and mTORC1 (below) on an IP of AKAP8L in iNs. Molecular weights are in KDa. L=Ladder, IN=Input, FT=Flow-through, IP=Immunoprecipitation, IgG=IgG control. An asterisk marks the expected band(s) of each named protein. Boxes highlight the molecular weight where expected signal should be compared across lanes.

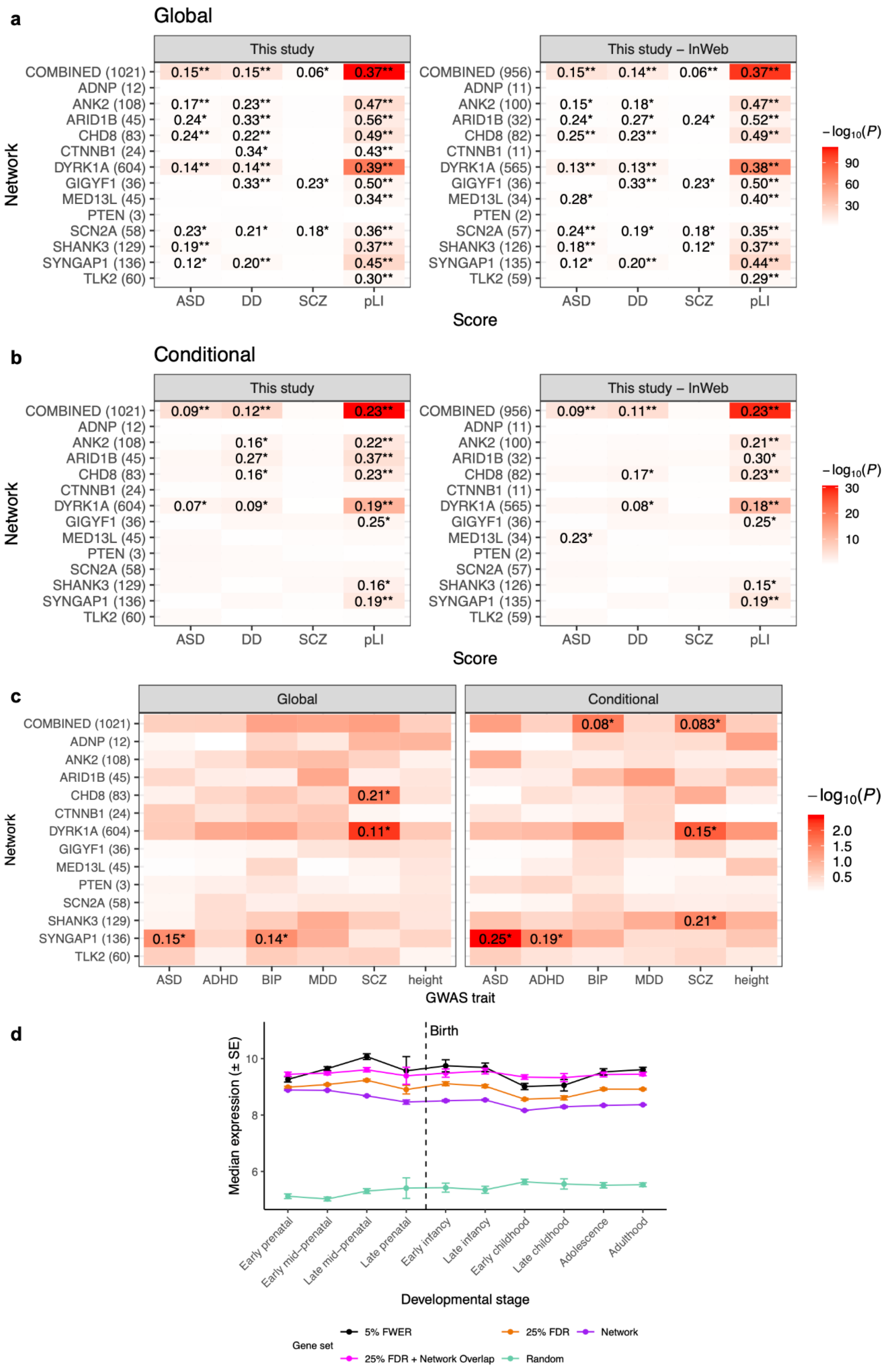

a
Global
This study
This study – InWeb
Network
Score
COMBINED (1021)
ADNP (12)
ANK2 (108)
ARID1B (45)
CHD8 (83)
CTNNB1 (24)
DYRK1A (604)
GIGYF1 (36)
MED13L (45)
PTEN (3)
SCN2A (58)
SHANK3 (129)
SYNGAP1 (136)
TLK2 (60)
ASD
DD
SCZ
pLI
−log10(P)
b
Conditional
c
Global
Conditional
GWAS trait
ADHD
BIP
MDD
height
d
Birth
Median expression (± SE)
Developmental stage
Early prenatal
Early mid-prenatal
Late mid-prenatal
Late prenatal
Early infancy
Late infancy
Early childhood
Late childhood
Adolescence
Adulthood
Gene set
5% FWER
25% FDR
Network
25% FDR + Network Overlap
Random

**Supplementary Figure 10. Rare variant enrichment, common variant enrichment, and BrainSpan expression of the ASD PPI network. (a)-(b)** Rare variant or pLI score enrichment of the network or index protein-specific sub-networks compared to the rest of the genome (Global; a) or to non-interactors (Conditional; b), with (This study) and without (This study - InWeb) including known InWeb interactors. The number of genes in each network is shown in parentheses on the y-axis. Enrichment P-values were calculated using one-tailed Kolmogorov-Smirnov tests and gene-based association statistics from ASD, DD, or SCZ exome sequencing data or gnomAD pLI scores. KS test statistics reaching nominal ($P < 0.05$) or Bonferroni ($P < 0.05/14$, adjusting for 14 networks) significance are shown in the heat map followed by single or double asterisks, respectively. **(c)** Common variant enrichment of the network or index protein-specific sub-networks compared to the rest of the genome (Global) or to non-interactors (Conditional). Enrichment P-values were calculated using MAGMA and GWAS data for ASD, ADHD, BIP, SCZ, and height. Enrichment coefficients reaching nominal significance are shown in the heat map followed by an asterisk. **(d)** Frontal cortex RNA expression of gene sets across ten developmental stages. Median expression and standard error (SE) of each gene set were derived from the BrainSpan exon microarray dataset. Gene sets plotted include: 5% FWER and 25% FDR, FWER ≤ 0.05 and FDR ≤ 0.25 genes in Satterstrom *et al.*; Network, interactor genes in the PPI network; 25% FDR + Network Overlap, overlap between the 2 gene sets; Random, random genes from the BrainSpan dataset.

**Captions for Supplementary Tables 1-12**

**Supplementary Table 1. Details of index protein antibodies that have been evaluated for IP.**

**Supplementary Table 2. IP-MS datasets analyzed in this study.**

**Supplementary Table 3. Summary of interactors and non-interactors in individual IP-MS datasets and consolidated PPI networks.**

**Supplementary Table 4. Comparison of IPs of the same index protein.**

**Supplementary Table 5. Summary of interactions tested in forward or reverse IP-WB validation experiments.**

**Supplementary Table 6. Tissue, cell type, and SynGO enrichment analyses for the combined PPI network.**

**Supplementary Table 7. Enriched GO cellular component (CC) terms for ANK2 interactors identified exclusively in wild-type (WT) or giant ANK2 KO NPCs.**

**Supplementary Table 8. Enrichment analyses for the IGF2BP1-3 targets.**

**Supplementary Table 9. Rare variant and pLI score enrichment of the PPI networks.**

**Supplementary Table 10. Common variant risk enrichment of the PPI networks estimated using GWAS data and MAGMA.**

**Supplementary Table 11. Genetic variants of IGF2BP1-3 and their associations to neuropsychiatric disorders.**

**Supplementary Table 12. IP-MS data files deposited to MassIVE.**
